# Global Excess Maternal Mortality During the COVID-19 Pandemic: Country-Level Counterfactual Estimates and Associated Factors

**DOI:** 10.64898/2026.06.30.26356947

**Authors:** Jaimini Hitendra Gajjar, Hamed Karami, Holly A. Hayes, Meredith A. Dixon, Greta M. Massetti, Gerardo Chowell

## Abstract

**Synopsis:** *Study question:* How did maternal deaths and MMR during 2020-2023 depart from pre-pandemic country trajectories, and were those departures associated with reported disruptions in antenatal, facility-based delivery, or postnatal care?

*What is already known:* Maternal mortality was changing before COVID-19; infection and service disruption increased maternal risk, but simple pre/post comparisons cannot separate these influences from underlying trends.

*What this study adds:* Across 195 countries and territories, we estimated 68,489 cumulative excess maternal deaths, with statistically detectable excess in 113. In 39 self-selected countries, broad national disruption measures were not clearly associated with signed MMR deviations, highlighting heterogeneity and the need for more granular measures of access and quality.

**Background:** COVID-19 increased maternal risk through infection and disruption of essential services, but maternal mortality was already changing before 2020, making simple pre-pandemic comparisons difficult.

**Objectives:** To estimate country-level departures in maternal deaths and maternal mortality ratios (MMR) during 2020-2023 from projected pre-pandemic trajectories and examine whether maternal health-service disruptions were associated with MMR deviations.

**Methods:** We fitted country-specific counterfactual models to WHO/UN MMEIG estimates from 2000-2019 and projected maternal deaths and MMR through 2023 under a no-pandemic scenario. Excess was defined as the positive difference between the MMEIG estimate and counterfactual value. In a self-selected subset of 39 countries (63 country-year observations), we modelled log(MMEIG-estimated MMR/counterfactual MMR) as a function of three service-disruption measures and four baseline or year covariates.

**Results:** Across 195 countries and territories, cumulative excess maternal deaths were estimated at 68,489 (95% UI 34,706, 147,118); 113 had statistically detectable cumulative excess. The largest regional burdens were in South-East Asia, the Eastern Mediterranean, and Africa. In the 39-country subset, antenatal care, facility-based delivery, and postnatal care disruption were not clearly associated with signed MMR deviation in the full or separate models. No single country materially changed the estimated coefficients in leave-one-country-out analyses, and excluding Namibia, which had the two most unusual observations, did not alter the conclusions.

**Conclusions:** Maternal mortality departed substantially but unevenly from pre-pandemic trajectories. The absence of associations with broad national disruption measures should not be interpreted as evidence that service disruption was unimportant; it instead highlights the need for more detailed measures of service access and quality, alongside protection of the maternal-care continuum during emergencies.

## Background

Maternal mortality remains a major public health challenge. WHO/UN Maternal Mortality Estimation Inter-Agency Group (MMEIG) estimates indicate that approximately 260,000 maternal deaths occurred globally in 2023, corresponding to a maternal mortality ratio (MMR) of 197 deaths per 100,000 live births.^1^ Although global MMR declined substantially between 2000 and 2023, progress slowed before the pandemic and remains insufficient to meet Sustainable Development Goal 3.1.^1,2^ Persistent differences across countries reflect variation in access to care, quality of services, health-system capacity, and broader structural conditions.^3,4^ COVID-19 introduced both direct and indirect risks to maternal health. Pregnant individuals with SARS-CoV-2 infection faced higher risks of severe disease and adverse maternal and neonatal outcomes.^5,6^ This direct clinical risk is important when interpreting pandemic-period maternal mortality because higher mortality could reflect infection itself, reduced or delayed obstetric care, or both pathways operating together. At the same time, the pandemic disrupted antenatal, intrapartum, and postnatal care, reduced facility-based service use, strained the health workforce and supply chains, and interrupted referral systems in many settings.^7–10^ Vaccination became available during the pandemic, but guidance and access for pregnant populations varied across countries, adding another source of difference in maternal risk during this period.^11^

Global maternal mortality analyses have described trends through 2023 and the burden of COVID-19 infection among pregnant women.^12^ However, maternal mortality was already changing before 2020, so pandemic-period values are difficult to interpret through simple comparisons with the immediately preceding year. A counterfactual approach can instead compare pandemic-period estimates with the trajectory expected from pre-pandemic trends.^13,14^ Studies of service utilisation also reported substantial variation across countries and services, making it uncertain whether national disruption measures would track mortality deviations consistently.^7,8^ This distinction matters because the same disruption category can represent very different service losses depending on baseline coverage and health-system capacity. It therefore remains unclear whether countries reporting greater disruption of maternal health services experienced larger departures from their expected MMR trajectories. We addressed two related questions. First, we estimated country-level deviations in maternal deaths and MMR during 2020-2023 relative to counterfactual projections calibrated to WHO/UN MMEIG trends from 2000-2019. Second, in a self-selected subset of 39 countries, we examined the log ratio of MMEIG-estimated to counterfactual MMR and evaluated antenatal care disruption, facility-based delivery disruption, postnatal care disruption, baseline MMR, antenatal care coverage, skilled birth attendance, and year. The country-level regression was exploratory and ecological rather than causal, and estimated excess should be interpreted as departure from a modelled no-pandemic baseline rather than as deaths caused solely by COVID-19.

## Methods

### Study design, data sources and analytic samples

We conducted a country-level ecological counterfactual modelling study with a nested exploratory analysis of service-disruption correlates. We used publicly available country-level estimates from Trends in Maternal Mortality 2000 to 2023, produced by the WHO/UN Maternal Mortality Estimation Inter-Agency Group (MMEIG), including annual maternal deaths and maternal mortality ratios (MMR) for 195 countries and territories.^1^ Maternal deaths and MMR were analysed as complementary outcomes: deaths describe absolute burden, while MMR describes maternal mortality relative to live births. The WHO/UN MMEIG estimates combine civil registration and vital statistics, surveys, censuses and specialised studies, with statistical adjustment for underreporting, misclassification and differences between data sources.^1,15,16^ In the 2025 MMEIG report, 2020 to 2022 were treated as COVID years using a modified COVID BMat setup, whereas 2023 used the default BMat framework.^1^ Ethics approval was not required because this study used publicly available aggregated country-level estimates and did not involve clinical or patient data.

All countries and territories with available MMEIG estimates were included in the counterfactual analysis and were grouped by WHO region. For descriptive figures, we highlighted Afghanistan, South Sudan, Chad, Nigeria, Guinea-Bissau, Liberia, Somalia and Lesotho. This selection was for visualisation only and did not affect model fitting or global estimates.

### Exposures and covariates for the explanatory analysis

To examine factors associated with departures from expected MMR, we assembled a separate country-year dataset. Measures of antenatal care disruption, facility-based delivery disruption and postnatal care disruption came from the WHO Global Pulse Survey on Continuity of Essential Health Services for 2021 and 2022.^17^ Pre-pandemic antenatal care coverage was measured as the percentage of women receiving at least four antenatal care visits, using the latest available pre-2020 country value from UNICEF data.^18^ Skilled birth attendance was the percentage of births attended by skilled health personnel, using the latest available pre-2020 value from the joint UNICEF/WHO database.^19^ Baseline MMR was the 2019 WHO/UN MMEIG estimate, and year was included to account for differences between 2021 and 2022.

We chose these seven predictors because they represent the main maternal care pathways highlighted in the pandemic literature. Multicountry studies documented disruptions in antenatal care, delivery services and postnatal care during COVID-19 and linked reductions in service use with adverse maternal and child health consequences.^7,8^ Antenatal care coverage and skilled birth attendance were included because they are established measures of access to care during pregnancy and childbirth and are closely connected to maternal mortality reduction.^2,20^ Baseline MMR accounted for the maternal mortality level already present before the pandemic, while year accounted for changing pandemic conditions and service recovery over time.^17^ We did not add a larger set of socioeconomic, policy or health-system indicators because the aim was to test factors directly related to maternal care and the available sample was modest. Pregnancy-specific COVID-19 vaccination coverage was also not included because comparable coverage was not consistently available across the countries and years in this analysis. The temporal ordering and links considered when selecting predictors are summarised in eFigure 1. The model was designed to describe associations and not to identify causal effects.

### Missing data

The explanatory analysis used complete country-year records with the outcome and all seven prespecified predictors. No missing predictor values were imputed. The intersection of the data sources yielded 63 country-year observations from 39 countries: 31 observations in 2021 and 32 in 2022, with 24 countries contributing both years and 15 contributing one year. The analysis was limited to 2021 and 2022 because all three service disruption measures could be aligned with annual MMR estimates in those years.

### Counterfactual modelling and excess definitions

We calibrated models separately to maternal deaths and MMR using WHO/UN MMEIG estimates from 2000 to 2019, excluding pandemic-period information. We used the established SubEpiPredict ensemble n-sub-epidemic framework to represent nonlinear pre-pandemic trends and generate counterfactual projections for 2020 to 2023.^21,22^ For each country and outcome, one- and two-component models were considered, model selection used the corrected Akaike Information Criterion, and uncertainty was quantified with 300 bootstrap resamples. Normal and Poisson error structures were considered. Full mathematical details and implementation settings are provided in Supplementary Section S1.

For each year from 2020 to 2023, excess maternal deaths were defined as the positive difference between the WHO/UN MMEIG estimate and the median counterfactual forecast. Negative deviations were set to zero because the primary analysis was intended to summarise additional above-baseline burden rather than net deviations in both directions. This positive-only operational definition has been used in prior applications of the same counterfactual approach to tuberculosis mortality, tuberculosis incidence, and drug overdose mortality.^23–25^ A related HIV/AIDS excess mortality study used the same observed versus expected counterfactual framework while retaining signed differences.^26^ In the present study, signed departures were therefore retained separately in the explanatory analysis through log(observed/expected MMR). Cumulative excess deaths were obtained by summing annual estimates and their corresponding bounds. Excess MMR was calculated in the same way and summarised as the sum of annual MMR deviations; this aggregate should not be interpreted as a single-year or population-weighted MMR.

### Explanatory regression analysis

For the explanatory analysis, the outcome was the log ratio of the WHO/UN MMEIG MMR to the expected counterfactual MMR, log(observed/expected). This measure retains both positive and negative departures from the expected trajectory, unlike the positive-only excess measure used for the primary burden estimates. WHO disruption categories were coded by severity: 0 for no or minimal disruption, defined as any decrease of less than 5% or any increase; 1 for a 5% to 25% decrease; 2 for a 26% to 50% decrease; and 3 for a decrease greater than 50%.

Baseline MMR was log-transformed, and 2021 was the reference year. Each disruption measure was entered as a single linear term across these ordered severity categories, a parsimonious specification chosen because the modest sample size could not support separate coefficients for each category.

We fitted a linear mixed-effects regression with a country-specific random intercept to account for repeated annual observations within countries. Because each country contributed at most two annual observations, the random intercept served to account for within-country correlation rather than to estimate between-country variance precisely. The initial model included antenatal care disruption, facility-based delivery disruption, postnatal care disruption, baseline MMR, antenatal care coverage, skilled birth attendance and year. Predictor correlations and variance inflation factors were examined before model selection. Starting with all seven predictors, backward selection based on Akaike Information Criterion removed a predictor only when its removal improved model fit; year and the country random intercept were retained throughout. Coefficients are presented with 95% confidence intervals, and the analysis is interpreted as associational rather than causal.

### Sensitivity analyses

Because the three service disruption measures were strongly correlated, three additional models examined each disruption measure separately with the same baseline covariates. A leave-one-country-out analysis then refitted the model after removing each country in turn to assess whether any single country drove the coefficients. The model was also repeated without Namibia because its 2021 and 2022 observations had the two largest standardised residuals and were the only observations above an absolute residual threshold of 2. These checks assessed robustness and were not used to exclude observations from the primary analysis.

## Results

### Counterfactual and excess maternal mortality

Figure 1 shows substantial heterogeneity in MMR departures from the counterfactual trajectories. Afghanistan, Somalia, Guinea-Bissau and Lesotho had WHO/UN MMEIG estimates above the median counterfactual projection during at least part of 2020-2023, whereas Nigeria, Liberia, South Sudan and Chad were generally close to or below the projected trajectories.

**Figure 1.**
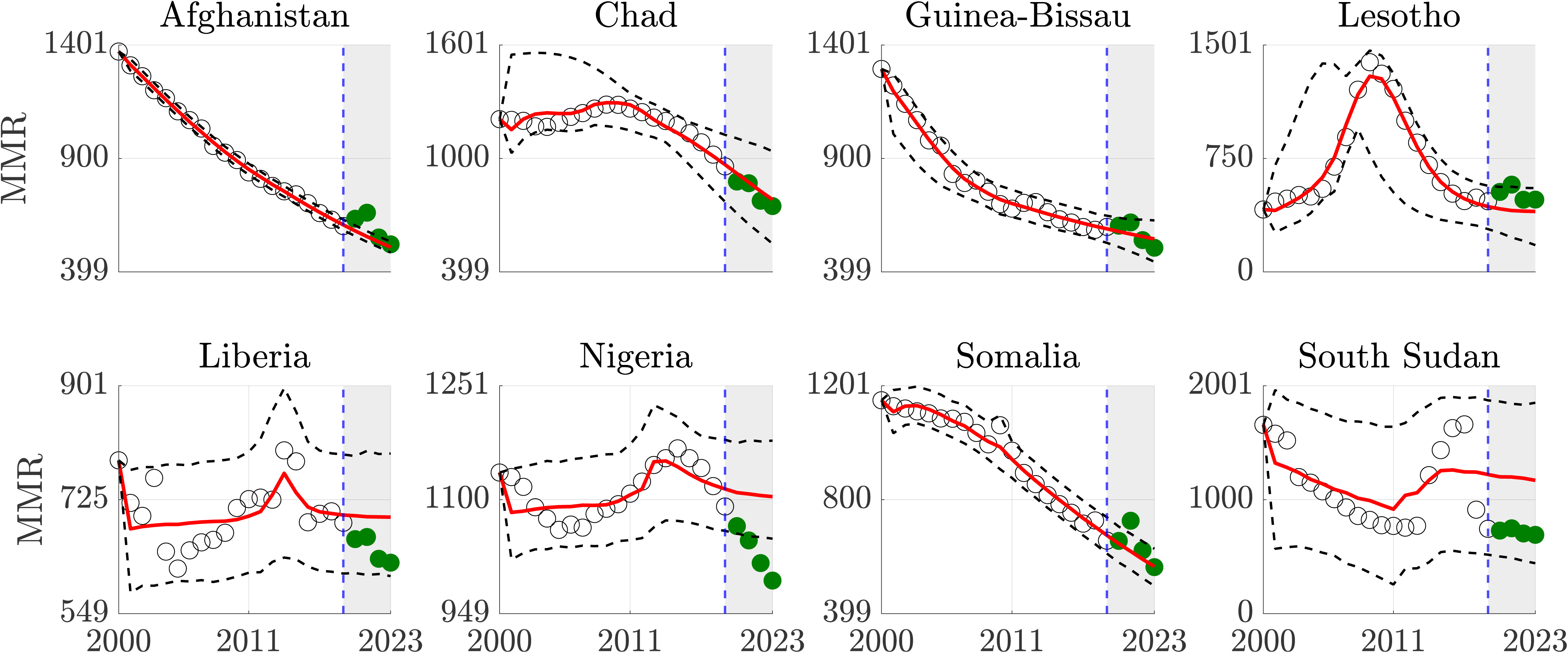
Calibration and forecasting of annual maternal mortality ratio (MMR) from 2000 to 2023 for eight illustrative high-burden countries: Afghanistan, South Sudan, Chad, Nigeria, Guinea-Bissau, Liberia, Somalia and Lesotho. The model was fitted to pre-pandemic WHO/UN MMEIG estimates from 2000-2019 and used to forecast MMR during 2020-2023 under a counterfactual no-pandemic scenario. Red lines show median model predictions, black dashed lines indicate 95% prediction intervals, hollow black circles represent WHO/UN MMEIG estimates during the calibration period, and green filled circles denote WHO/UN MMEIG estimates during the forecast period.

Among the eight illustrative settings, the lower 95% UI for cumulative excess MMR was above zero in Lesotho, 437 deaths per 100,000 live births (95% UI 14 to 1192), Afghanistan, 192 (111 to 284), and Somalia, 149 (46 to 412) (Table 1). Guinea-Bissau had a positive median estimate of 77 (0 to 357), while Chad, South Sudan, Liberia and Nigeria had median estimates of zero with positive upper bounds in several settings.

**Table 1.**
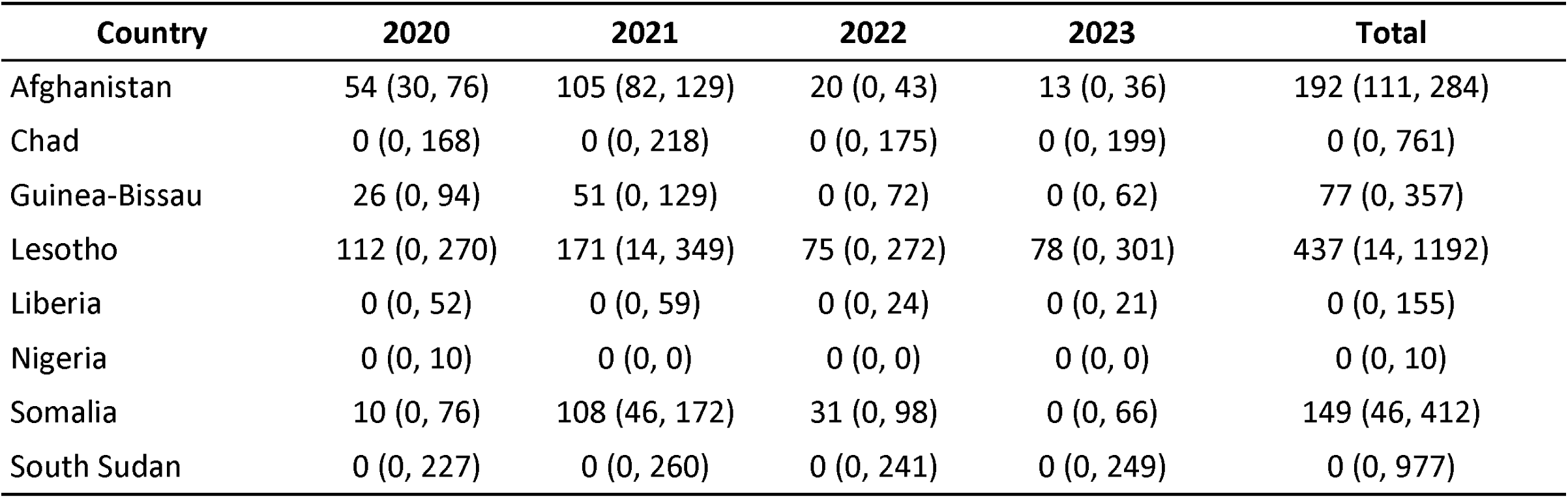
Estimated annual and cumulative excess maternal mortality ratio (MMR) from 2020 to 2023 in eight illustrative high-burden countries. Values are shown as median estimates with 95% uncertainty intervals. Cumulative excess MMR is the sum of annual deviations and should not be interpreted as a single-year rate.

For maternal deaths, Afghanistan and Somalia had the largest cumulative excess burdens among the illustrative countries, with 2,335 deaths (95% UI 1,148 to 4,350) and 1,815 (639 to 3,401), respectively (eTable 1). The lower 95% UI was above zero in both countries. Liberia, Guinea-Bissau and Lesotho had positive median cumulative estimates with intervals that included zero, while Chad and South Sudan had median estimates of zero but positive upper bounds.

Across all 195 modelled countries and territories, cumulative excess maternal deaths during 2020-2023 were estimated at 68,489 (95% UI 34,706 to 147,118), and aggregate excess MMR was 10,154 (4,568 to 23,744). The largest regional excess death burdens were in South-East Asia, the Eastern Mediterranean and Africa (Table 2). Overall, the lower 95% UI was above zero for cumulative excess maternal deaths in 113 countries and for aggregate excess MMR in 133 countries. Figure 2 shows that absolute excess deaths were concentrated mainly in parts of sub-Saharan Africa and South Asia, while aggregate excess MMR highlighted larger risk departures across parts of sub-Saharan Africa and Latin America. Additional regional and country-level counterfactual and excess mortality results are presented in eFigures 4–80.

**Figure 2.**
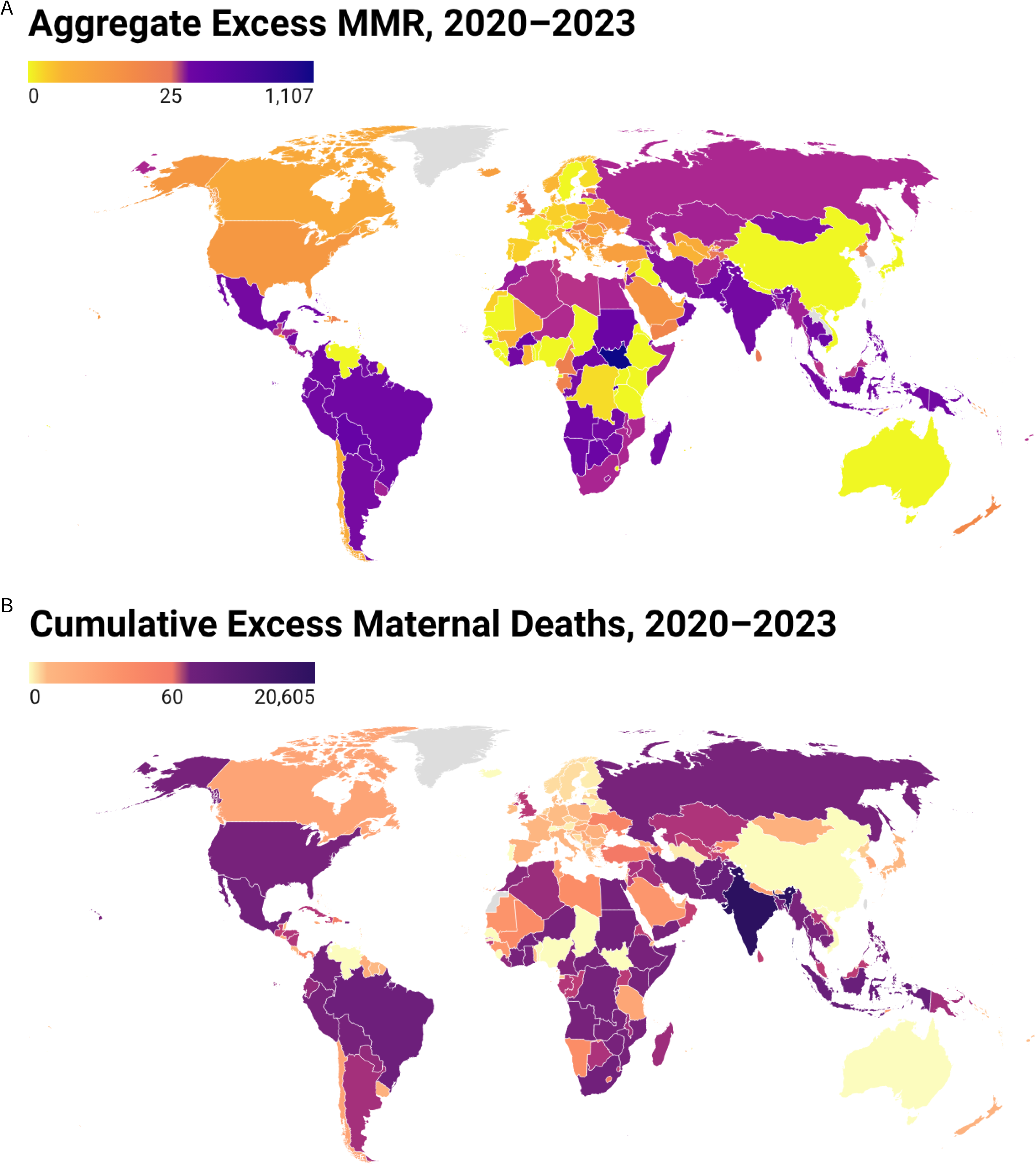
Global maps of excess maternal mortality during 2020-2023. Panel A shows aggregate excess maternal mortality ratio (MMR), calculated as the sum of annual excess MMR deviations across 2020-2023. Panel B shows cumulative excess maternal deaths over the same period. Country-level estimates were derived using an ensemble n-sub-epidemic modelling framework calibrated to pre-pandemic WHO/UN MMEIG estimates from 2000-2019 and projected through 2020-2023 under a counterfactual no-pandemic scenario. Darker shades indicate higher values. Countries not included in the analytic dataset or not mapped are shown in grey. The maps were generated using Datawrapper.

**Table 2.** Global and WHO-region cumulative excess maternal deaths and aggregate excess maternal mortality ratio (MMR) during 2020-2023. Aggregate excess MMR is the sum of country-level annual deviations and should not be interpreted as a single-year or population-weighted regional MMR. Values are median estimates with 95% uncertainty intervals.

| Region | Countries included | Cumulative excess maternal deaths (95% UI) | Aggregate excess MMR (95% UI) | Countries with excess deaths | Countries with excess MMR |
| --- | --- | --- | --- | --- | --- |
| Global | 195 | 68,489 (34,706, 147,118) | 10,154 (4,568, 23,744) | 113 | 133 |
| African Region | 47 | 13,789 (2,920, 58,232) | 3,547 (608, 13,308) | 15 | 16 |
| Region of the Americas | 36 | 9,583 (7,334, 13,538) | 2,658 (1,668, 3,858) | 23 | 30 |
| Eastern Mediterranean Region | 22 | 13,886 (5,216, 25,751) | 1,130 (653, 2,129) | 19 | 21 |
| European Region | 53 | 1,210 (606, 2,022) | 977 (724, 1,321) | 33 | 40 |
| South-East Asia Region | 11 | 28,030 (17,669, 43,597) | 643 (447, 1,079) | 7 | 8 |
| Western Pacific Region | 26 | 1,991 (961, 3,978) | 1,199 (468, 2,049) | 16 | 18 |

### Factors associated with MMR deviations

The explanatory analysis included 31 country observations in 2021 and 32 in 2022. Twenty-four countries contributed observations in both years, while 15 contributed data for one year only, yielding 63 country-year observations from 39 unique countries. In the model containing all seven predictors, the three service disruption coefficients were close to zero with confidence intervals spanning zero, while baseline maternal mortality and 2022 had negative coefficients and skilled birth attendance had a small positive coefficient (Figure 3 and eTable 2).

**Figure 3.**
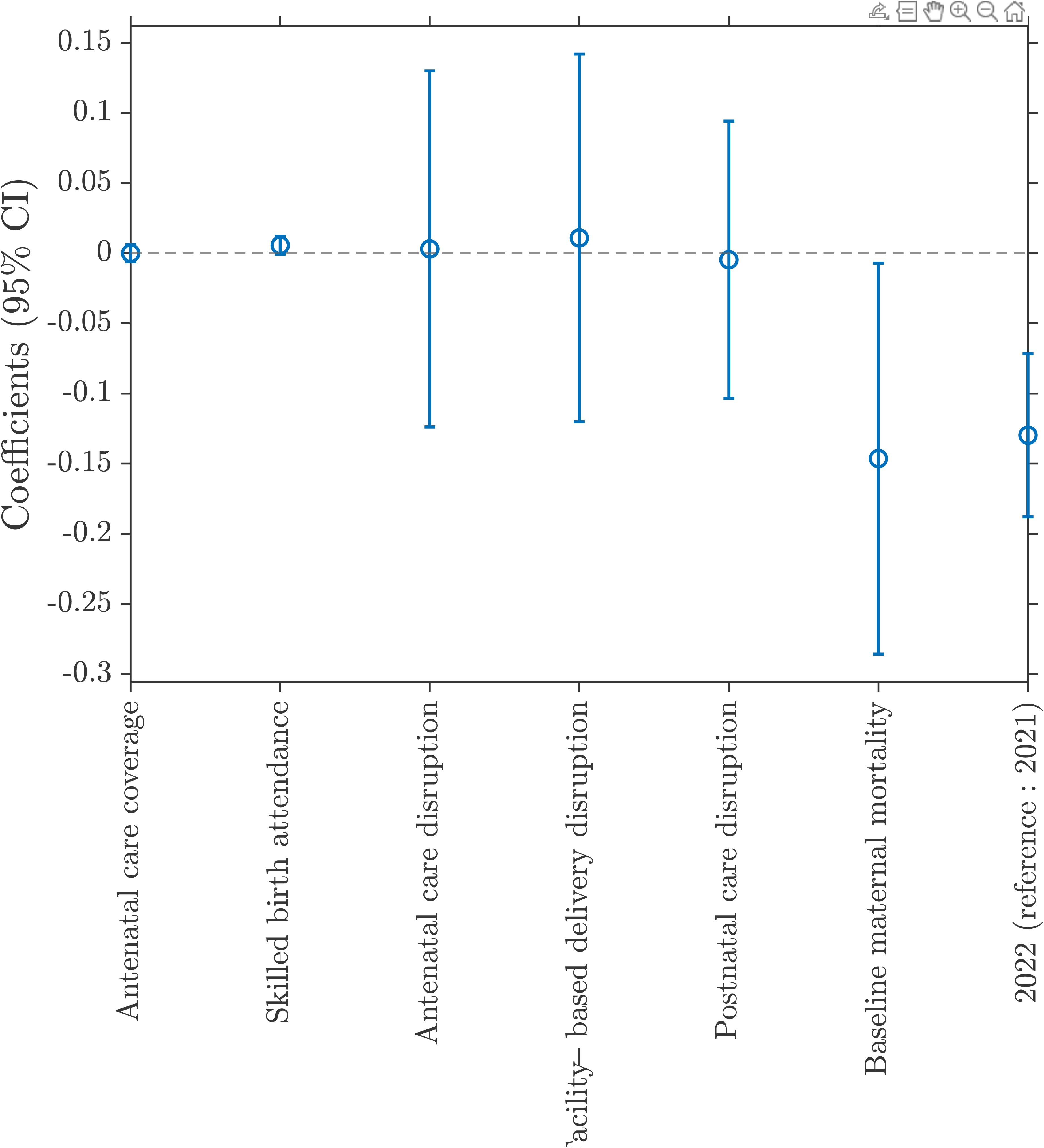
Coefficients and 95% confidence intervals from the regression containing all seven predictors. The outcome was log(MMEIG-estimated MMR/counterfactual MMR). Baseline maternal mortality was log-transformed and 2021 was the reference year. Repeated observations within countries were accounted for using a country-specific random intercept.

Backward selection retained baseline maternal mortality, skilled birth attendance and year (Table 3), with AIC decreasing from −3.01 to −10.89. Higher baseline maternal mortality was associated with a lower observed-to-expected MMR ratio (coefficient −0.15, 95% CI −0.29, - 0.01), and the ratio was lower in 2022 than in 2021 (coefficient −0.13, 95% CI −0.19, −0.07). Skilled birth attendance had a small positive coefficient, but its confidence interval included the null (coefficient 0.0056, 95% CI −0.00001, 0.0112).

**Table 3.**
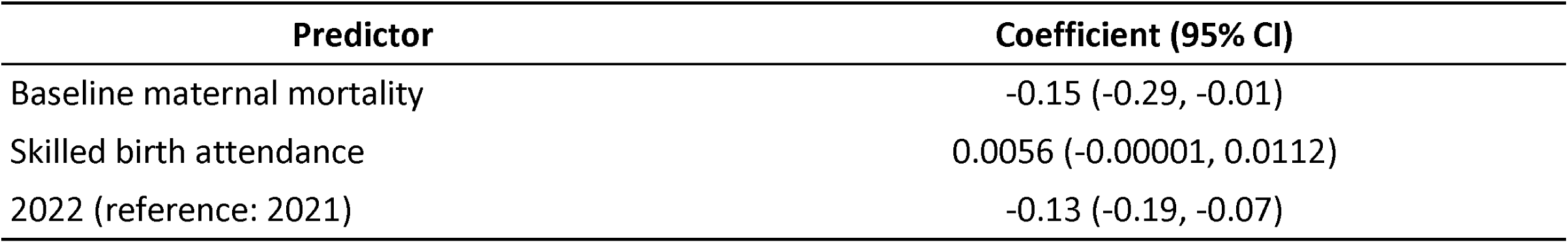
Predictors retained after backward AIC selection. Outcome: log(MMEIG-estimated MMR/counterfactual MMR). Baseline maternal mortality was log-transformed and 2021 was the reference year. AIC was −3.01 with all seven predictors and −10.89 after selection.

| Predictor | Coefficient (95% CI) |
| --- | --- |
| Baseline maternal mortality | -0.15 (-0.29, -0.01) |
| Skilled birth attendance | 0.0056 (-0.00001, 0.0112) |
| 2022 (reference: 2021) | -0.13 (-0.19, -0.07) |

Testing each disruption measure separately gave the same pattern: antenatal care disruption 0.0087 (95% CI −0.0470, 0.0644), facility-based delivery disruption 0.0104 (−0.0499, 0.0707), and postnatal care disruption 0.0060 (−0.0555, 0.0676); all three confidence intervals included zero (eTable 4 and eFigure 2). In the leave-one-country-out analysis, no country produced a large change in the estimated coefficients (eFigure 3). Namibia was examined separately because its 2021 and 2022 observations had the two largest standardised residuals. Repeating the analysis without Namibia did not materially change the coefficients (eTable 5). The backward selection path is shown in eTable 3.

### Comment

#### Principal findings

Across 195 countries and territories, we estimated 68,489 excess maternal deaths during 2020-2023, with statistically detectable cumulative excess in 113 settings; departures from pre-pandemic WHO/UN MMEIG trajectories were highly heterogeneous. In the nested analysis of 39 self-selected countries with aligned predictor data, antenatal care disruption, facility-based delivery disruption and postnatal care disruption were not independently associated with signed MMR deviation, including when each measure was tested separately. Backward selection retained baseline maternal mortality, skilled birth attendance and year, with lower observed-to-expected MMR in 2022 than in 2021; these exploratory ecological associations should not be interpreted causally.

The lack of a detectable association with the three disruption measures should not be interpreted as evidence that maternal service disruption was unimportant. Previous studies documented reductions in antenatal, delivery and postnatal service use during COVID-19 and described adverse maternal and perinatal outcomes or modelled additional maternal deaths under service reduction scenarios.^7–10^ Our analysis asks a different question: whether broad national disruption measures explain differences between countries in annual departure from their own expected MMR trajectories. Multicountry evidence shows that service use changed unevenly across countries and services rather than following a single global pattern.^7,8^ These indicators may not capture timing, duration, quality of care or within-country variation. The three measures were also strongly correlated, although separate models produced the same null pattern. The findings therefore support a multifactorial interpretation rather than a simple relationship between reported service disruption and excess maternal mortality.

Direct clinical effects of SARS-CoV-2 and changes in the pandemic context remain important. COVID-19 infection during pregnancy has been associated with greater risk of severe disease and adverse maternal and neonatal outcomes.^5,6^ The lower observed-to-expected MMR in 2022 may be consistent with health-system adaptation, recovery of services and broader changes in the pandemic, but our analysis cannot distinguish among these mechanisms. Vaccination may also have contributed to changing risk, and guidance for vaccination during pregnancy varied across countries.^11^ Vaccination coverage in pregnancy was not included because comparable country-year data were not consistently available. The inverse association with baseline MMR should not be interpreted as protective because the outcome measures relative departure from each country’s own counterfactual. The small, imprecisely estimated positive coefficient for skilled birth attendance should likewise be interpreted cautiously because country-level associations may reflect residual confounding. An annual national disruption category may also miss short periods of severe strain, recovery within the same year, or protection associated with vaccination. Lesotho also has a high HIV burden, which is relevant because HIV is associated with substantially higher pregnancy-related mortality; this context may contribute to its high baseline risk and unusual trajectory.^27,28^

The absence of excess with a lower 95% UI above zero in some countries also requires caution. WHO/UN MMEIG values are model-based estimates, and maternal mortality measurement remains affected by incomplete civil registration, underreporting and misclassification.^16,29,30^ Pandemic disruption may have further affected registration systems. Wide uncertainty intervals in some settings likely reflect uncertainty in both the underlying data and the fitted pre-pandemic trajectory. Maternal deaths and MMR also provide different information because MMR is relative to live births and can change with fertility. The signed log ratio used in the explanatory analysis complements the positive-only excess estimates by retaining deviations both above and below the counterfactual baseline.

### Strengths of the study

Strengths of this study include the use of a consistent counterfactual framework across 195 countries and territories, evaluation of both maternal deaths and MMR, and accounting for pre-pandemic nonlinear trends rather than relying on simple before and after comparisons. The explanatory analysis also included model selection and influence checks. No single country materially changed the estimated coefficients, and excluding the two Namibia observations with the largest standardised residuals did not alter the conclusions.

### Limitations of the data

Several limitations remain. The primary analysis compares WHO/UN MMEIG model-based pandemic-period estimates with counterfactual forecasts calibrated to pre-pandemic model-based estimates rather than raw surveillance counts, and MMEIG uncertainty may not be fully propagated into the reported intervals. The counterfactual assumes that pre-pandemic trends would have continued, which may be less plausible where conflict, climate shocks, health-system reforms, funding changes or other events occurred. The explanatory analysis was limited to 63 country-year observations from the 39 countries that responded to the WHO Global Pulse Survey and could be matched to the other predictors; because inclusion depended on survey response, the sample is self-selected and should not be treated as globally representative. Its predictors are ecological country-level measures, the disruption indicators are relatively coarse, and the analysis cannot separate deaths directly related to SARS-CoV-2 from deaths associated with delayed care, service disruption or other shocks. These limitations restrict causal interpretation.

### Interpretation

From a public health perspective, the absence of a national-level association argues for better measurement of disruption, not for deprioritising continuity of care. Emergency preparedness should protect antenatal, intrapartum, emergency obstetric, referral, and postnatal services while pairing service-availability measures with time-resolved indicators of quality, staffing, referral delay, and pregnancy-specific vaccination. Strengthening civil registration, maternal death notification, confidential enquiry, and maternal and perinatal death surveillance and response would improve both prevention and interpretation of future mortality signals.^29,30,33^ Risk communication and community engagement are also important for addressing fear, misinformation, stigma and delays in care seeking, particularly in rural and underserved populations.^31,32^ Clear and equitable vaccination guidance during pregnancy should also be considered when vaccines are part of future emergency response.^11^

### Conclusions

Maternal mortality departed substantially but unevenly from pre-pandemic trajectories during 2020-2023. In a self-selected subset of 39 countries, broad national disruption measures were not clearly associated with signed MMR deviation; this should not be interpreted as evidence that service disruption was unimportant. The findings indicate that pandemic-related excess maternal mortality cannot be attributed to a single measured pathway and underscore the need to protect the full continuum of maternal care while strengthening timely measurement of service access and quality.

## Declarations

### Funding

This study was partially supported by NSF ACED #2435886. The funder had no role in the study design, analysis, interpretation, manuscript preparation, or decision to submit.

### Conflicts of interest

The authors declare no conflicts of interest.

Not applicable. This study used publicly available aggregated country-level data and was not based on clinical or patient data.

Consent to participate. Not applicable.

Consent for publication. Not applicable.

Data availability statement Data are publicly available from the WHO/UN MMEIG, WHO Global Pulse Survey, and UNICEF sources cited in the manuscript. Derived analytic data are available at https://github.com/hkarami-GSU/Maternal_Mortality.

Code availability. Analysis code is available at https://github.com/hkarami-GSU/Maternal_Mortality.

### Authors’ contributions

JHG, HK, and GC conceived and designed the study. HK and JHG curated and analysed the data. HK implemented the modelling and prepared the tables and figures.

JHG, HK, and GC drafted the manuscript. HAH, MAD, GMM, and GC contributed to interpretation and critical revision. GC supervised the study. All authors reviewed and approved the final manuscript.

AI-use disclosure: Generative AI tools were used to assist with language editing and code cleanup. The authors reviewed, verified, and take full responsibility for all manuscript content.

## Data Availability

Input data are publicly available from the WHO/UN MMEIG maternal mortality estimates. Derived outputs supporting the main tables and figures are provided in the Supplementary Material, including Supplementary Sections S1-S8.

https://github.com/hkarami-GSU/Maternal_Mortality

## Notes

### Competing Interest Statement

The authors have declared no competing interest.

### Summary of Updates

The manuscript has been substantially revised. The title, abstract, methods, results, discussion, figures, tables, and supplementary material have been updated. We added an exploratory country-level analysis examining associations between maternal health service disruptions and deviations from expected maternal mortality, including sensitivity and influence analyses. The presentation and interpretation of the counterfactual estimates were also clarified, and the supplementary analyses and figures were expanded and updated.

