## Supplementary Material for "Global Excess Maternal Mortality During the COVID-19 Pandemic: Country-Level Counterfactual Estimates and Associated Factors"

### 1 eMethods

#### 1.1 Counterfactual forecasts

The counterfactual analysis was carried out separately for maternal deaths and maternal mortality ratio (MMR). For each country, WHO/UN MMEIG estimates from 2000 to 2019 were used for model fitting. The fitted model was then projected to 2020-2023. These projections describe the values expected if the pre-pandemic trajectory had continued.

Each trajectory was written as the sum of one or two generalised logistic components. For component $i$,

$$\frac{dC_i(t)}{dt} = A_i(t) r_i C_i(t)^{p_i} \left(1 - \frac{C_i(t)}{K_i}\right),$$

where $r_i$ controls the growth rate, $p_i$ controls the shape, and $K_i$ is the saturation level. The first component is active at the start. A later component becomes active after the previous component passes a threshold $C_{\text{thr}}$,

$$A_i(t) = \begin{cases} 1, & C_{i-1}(t) > C_{\text{thr}}, \\ 0, & \text{otherwise.} \end{cases}$$

Candidate thresholds were taken from a grid based on the cumulative fitted series. One-component and two-component models were considered. Parameters were estimated by nonlinear least squares with 30 starting points. Models were ranked using the corrected Akaike Information Criterion,

$$\text{AICc} = -2\log(L) + 2m + \frac{2m(m+1)}{n_d - m - 1},$$

where $m$ is the number of model parameters and $n_d$ is the number of observations used for fitting.

Uncertainty was estimated with 300 bootstrap samples. Normal and Poisson error structures were considered. The final forecast used the setting with better calibration-period coverage. The same steps were used for maternal deaths and MMR.

#### 1.2 Excess maternal deaths and excess MMR

For year $t$, excess maternal deaths were calculated as

$$E_t = \max(0, O_t - \widehat{O}_t),$$

where $O_t$ is the WHO/UN MMEIG estimate and $\widehat{O}_t$ is the median counterfactual forecast. The lower and upper bounds were calculated from the upper and lower forecast bounds,

$$E_t^L = \max(0, O_t - U_t), \qquad E_t^U = \max(0, O_t - L_t).$$

Annual excess values were summed over 2020-2023 to describe cumulative burden. The same calculation was applied to MMR. The summed MMR values describe cumulative annual deviation and are not a single-year MMR.

The primary excess measure was restricted to positive deviations because the aim was to measure additional burden above the counterfactual trajectory. A value below the counterfactual may reflect continued improvement, reporting differences, uncertainty in the source estimates, or uncertainty in the forecast. The explanatory analysis retained the direction of departure by using the signed log ratio described below.

##### 1.3 Regression dataset and predictors

The explanatory analysis used complete records and did not impute missing predictor values. The final dataset contained 63 country-year observations from 39 countries: 31 observations in 2021 and 32 in 2022, with 24 countries contributing both years. The outcome was

$$Y_{ct} = \log\left(\frac{\text{MMR}_{ct}^{\text{WHO/UN MMEIG}}}{\text{MMR}_{ct}^{\text{counterfactual}}}\right).$$

A value of zero means that the WHO/UN MMEIG MMR equalled the counterfactual value, a positive value means it was above the counterfactual, and a negative value means it was below the counterfactual.

Seven predictors were considered: antenatal care disruption, facility-based delivery disruption, postnatal care disruption, baseline MMR, antenatal care coverage, skilled birth attendance, and year. The three disruption variables came from the WHO Global Pulse Survey on Continuity of Essential Health Services. They were coded as 0 for an increase of at least 5% or a decrease below 5%, 1 for a 5-25% decrease, 2 for a 26-50% decrease, and 3 for a decrease above 50%. Baseline MMR was the 2019 WHO/UN MMEIG estimate and was log transformed. Antenatal care coverage was the latest available pre-2020 percentage of women receiving at least four antenatal visits. Skilled birth attendance was the latest available pre-2020 percentage of births attended by skilled health personnel. Year was treated as a category, with 2021 as the reference year.

##### 1.4 Regression analysis

A linear mixed-effects regression with a country-specific random intercept was used so that repeated observations from the same country were not treated as independent. The initial model included all seven predictors. Pairwise correlations and variance inflation factors were checked before model selection. All variance inflation factors were below 10.

A backward AIC procedure was used to obtain a simpler model. Year and the country-specific intercept were kept in every model. At each step, each remaining predictor was removed one at a time. A removal was kept only when it reduced AIC. The procedure stopped when no further removal reduced AIC.

##### 1.5 Sensitivity analyses

Three additional models tested antenatal care disruption, facility-based delivery disruption, and postnatal care disruption separately. Each model also included baseline MMR, antenatal care coverage, skilled birth attendance, year, and the country-specific intercept.

Observation-level residuals were reviewed to identify unusual observations. Two observations from Namibia, one in 2021 and one in 2022, had absolute Pearson residuals above 2; none exceeded 3. The selected model was refitted after removing Namibia. The model was also refitted after removing each country one at a time. For each removal, the largest change in a coefficient was divided by the standard error of that coefficient in the model containing all seven predictors.

#### 2 Supplementary tables and figures

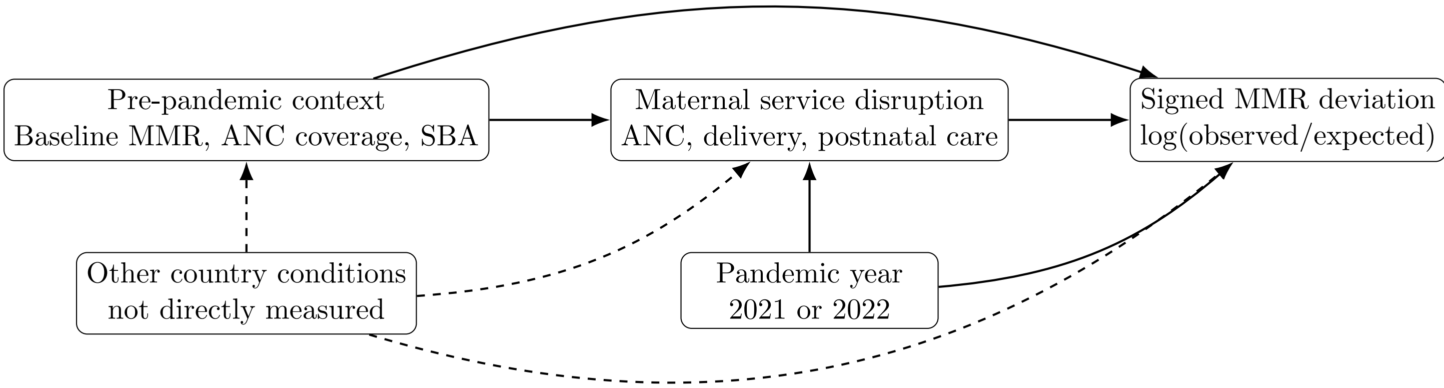


eFigure 1: Conceptual diagram for the explanatory analysis. Arrows show the temporal ordering used to select variables. The analysis was associational and was not designed to identify causal effects.

eTable 1: Estimated annual and cumulative excess maternal deaths from 2020 to 2023 in eight illustrative high-burden countries. Values are median estimates with 95% uncertainty intervals.

| Country | 2020 | 2021 | 2022 | 2023 | Total |
|---|---|---|---|---|---|
| Afghanistan | 719 (199, 1227) | 1469 (949, 1996) | 147 (0, 665) | 0 (0, 463) | 2335 (1148, 4350) |
| Chad | 0 (0, 0) | 0 (0, 185) | 0 (0, 0) | 0 (0, 0) | 0 (0, 185) |
| Guinea-Bissau | 30 (0, 74) | 53 (0, 101) | 14 (0, 71) | 4 (0, 70) | 101 (0, 316) |
| Lesotho | 18 (0, 85) | 40 (0, 109) | 0 (0, 50) | 0 (0, 51) | 58 (0, 295) |
| Liberia | 27 (0, 167) | 74 (0, 233) | 63 (0, 258) | 101 (0, 332) | 265 (0, 990) |
| Nigeria | 0 (0, 0) | 0 (0, 0) | 0 (0, 0) | 0 (0, 0) | 0 (0, 0) |
| Somalia | 227 (0, 613) | 1004 (609, 1410) | 442 (30, 832) | 142 (0, 545) | 1815 (639, 3401) |
| South Sudan | 0 (0, 833) | 0 (0, 1110) | 0 (0, 1009) | 0 (0, 1110) | 0 (0, 4062) |

eTable 2: Regression containing all seven predictors.

| Predictor | Coefficient | 95% CI |
|---|---|---|
| Antenatal care coverage | -0.0001 | -0.0061, 0.0060 |
| Skilled birth attendance | 0.0056 | -0.0007, 0.0119 |
| Antenatal care disruption | 0.0030 | -0.1238, 0.1299 |
| Facility-based delivery disruption | 0.0109 | -0.1202, 0.1419 |
| Postnatal care disruption | -0.0047 | -0.1035, 0.0941 |
| Baseline maternal mortality | -0.1464 | -0.2857, -0.0071 |
| 2022 (reference: 2021) | -0.1297 | -0.1878, -0.0717 |

eTable 3: Backward AIC selection path. The named predictor was removed at each step.

| Step | Predictor removed | AIC |
|---|---|---|
| 0 | None | -3.011 |
| 1 | Antenatal care coverage | -5.011 |
| 2 | Antenatal care disruption | -7.008 |
| 3 | Postnatal care disruption | -9.001 |
| 4 | Facility-based delivery disruption | -10.887 |

eTable 4: Single-disruption sensitivity models.

| Predictor | Coefficient | 95% CI |
|---|---|---|
| Antenatal care disruption | 0.0087 | -0.0470, 0.0644 |
| Facility-based delivery disruption | 0.0104 | -0.0499, 0.0707 |
| Postnatal care disruption | 0.0060 | -0.0555, 0.0676 |

eTable 5: Selected model before and after excluding Namibia.

| Predictor | Primary analysis | Without Namibia |
|---|---|---|
| Baseline maternal mortality | -0.1480 (-0.2866, -0.0094) | -0.1486 (-0.2888, -0.0084) |
| Skilled birth attendance | 0.0056 (-0.0000, 0.0112) | 0.0056 (-0.0001, 0.0113) |
| 2022 (reference: 2021) | -0.1313 (-0.1884, -0.0742) | -0.1176 (-0.1641, -0.0711) |

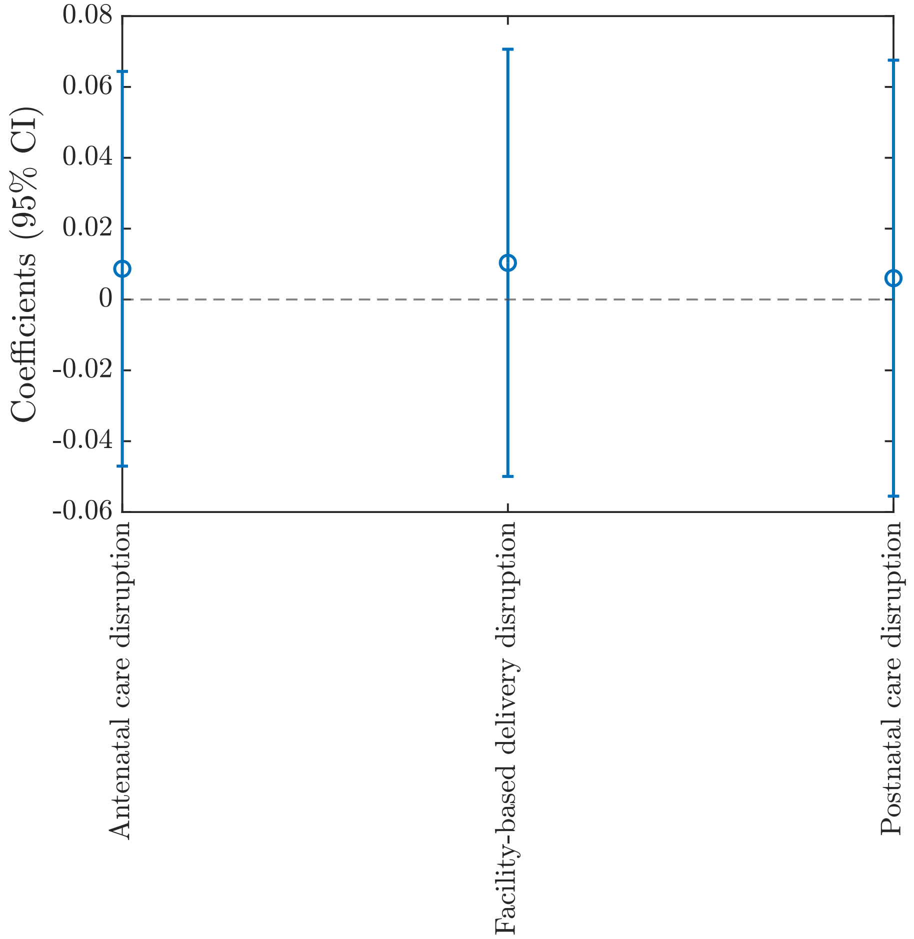


eFigure 2: Coefficients and 95% confidence intervals from the three single-disruption sensitivity models.

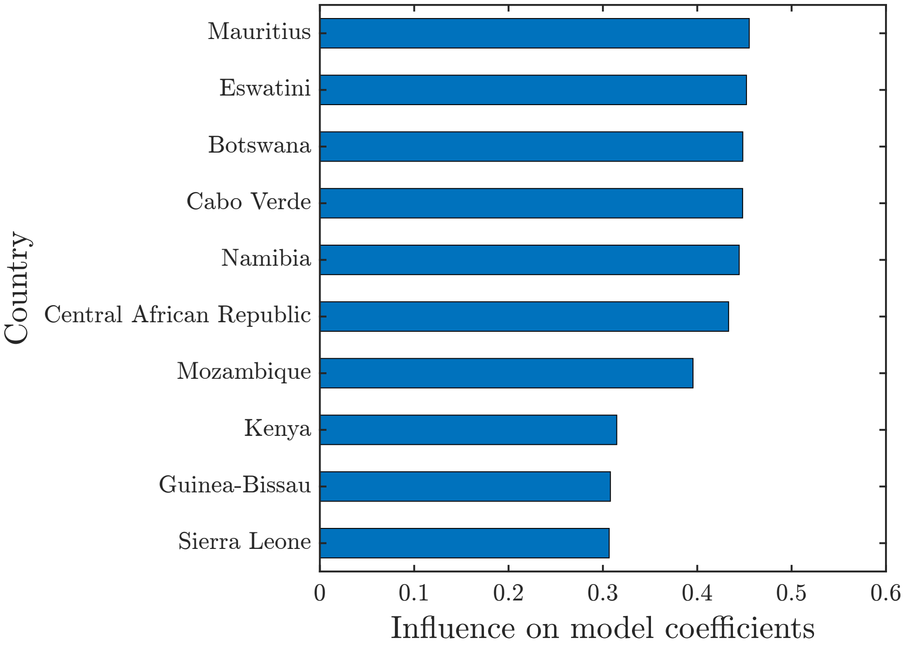


eFigure 3: Ten countries with the largest coefficient changes in the leave-one-country-out analysis. Influence was defined as the largest coefficient change after removing a country, divided by the standard error of that coefficient in the model containing all seven predictors.

### 3 Regional and country-level figures

#### 3.1 South-East Asia Region

##### 3.1.1 Maternal deaths

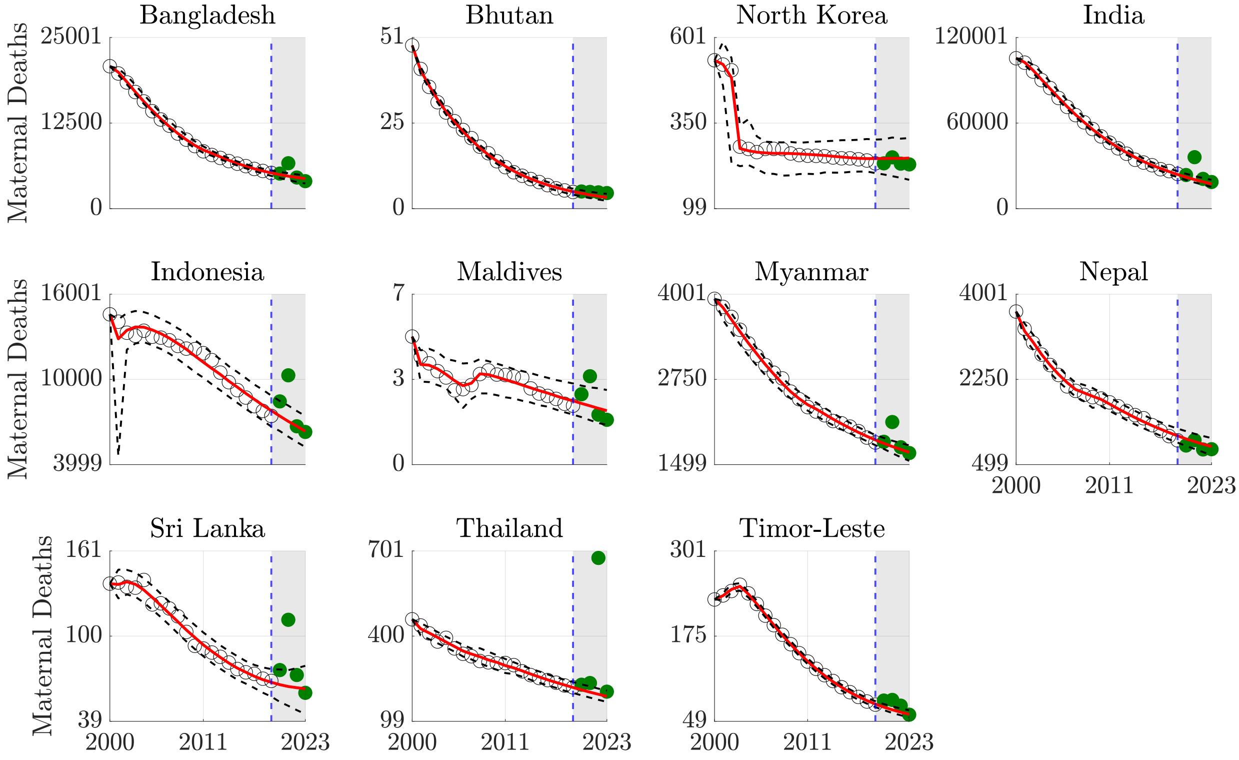


eFigure 4: Counterfactual maternal death forecasts, Southeast Asia Region.

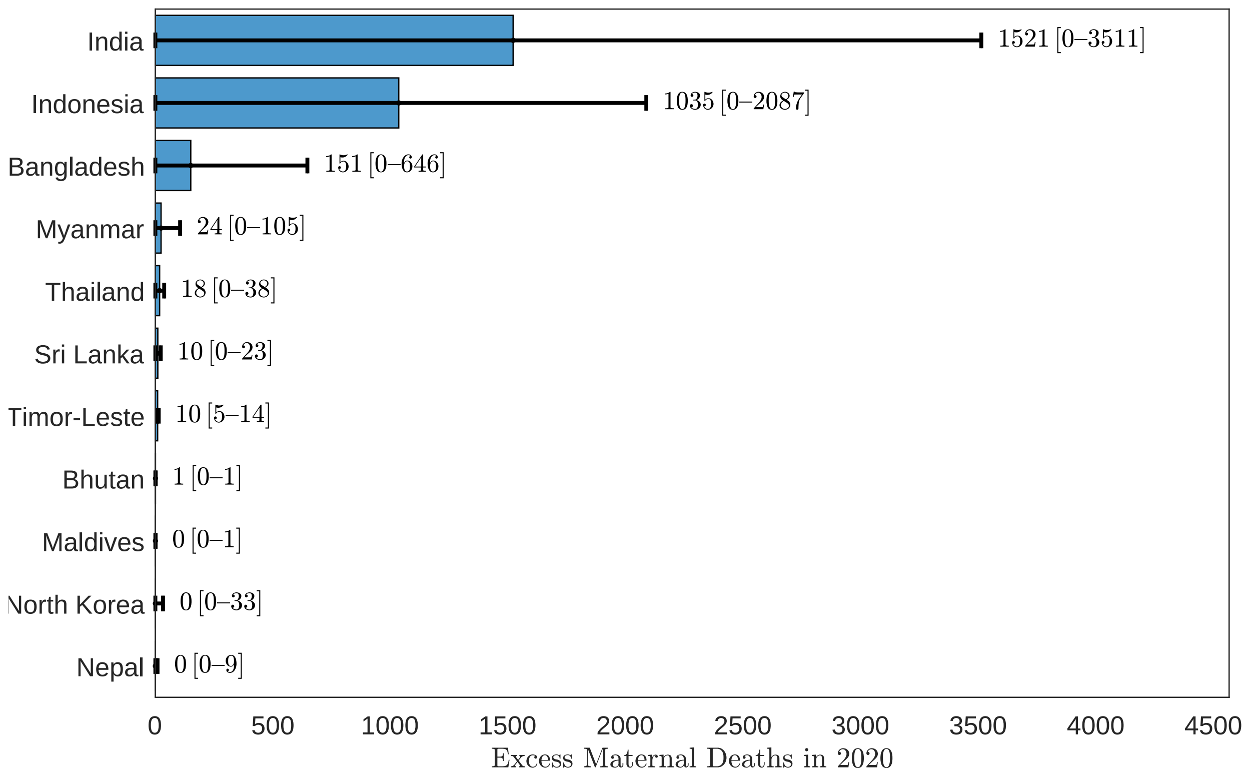


eFigure 5: Excess maternal deaths, Southeast Asia Region, 2020.

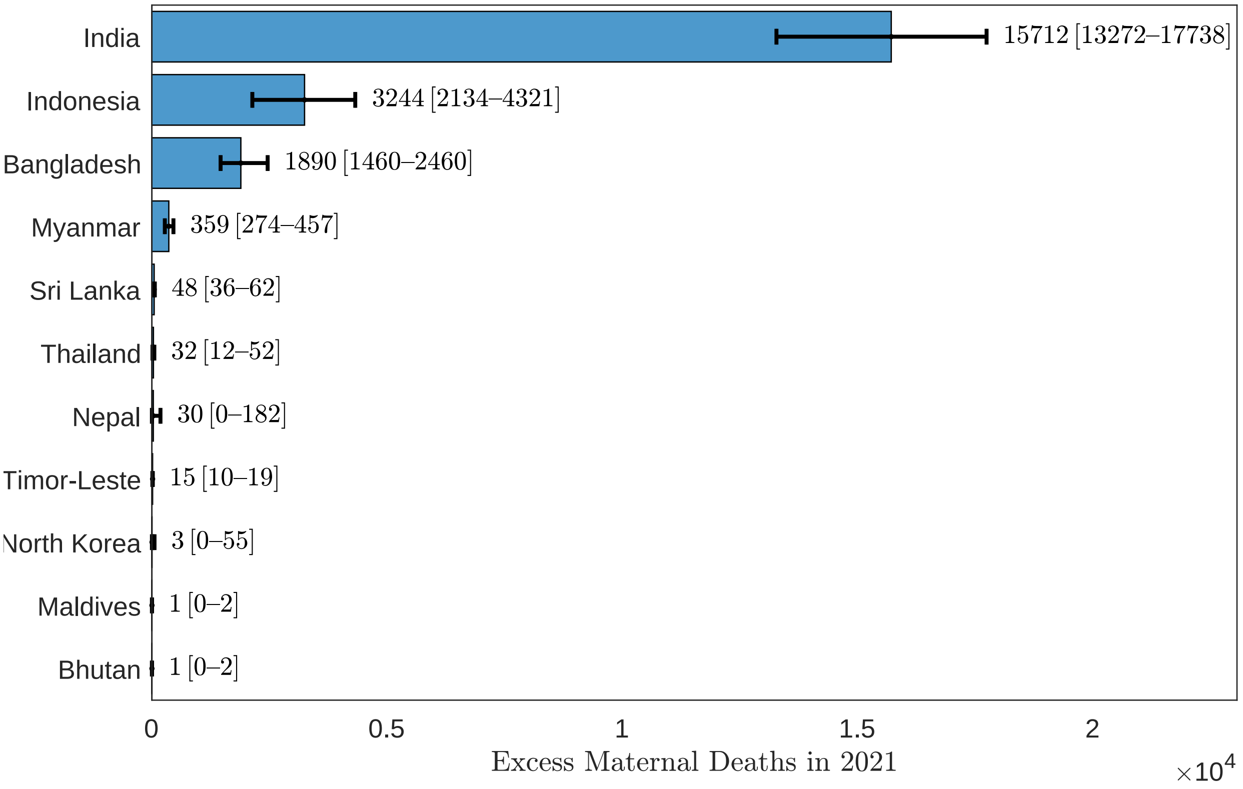


eFigure 6: Excess maternal deaths, Southeast Asia Region, 2021.

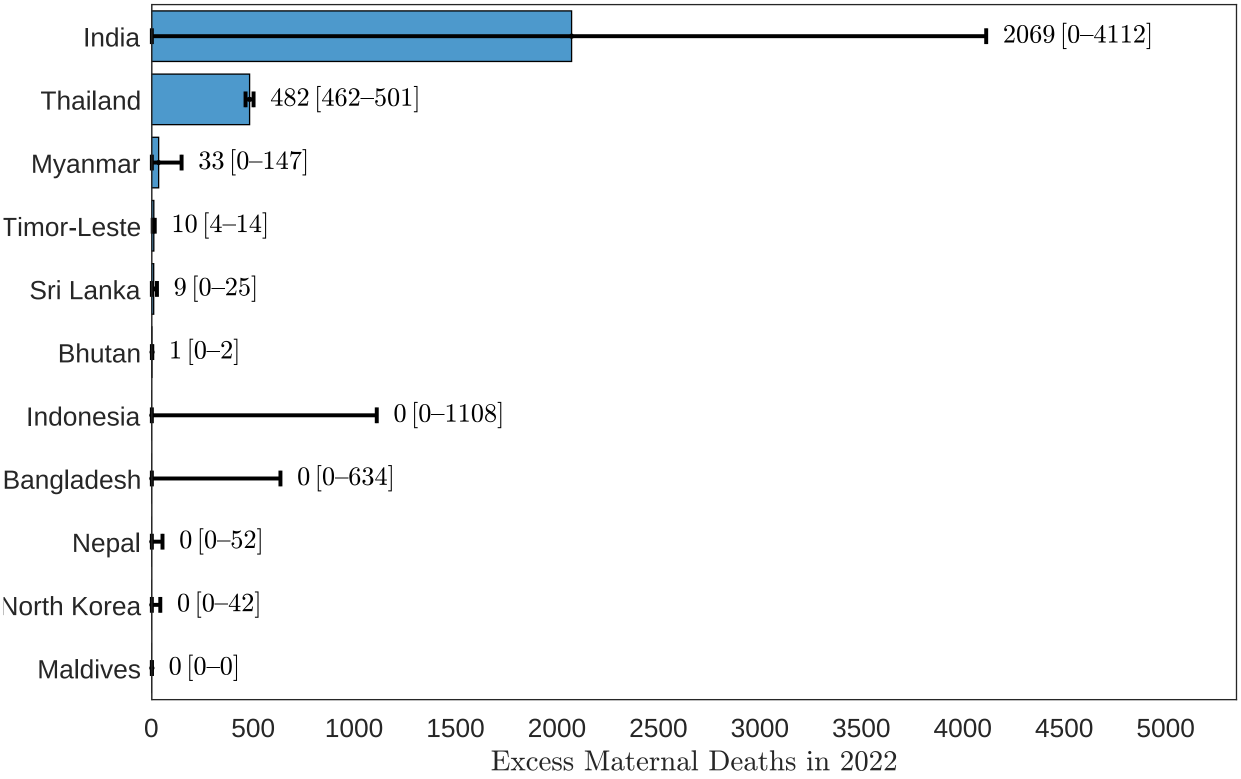


eFigure 7: Excess maternal deaths, Southeast Asia Region, 2022.

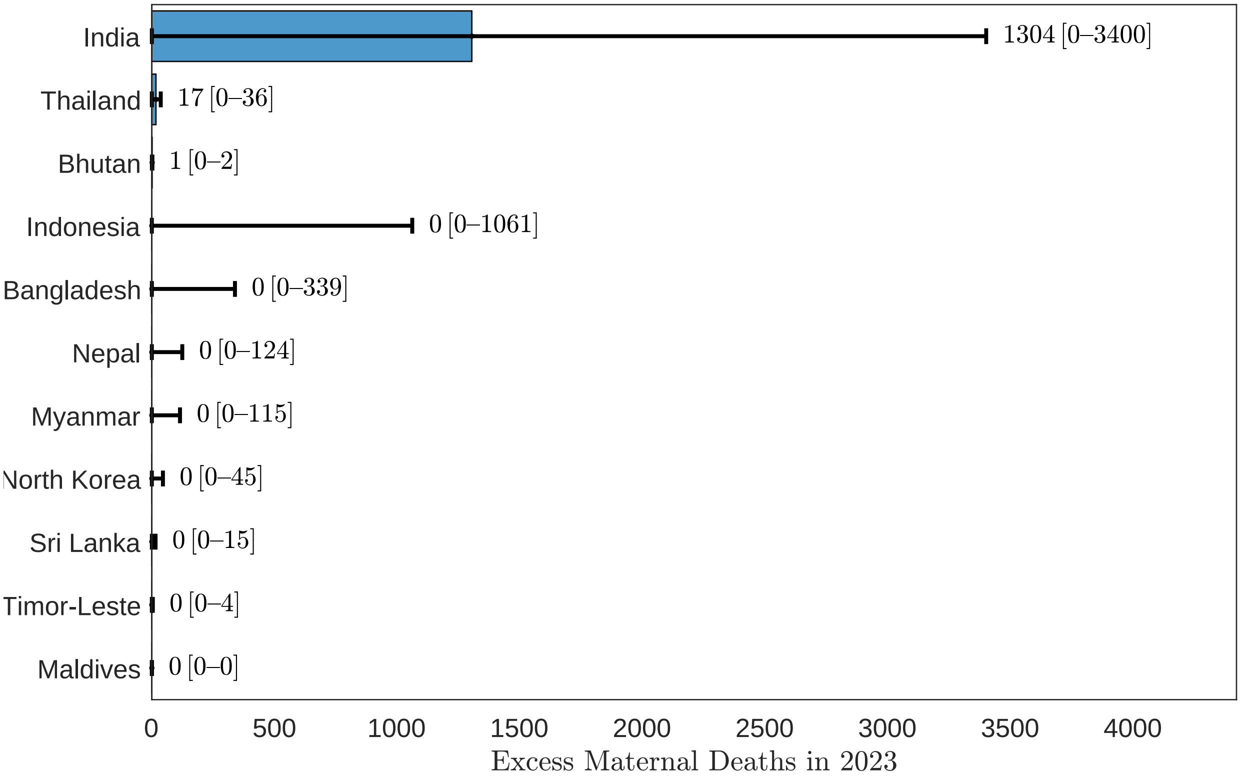


eFigure 8: Excess maternal deaths, Southeast Asia Region, 2023.

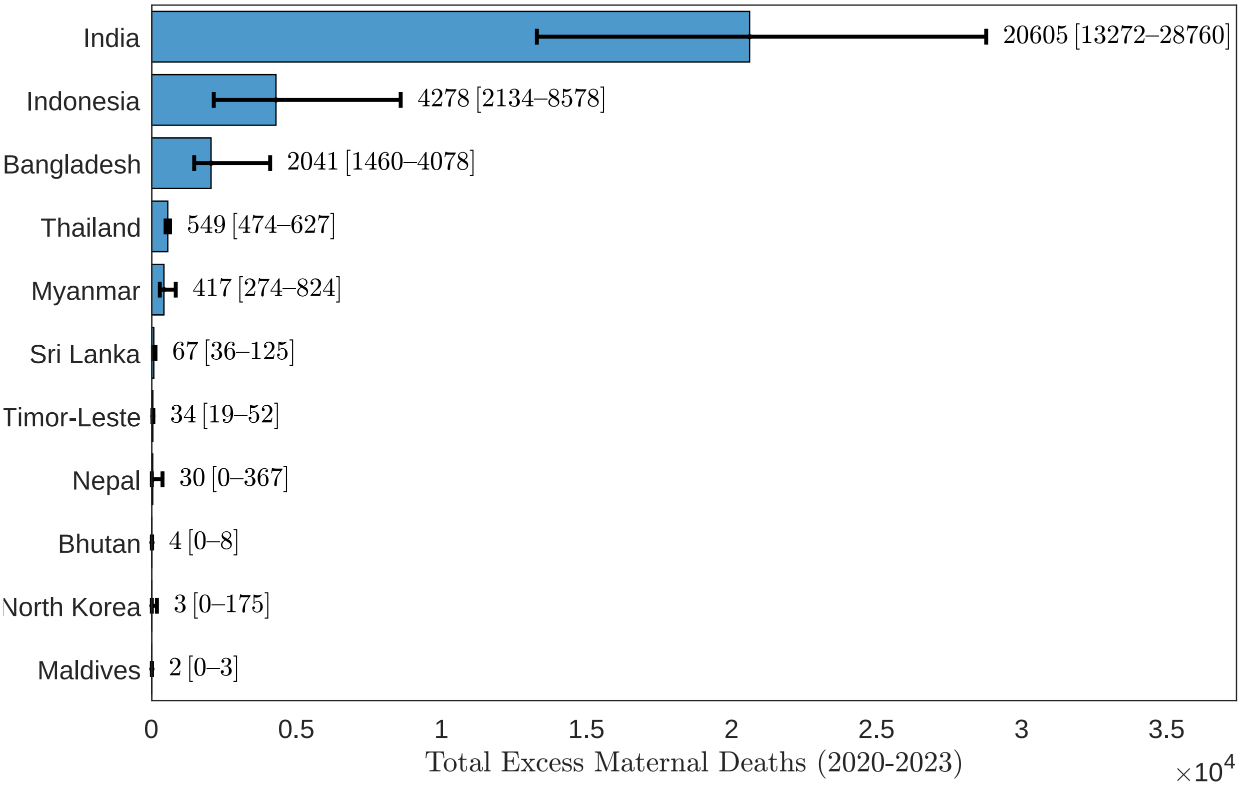


eFigure 9: Excess maternal deaths, Southeast Asia Region, 2020-2023.

##### 3.1.2 MMR

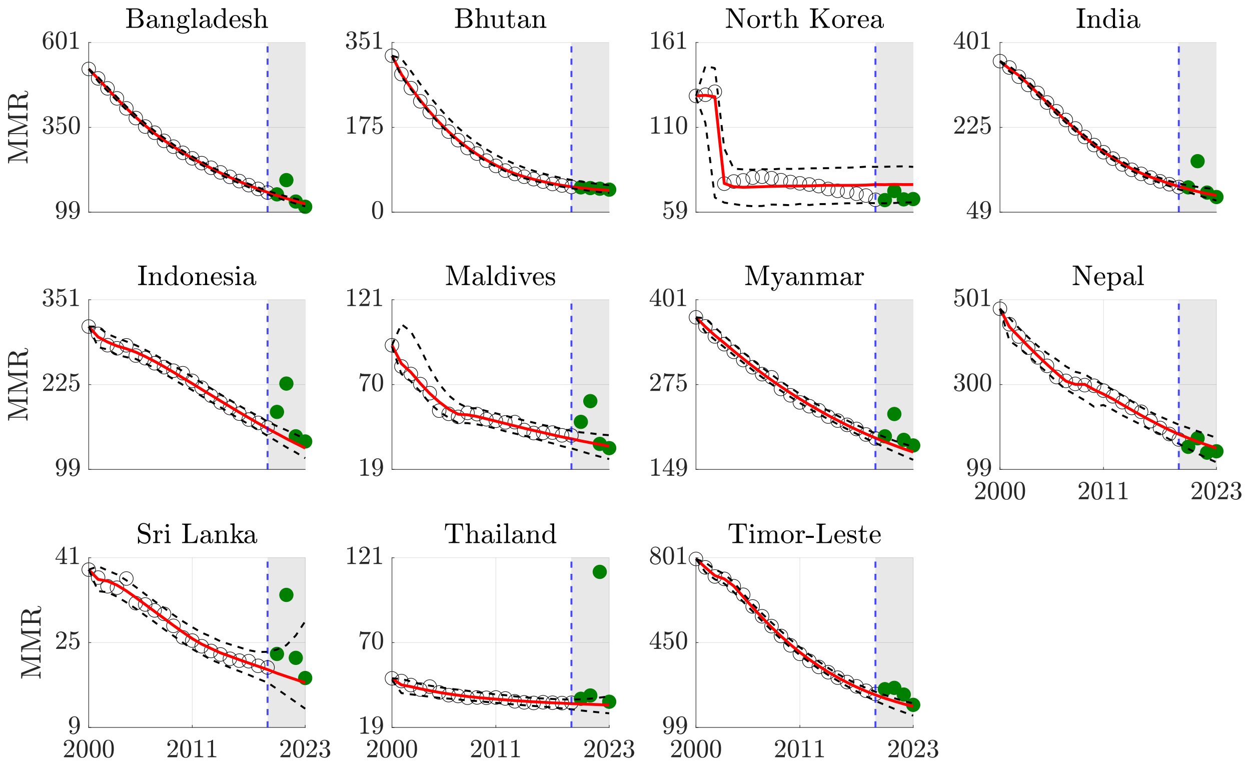


eFigure 10: Counterfactual maternal mortality ratio forecasts, Southeast Asia Region.

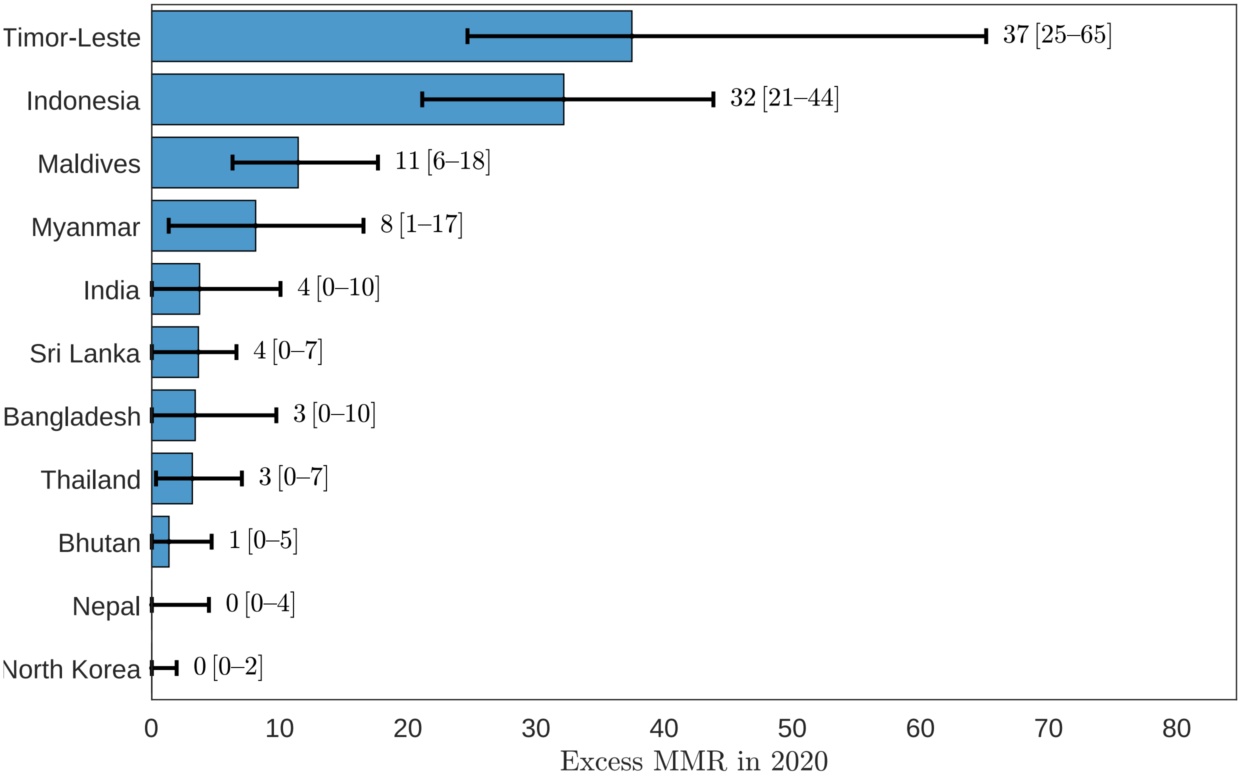


eFigure 11: Excess maternal mortality ratio, Southeast Asia Region, 2020.

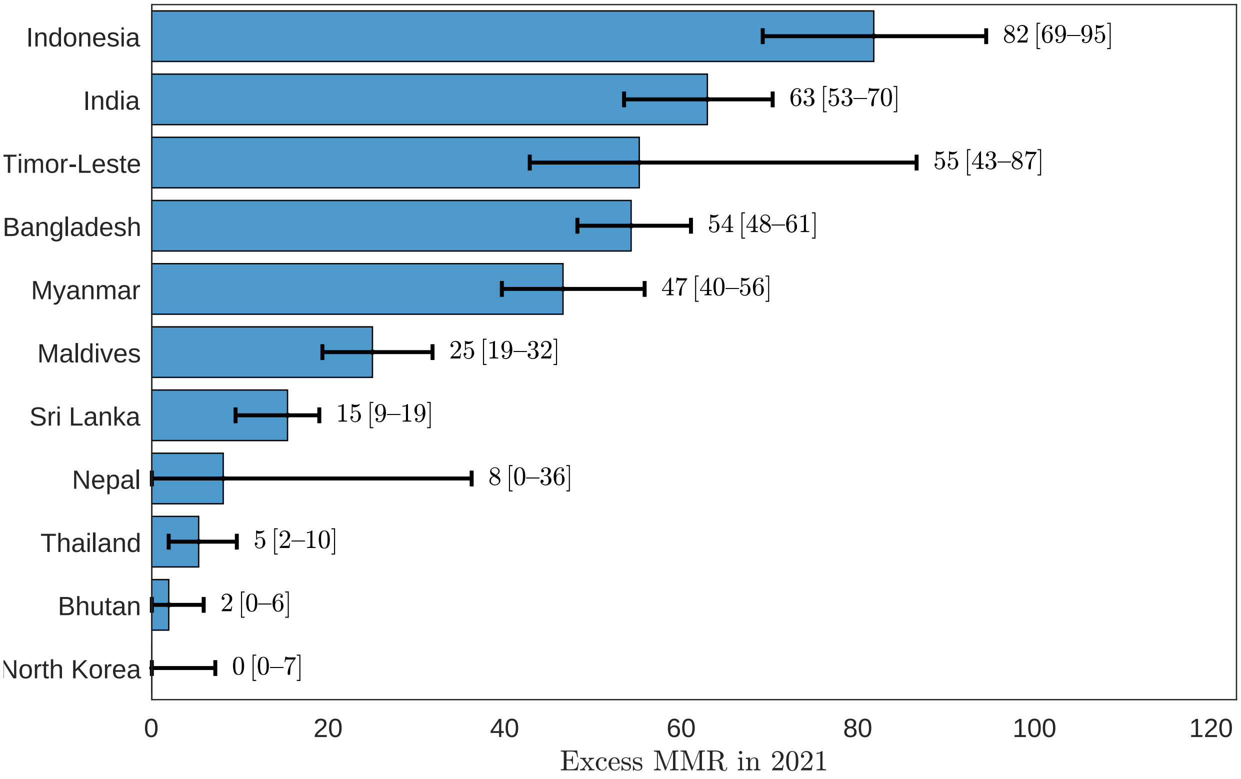


eFigure 12: Excess maternal mortality ratio, Southeast Asia Region, 2021.

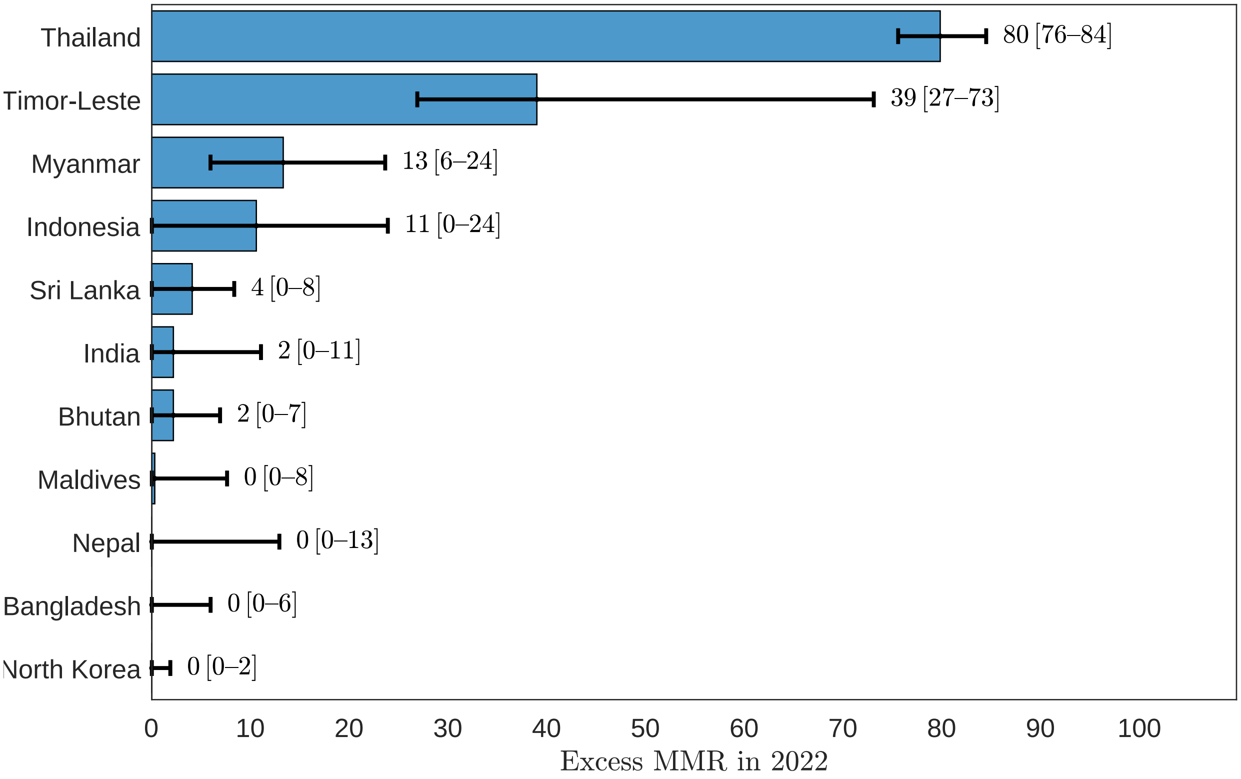


eFigure 13: Excess maternal mortality ratio, Southeast Asia Region, 2022.

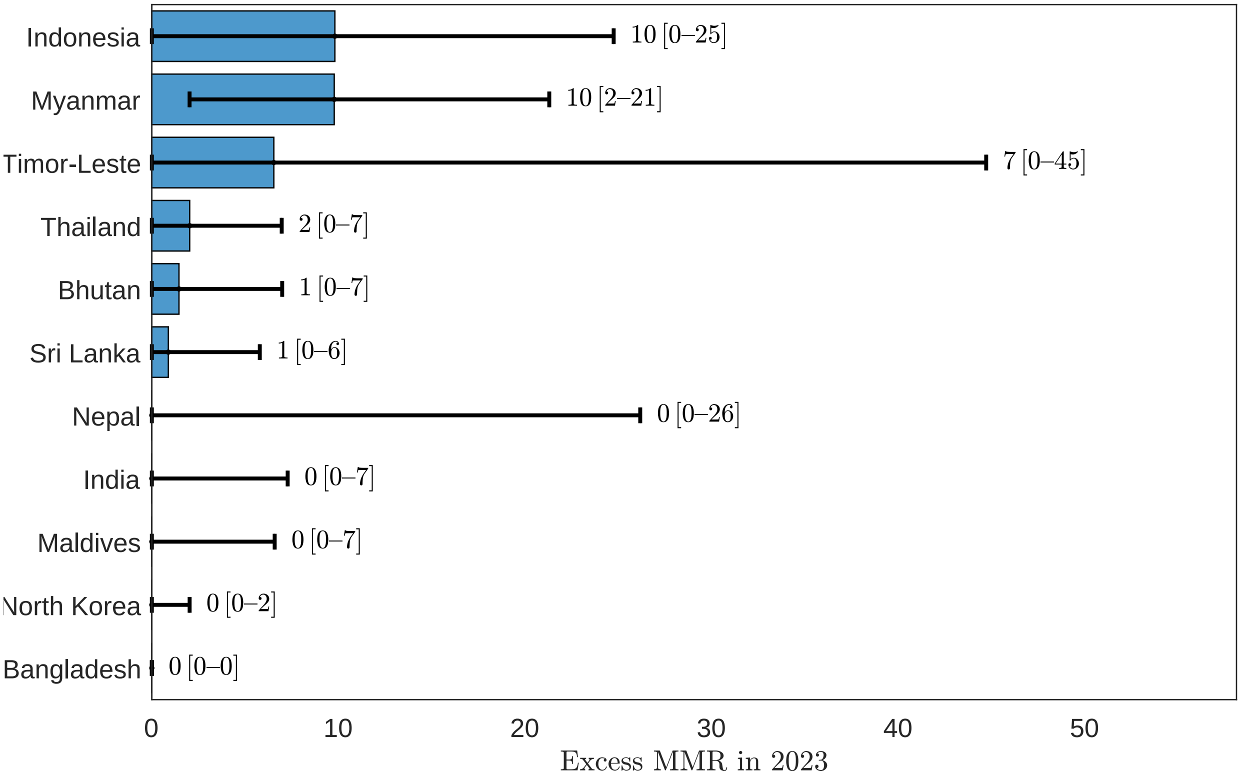


eFigure 14: Excess maternal mortality ratio, Southeast Asia Region, 2023.

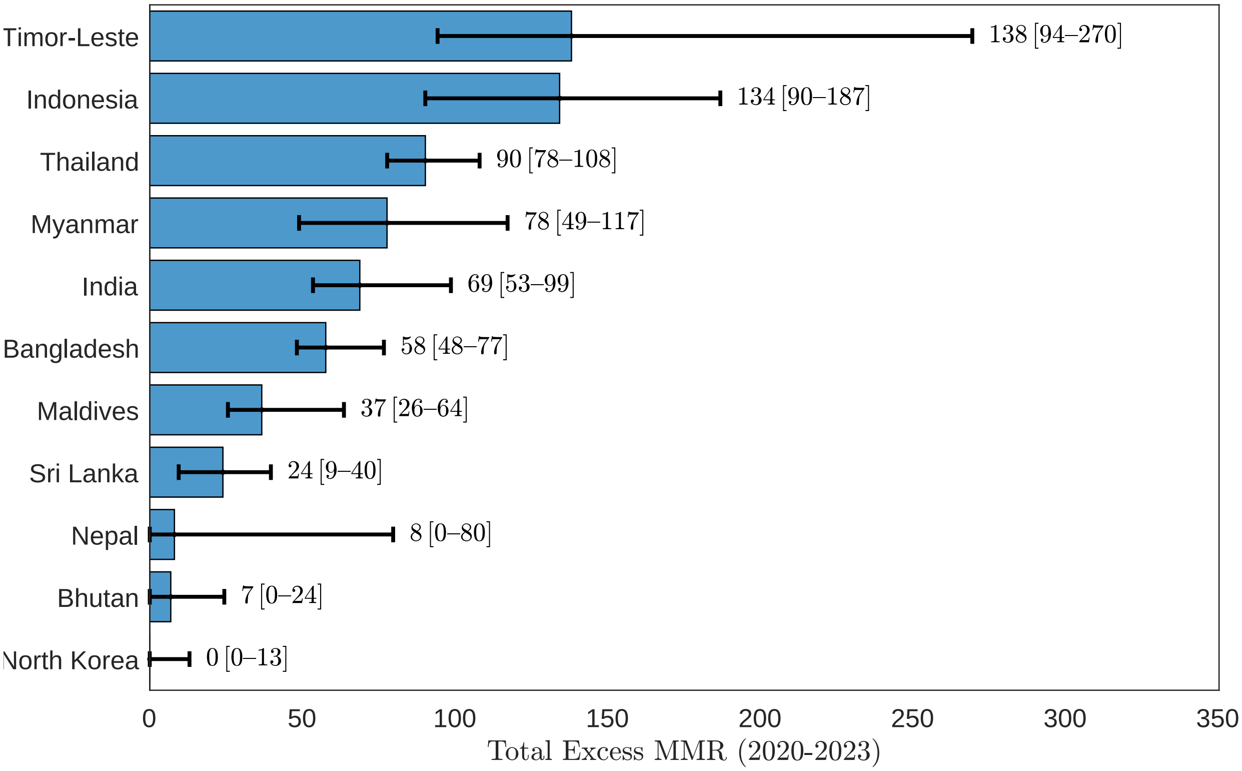


eFigure 15: Excess maternal mortality ratio, Southeast Asia Region, 2020-2023.

#### 3.2 European Region

##### 3.2.1 Maternal deaths

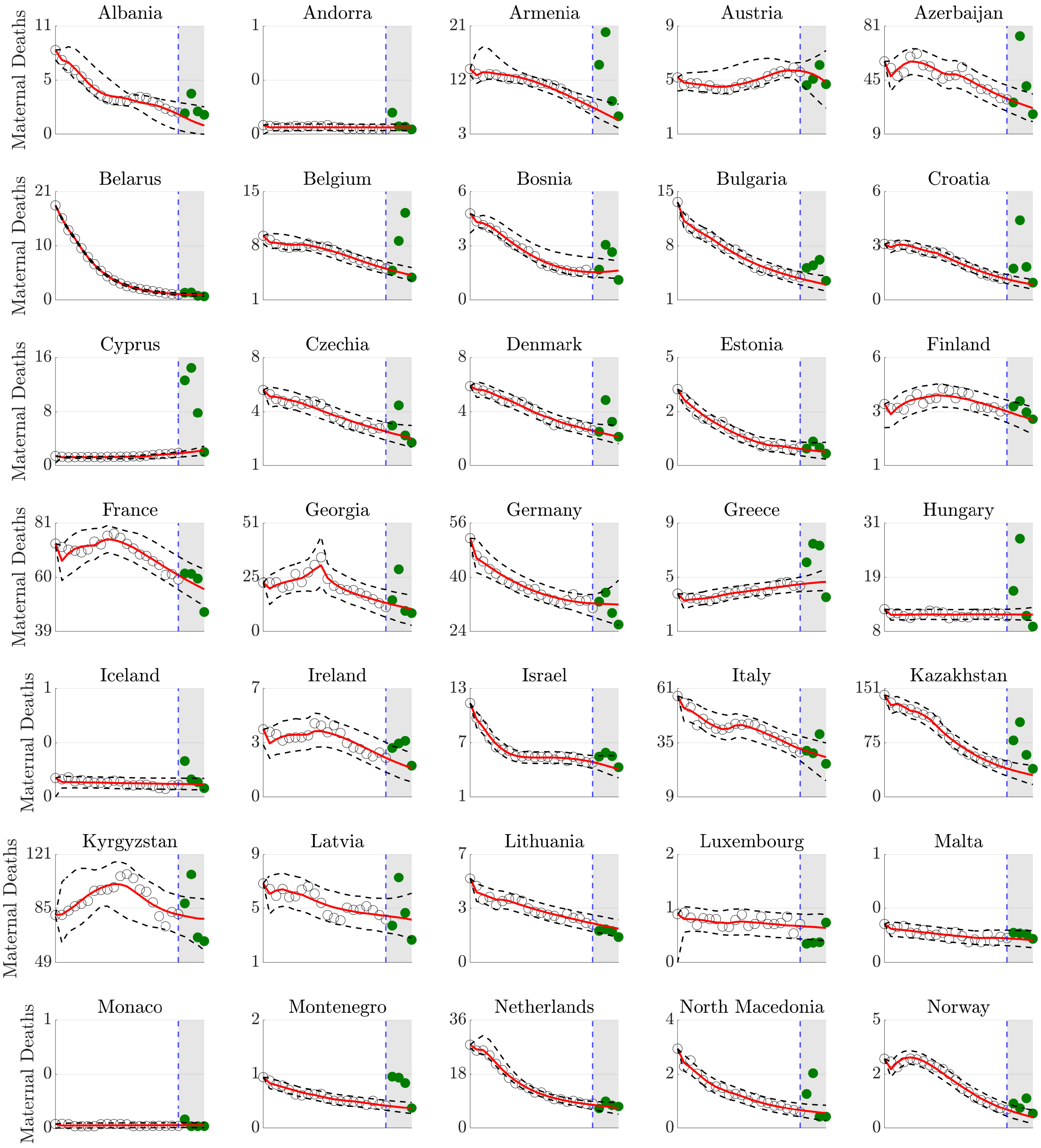


eFigure 16: Counterfactual maternal death forecasts, European Region.

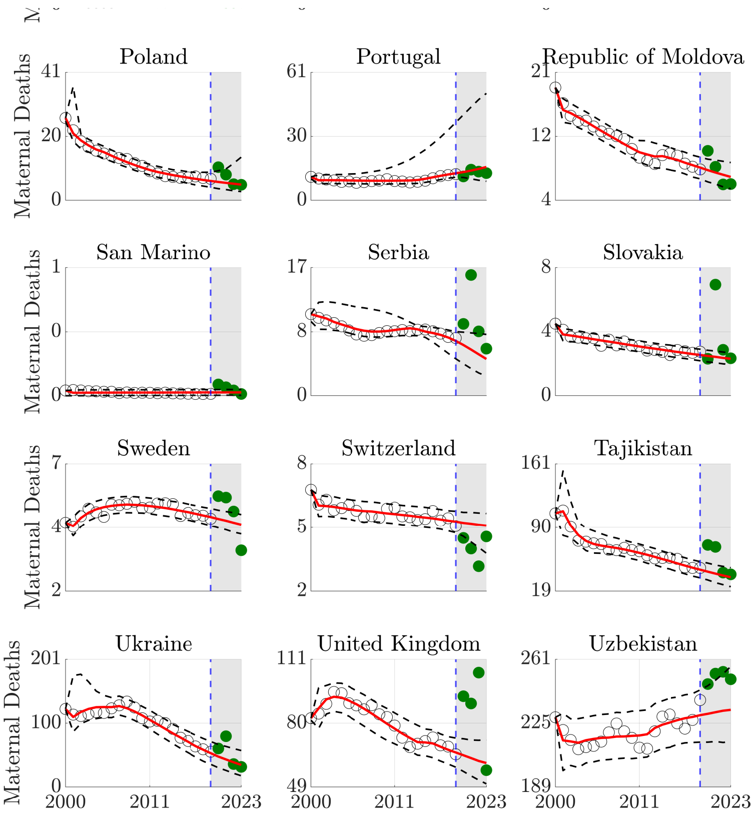


Counterfactual maternal death forecasts, European Region (continued).

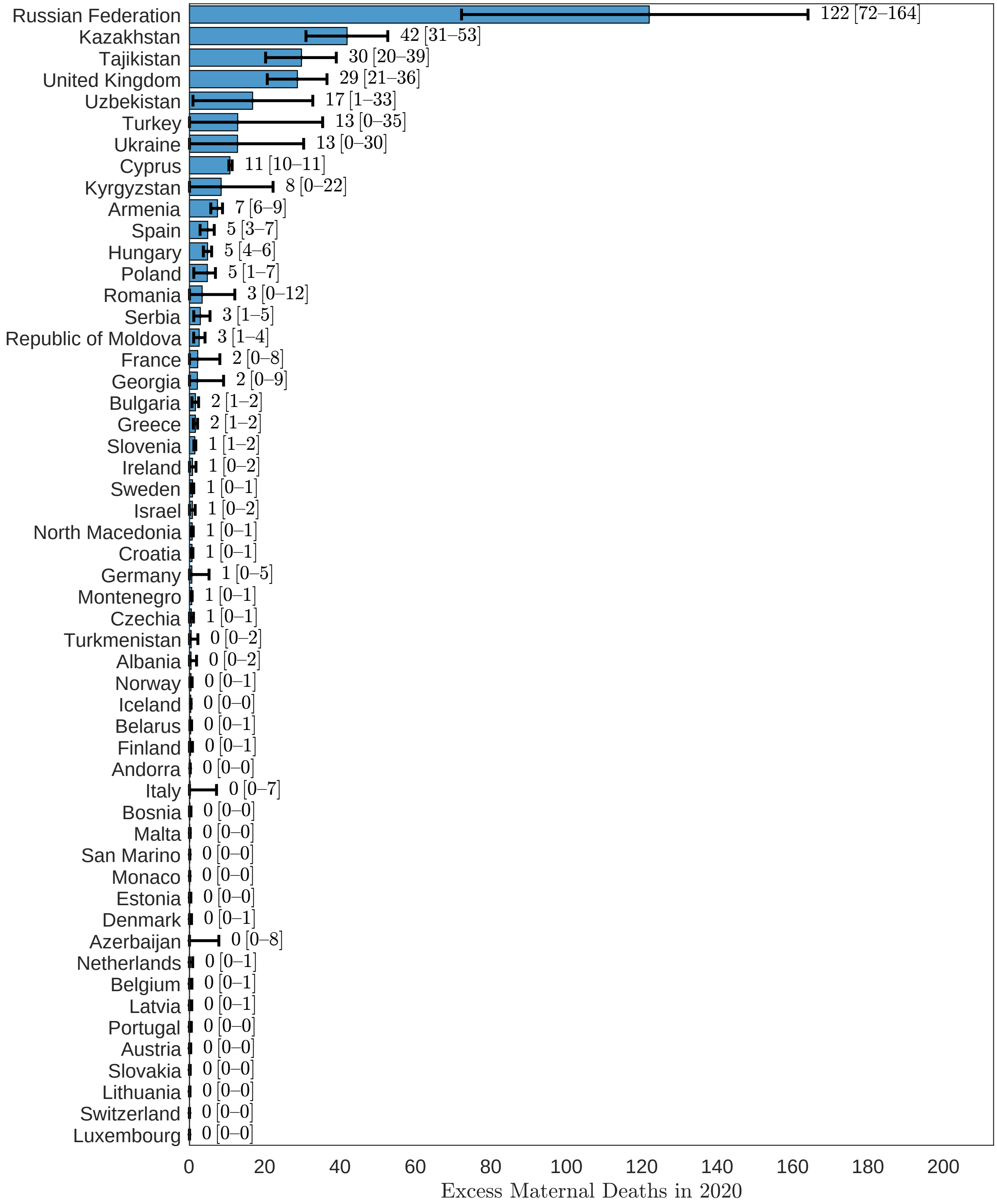


eFigure 17: Excess maternal deaths, European Region, 2020.

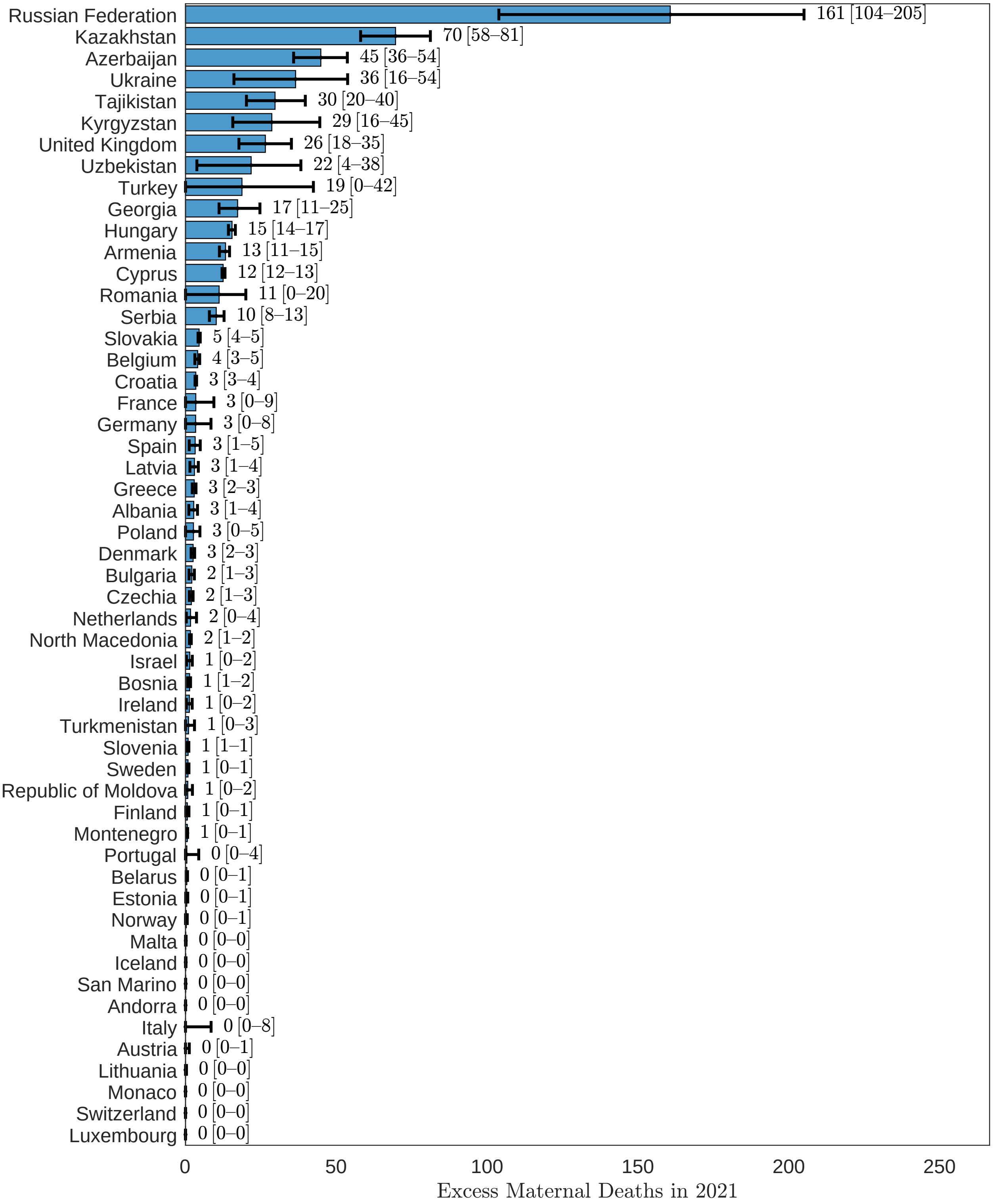


eFigure 18: Excess maternal deaths, European Region, 2021.

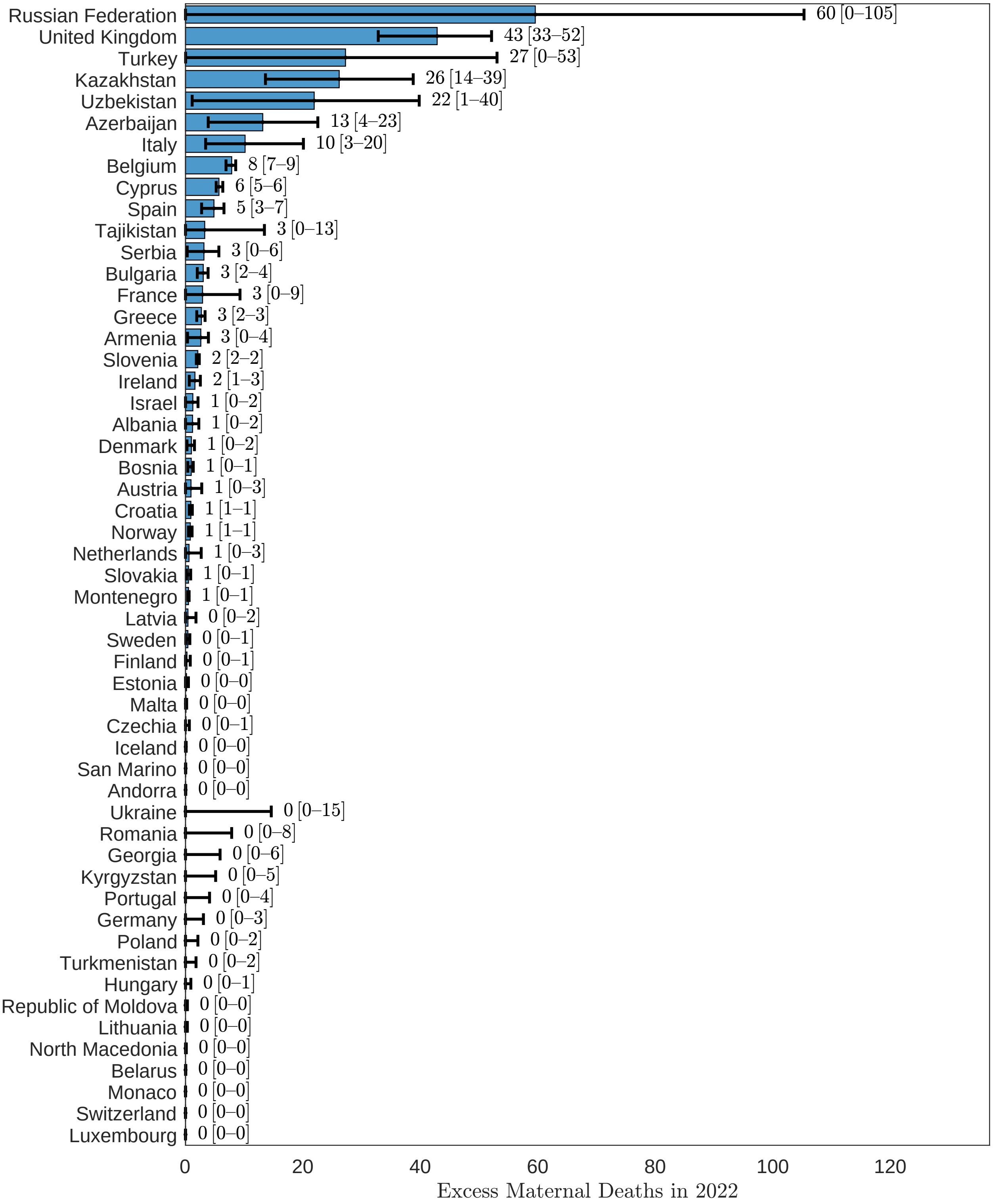


eFigure 19: Excess maternal deaths, European Region, 2022.

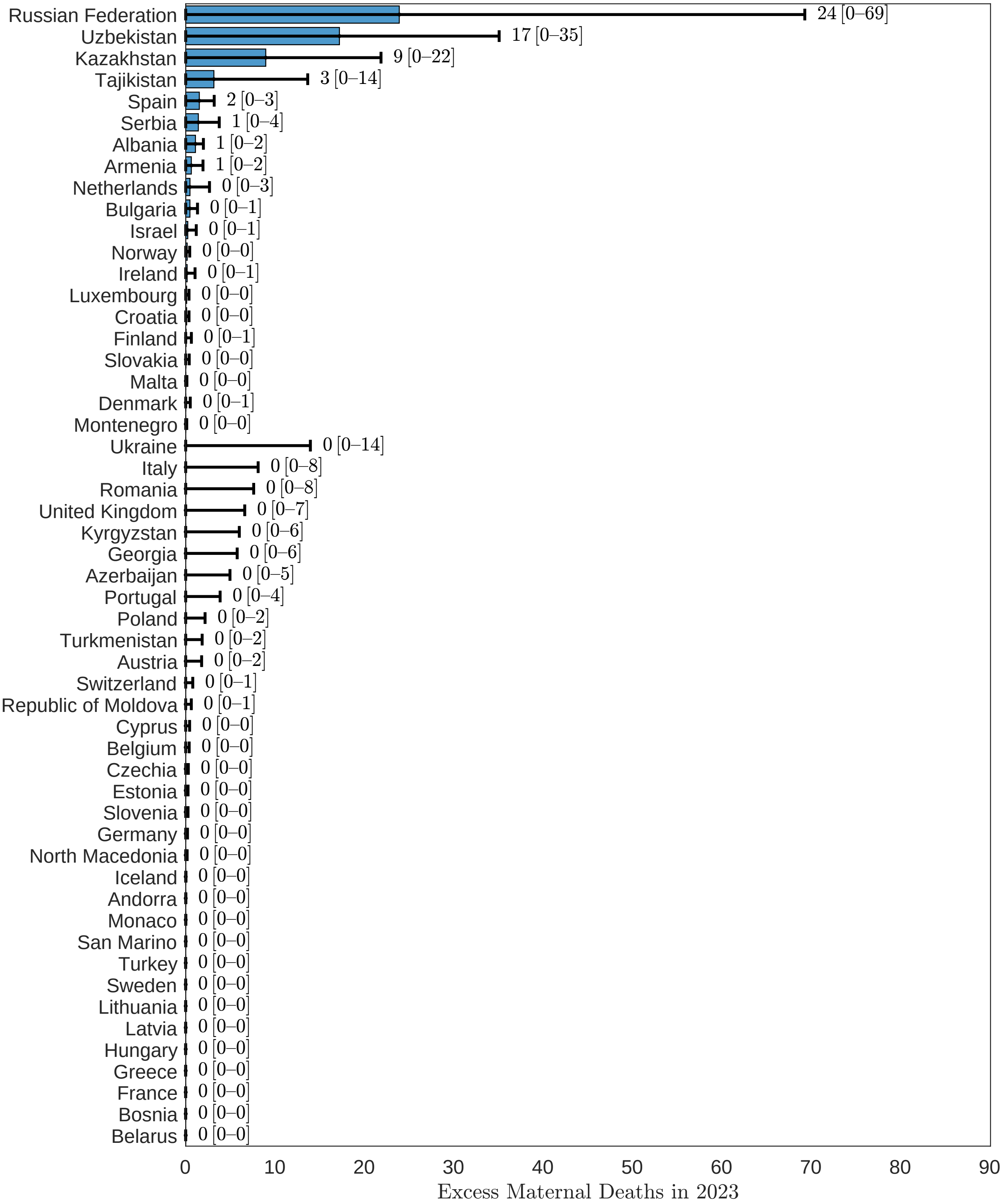


eFigure 20: Excess maternal deaths, European Region, 2023.

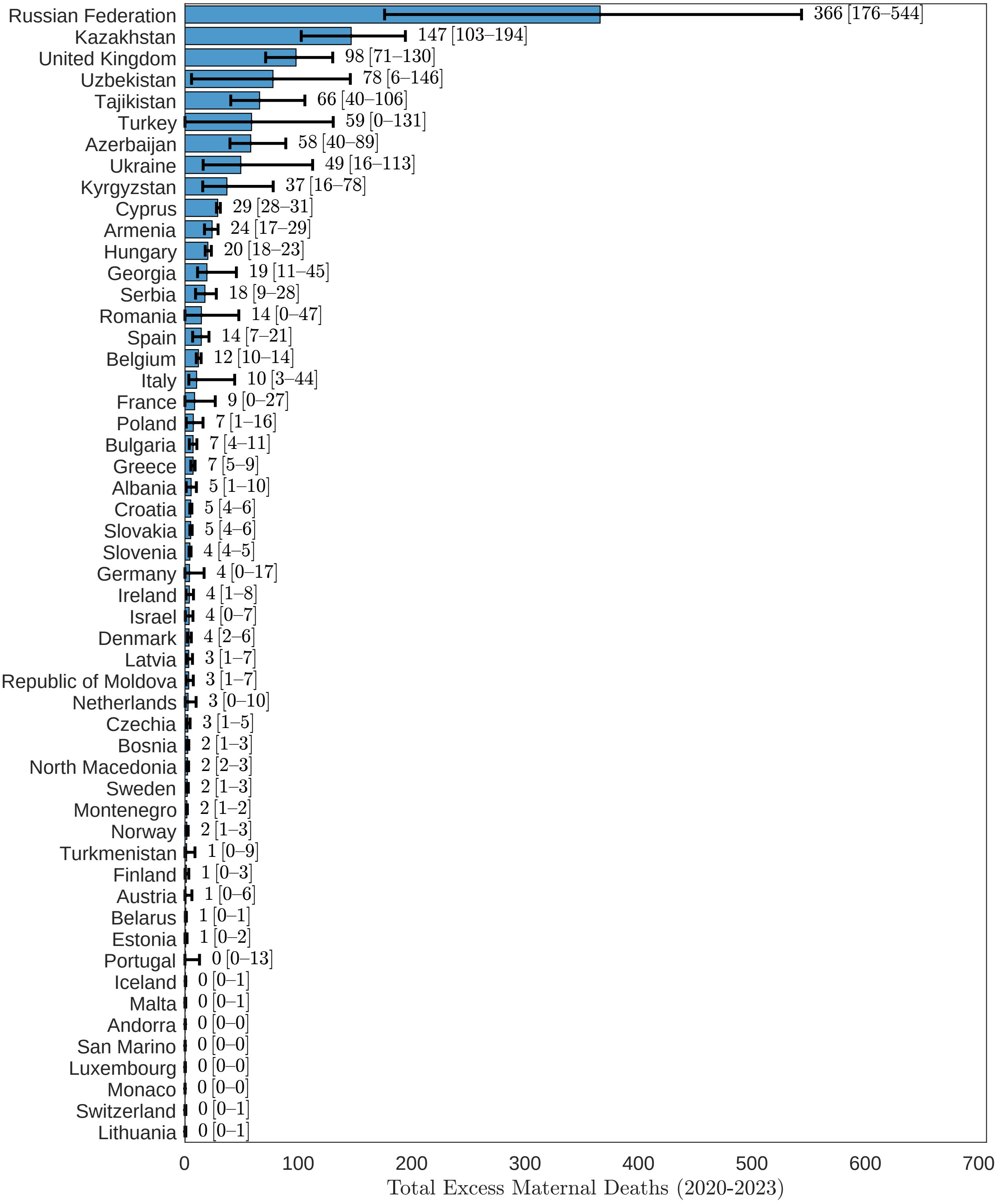


eFigure 21: Excess maternal deaths, European Region, 2020-2023.

##### 3.2.2 MMR

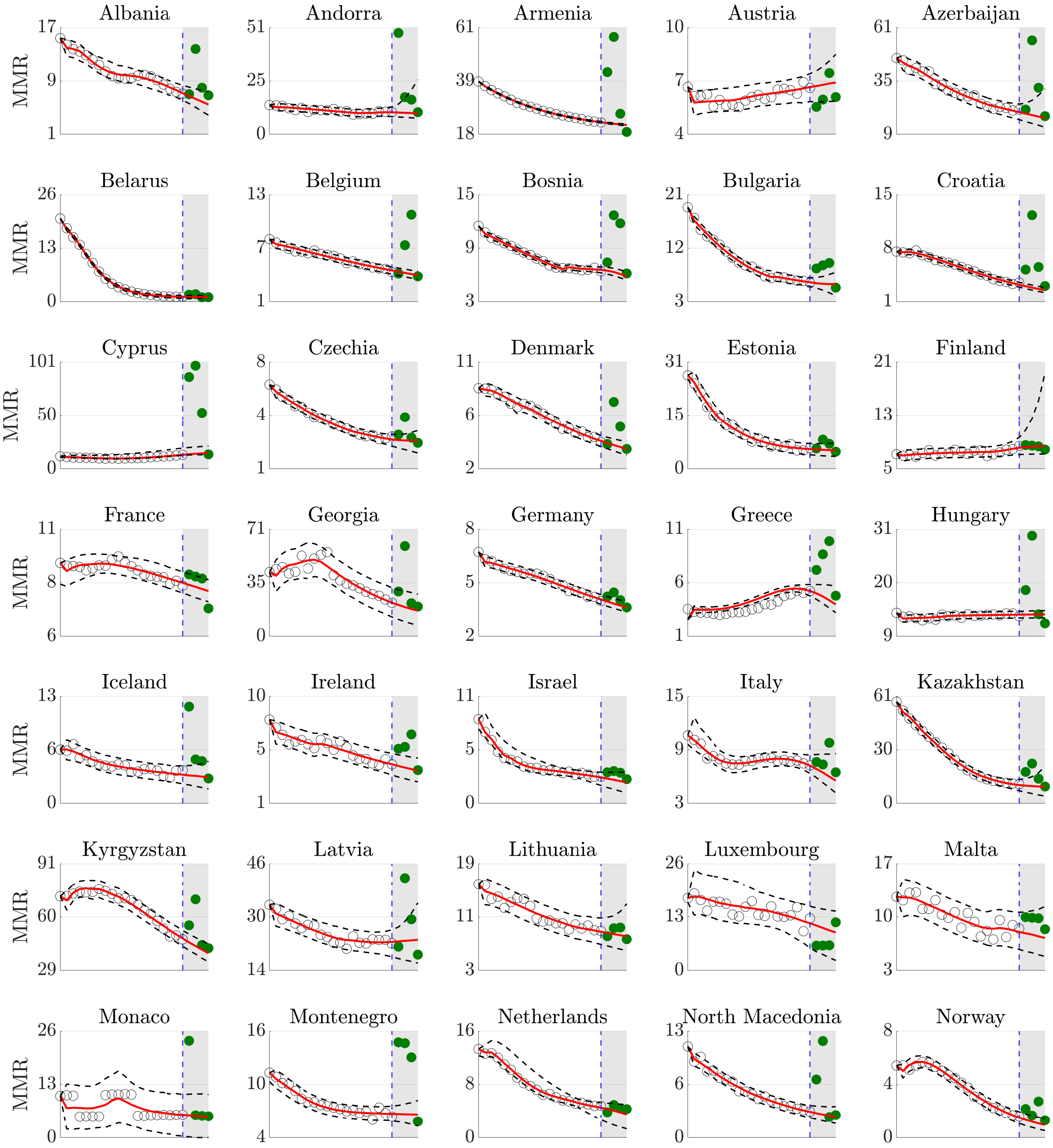


eFigure 22: Counterfactual maternal mortality ratio forecasts, European Region.

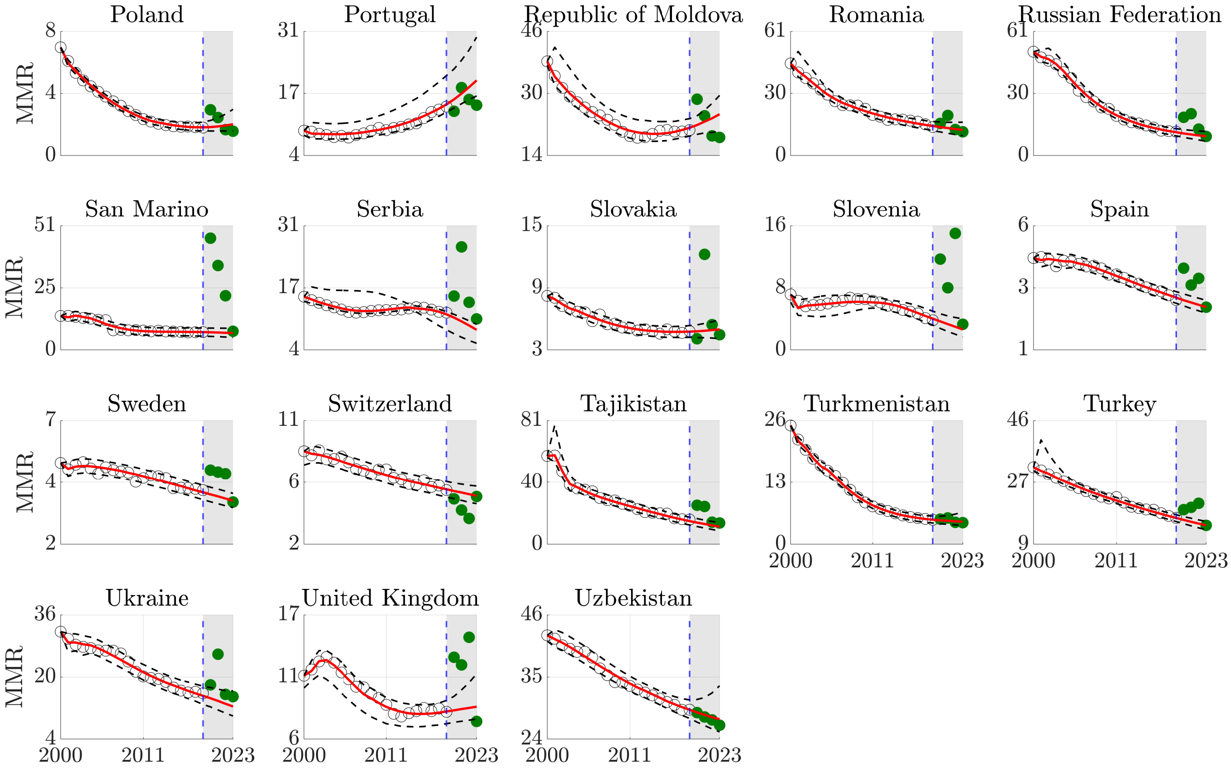


Counterfactual maternal mortality ratio forecasts, European Region (continued).

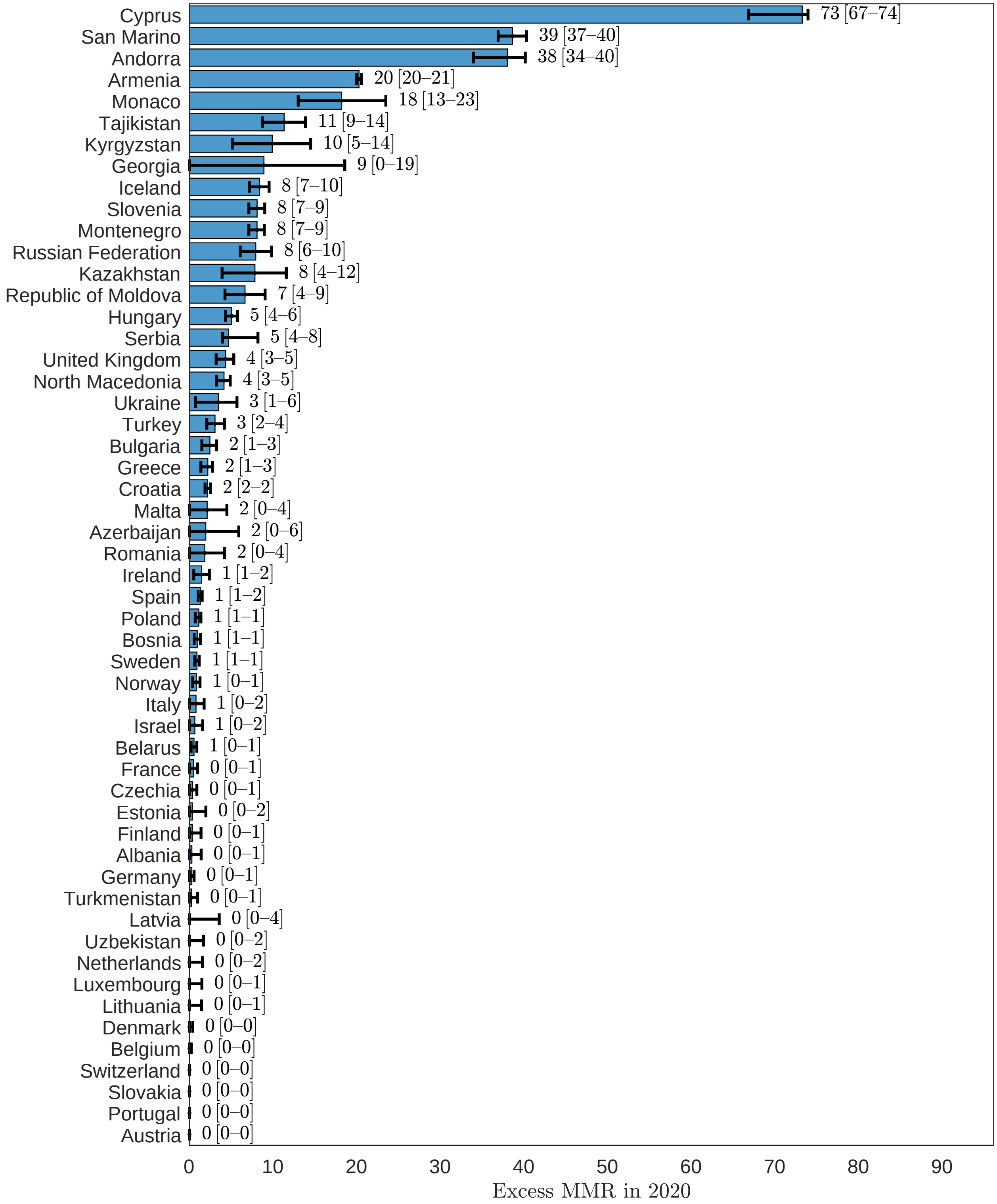


eFigure 23: Excess maternal mortality ratio, European Region, 2020.

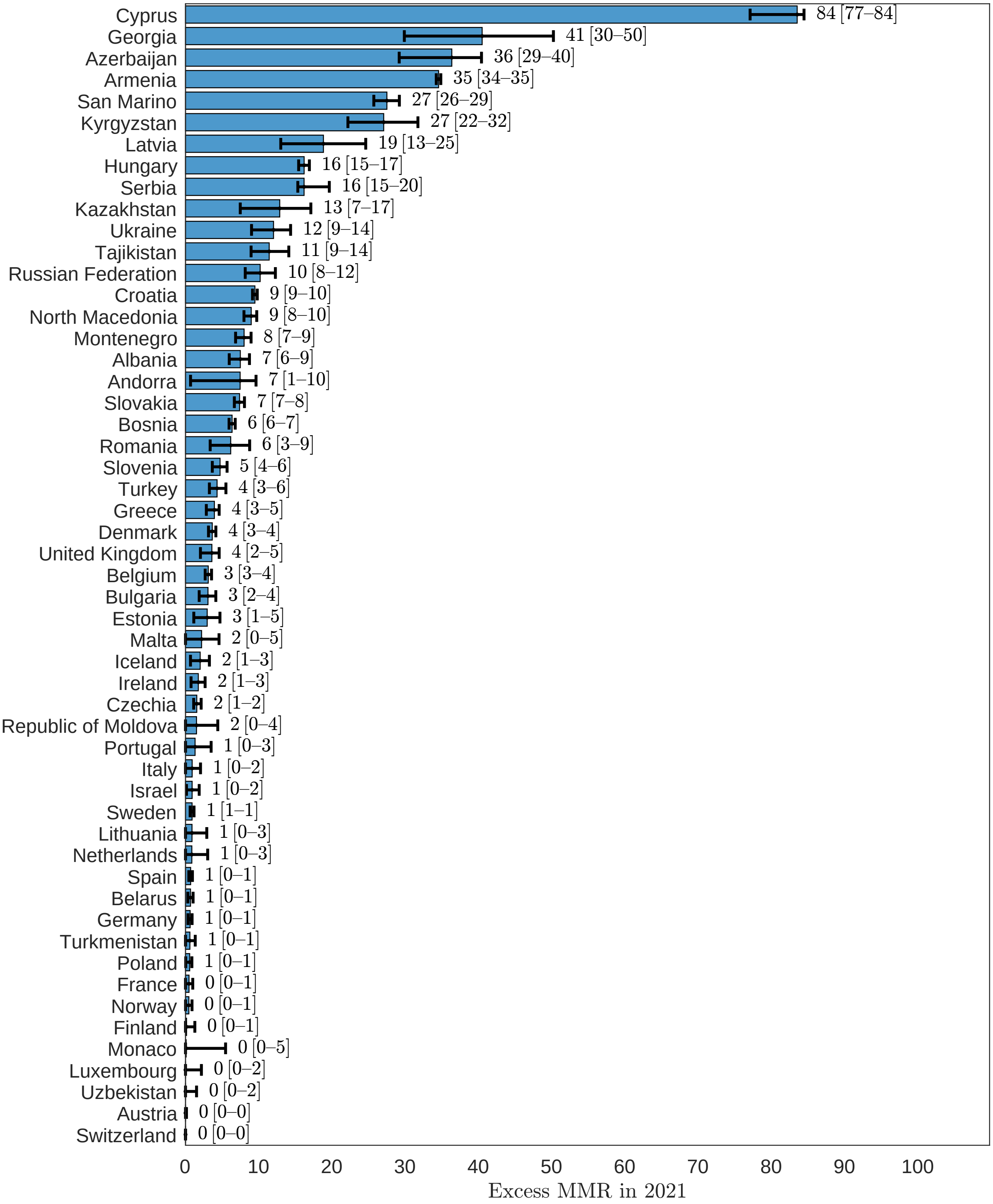


eFigure 24: Excess maternal mortality ratio, European Region, 2021.

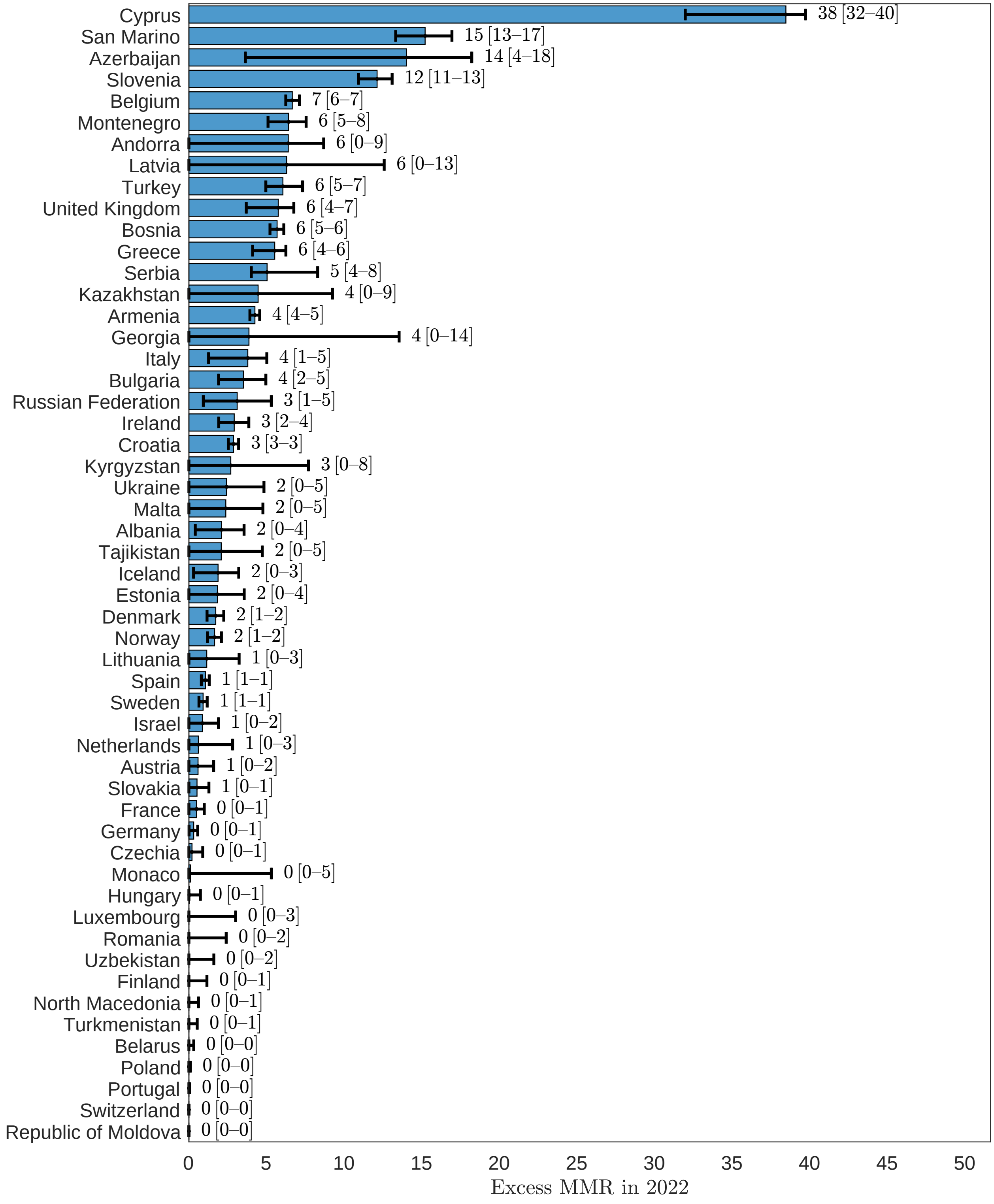


eFigure 25: Excess maternal mortality ratio, European Region, 2022.

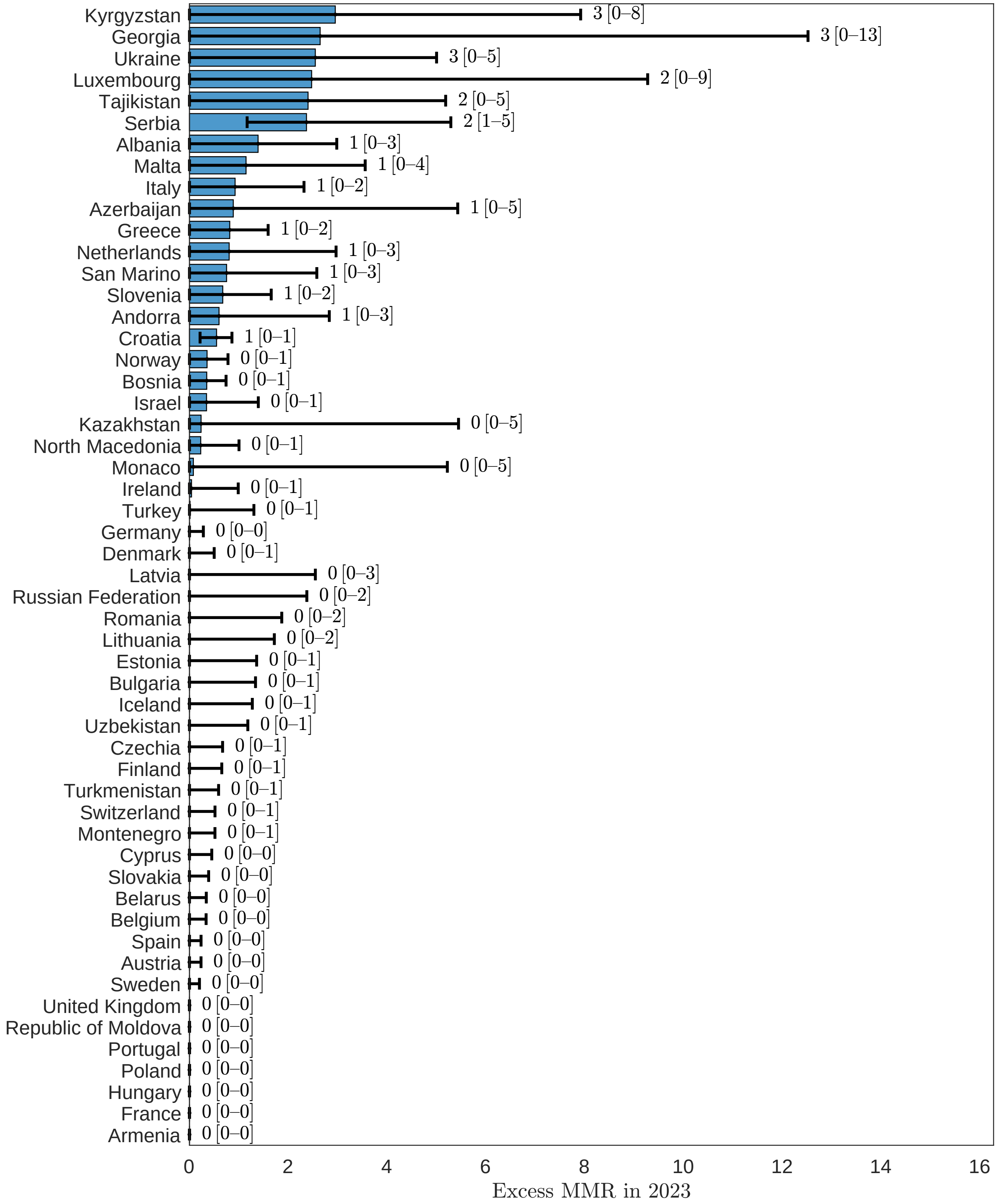


eFigure 26: Excess maternal mortality ratio, European Region, 2023.

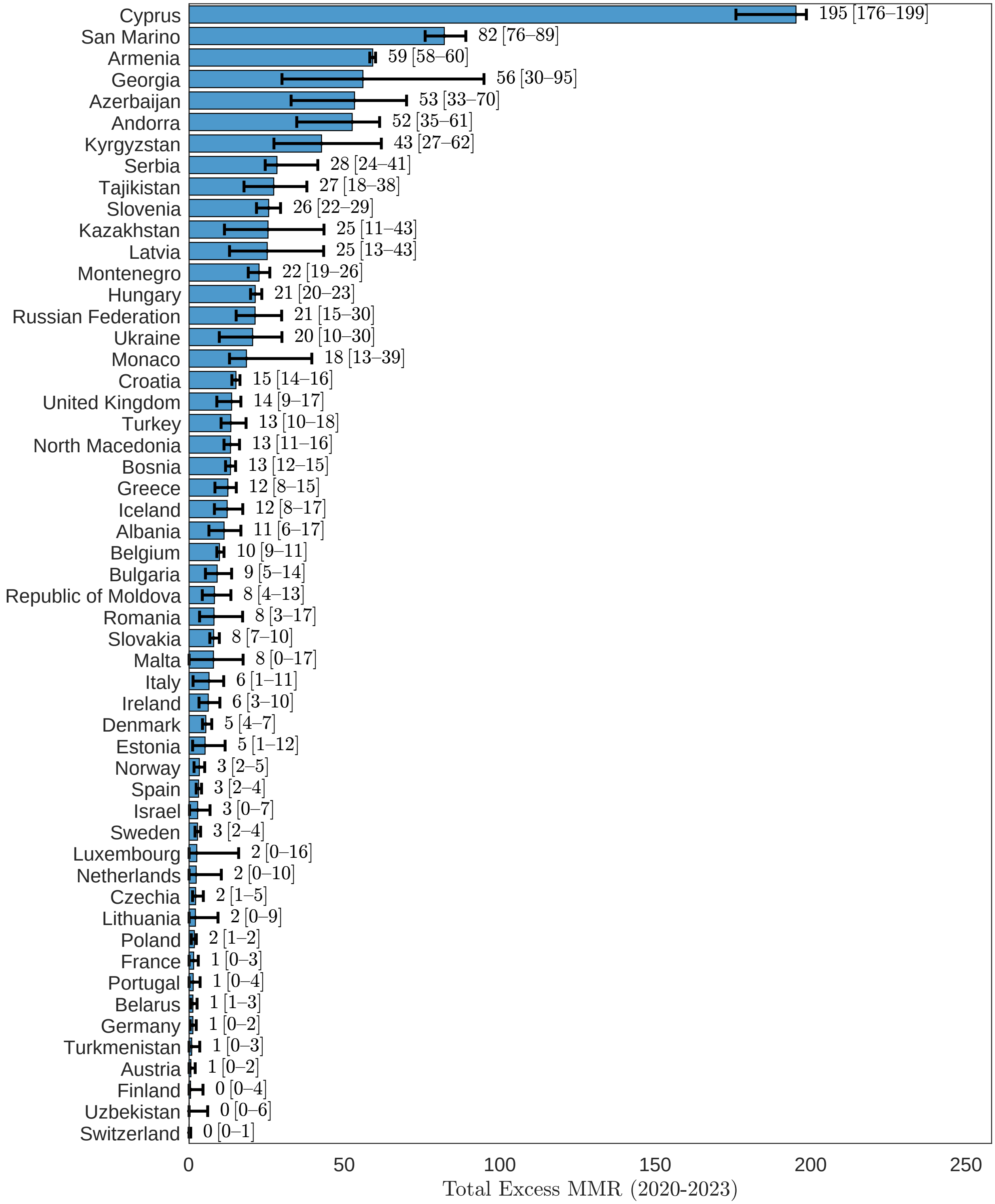


eFigure 27: Excess maternal mortality ratio, European Region, 2020-2023.

#### 3.3 Eastern Mediterranean Region

##### 3.3.1 Maternal deaths

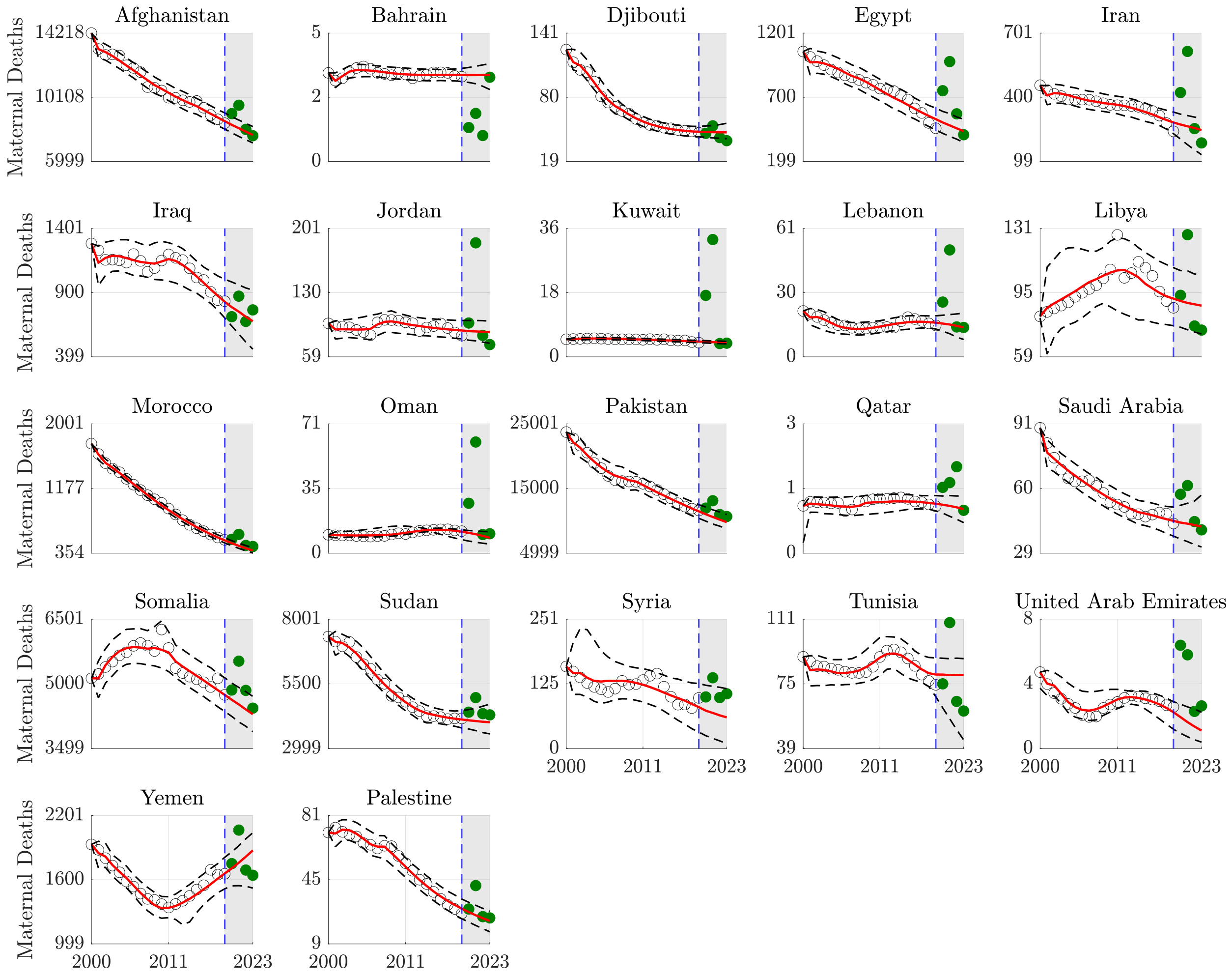


eFigure 28: Counterfactual maternal death forecasts, Eastern Mediterranean.

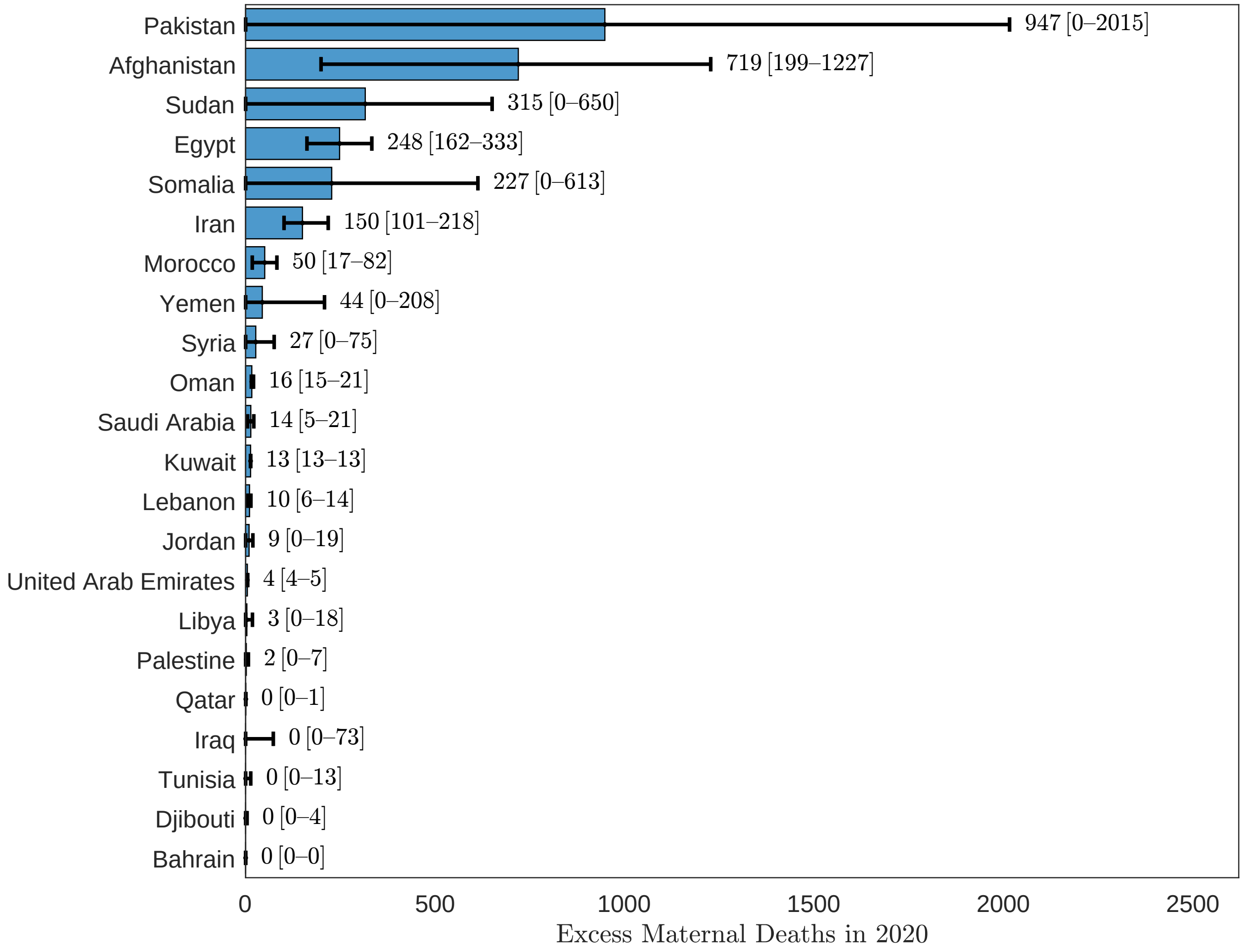


eFigure 29: Excess maternal deaths, Eastern Mediterranean, 2020.

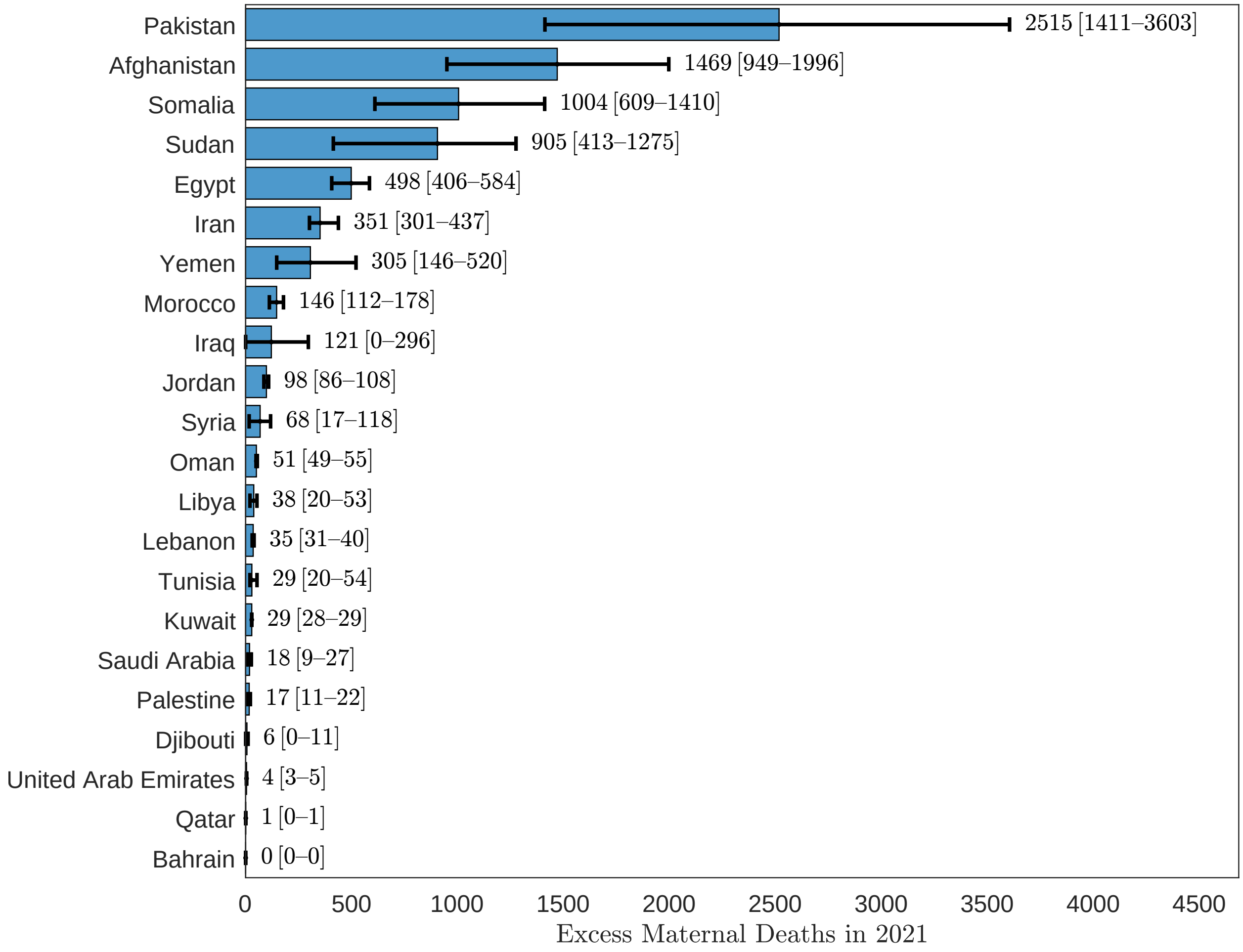


eFigure 30: Excess maternal deaths, Eastern Mediterranean, 2021.

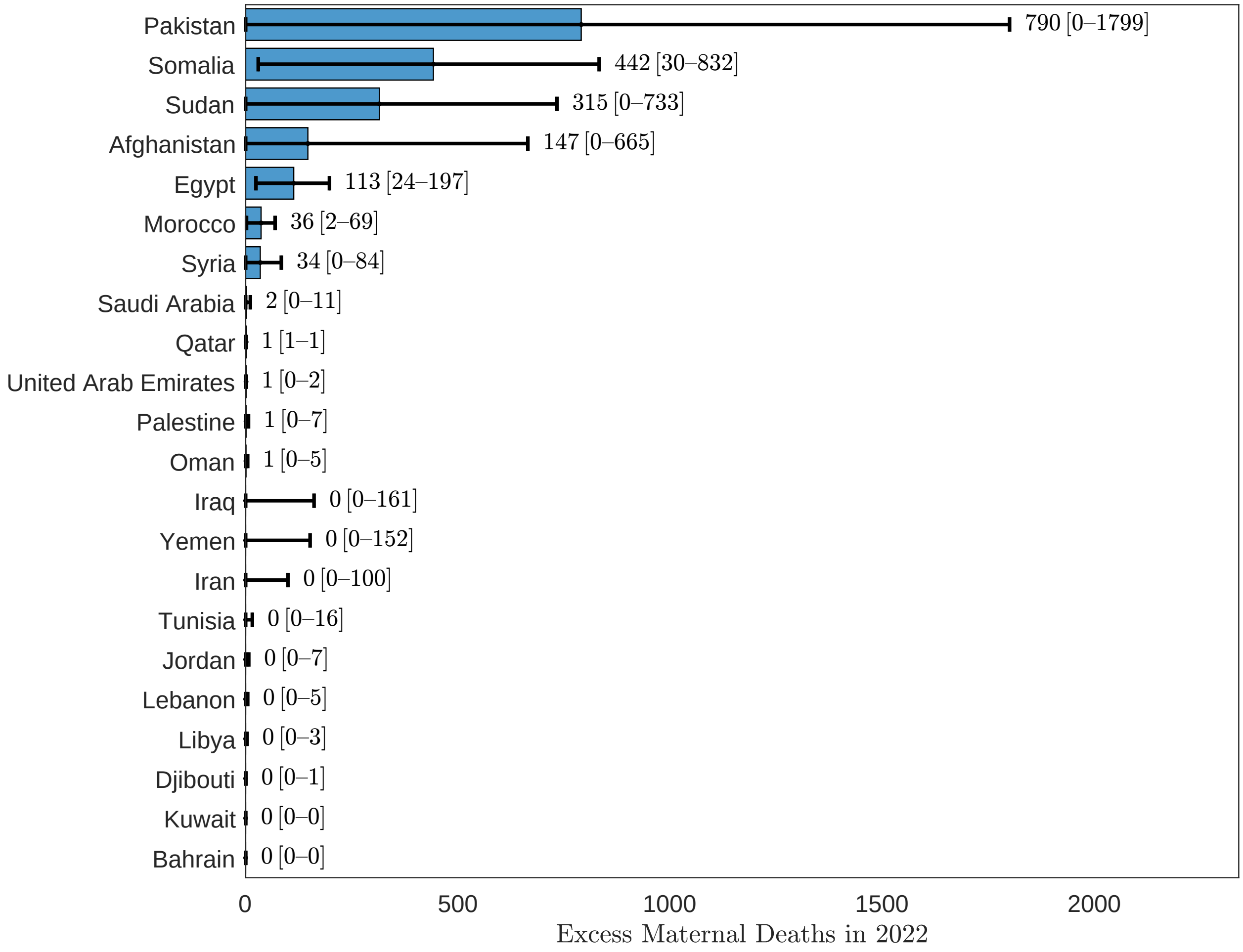


eFigure 31: Excess maternal deaths, Eastern Mediterranean, 2022.

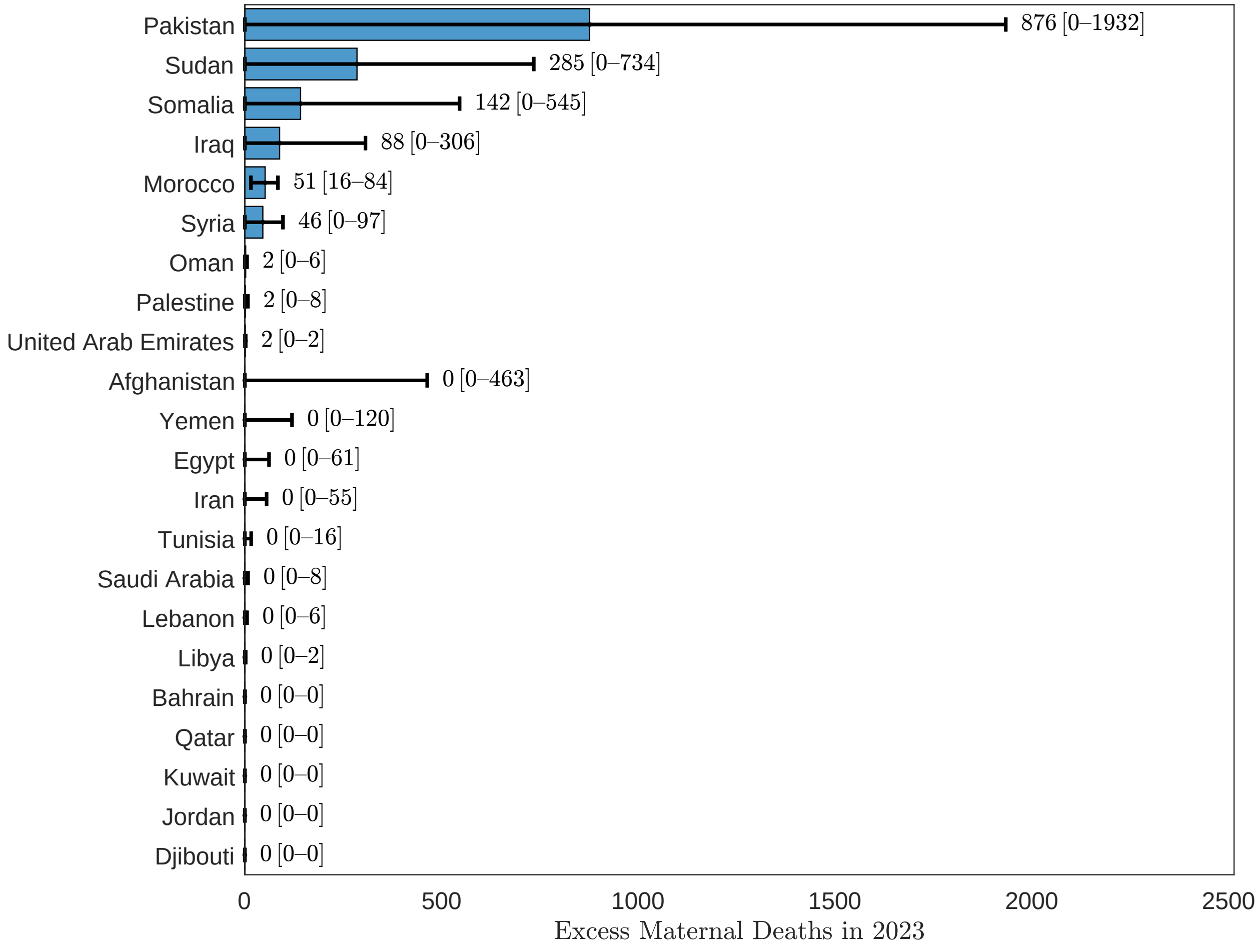


eFigure 32: Excess maternal deaths, Eastern Mediterranean, 2023.

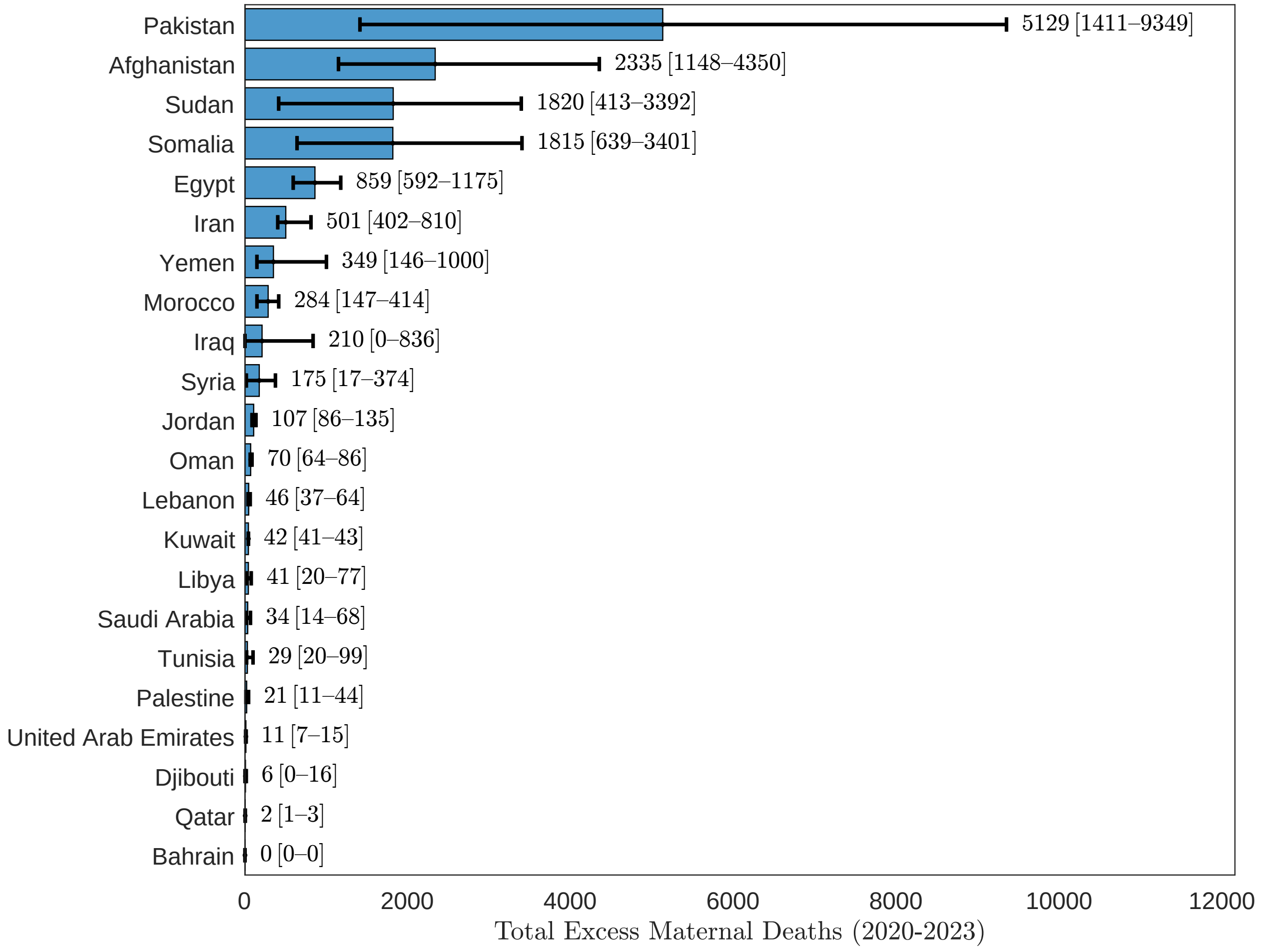


eFigure 33: Excess maternal deaths, Eastern Mediterranean, 2020-2023.

##### 3.3.2 MMR

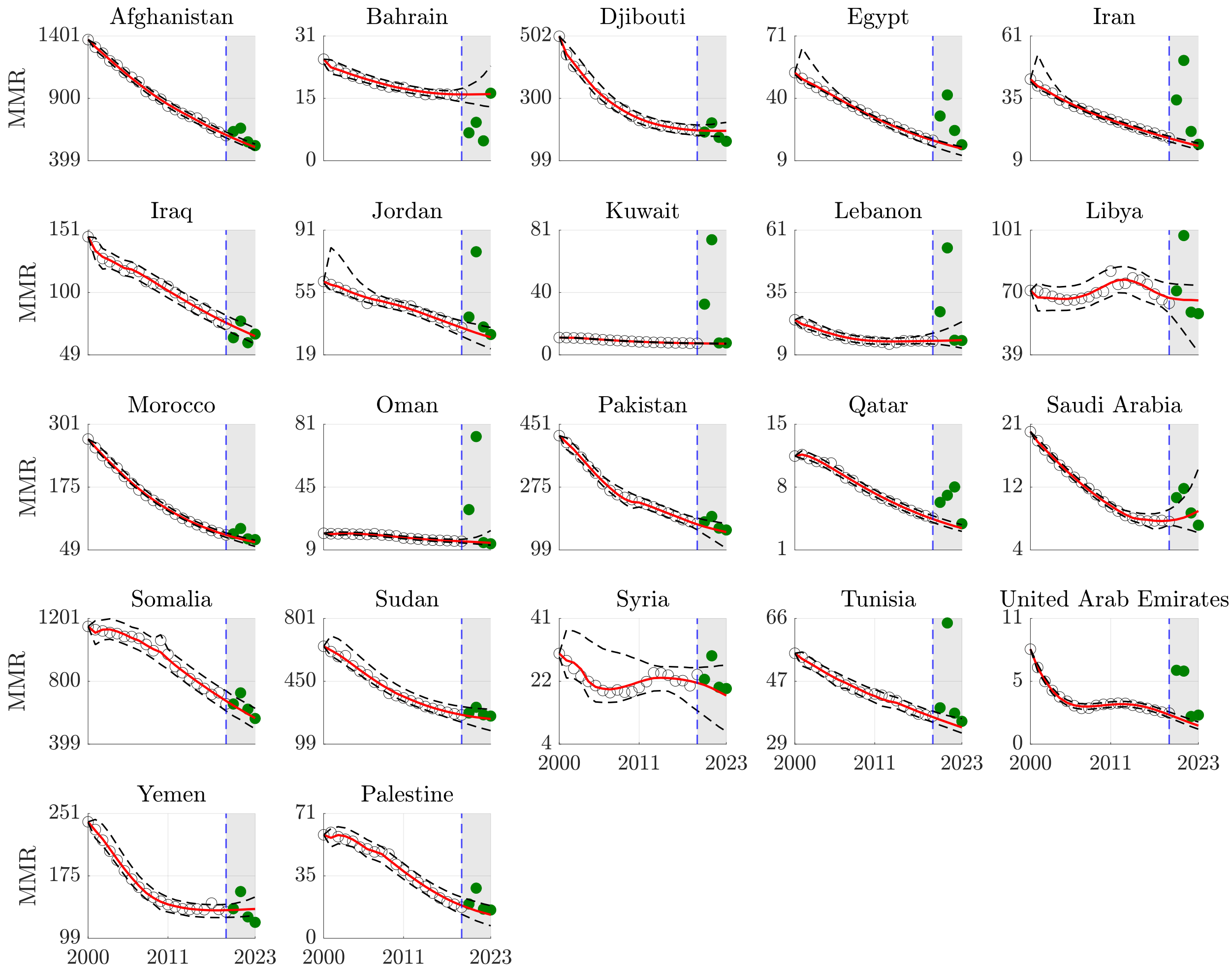


eFigure 34: Counterfactual maternal mortality ratio forecasts, Eastern Mediterranean.

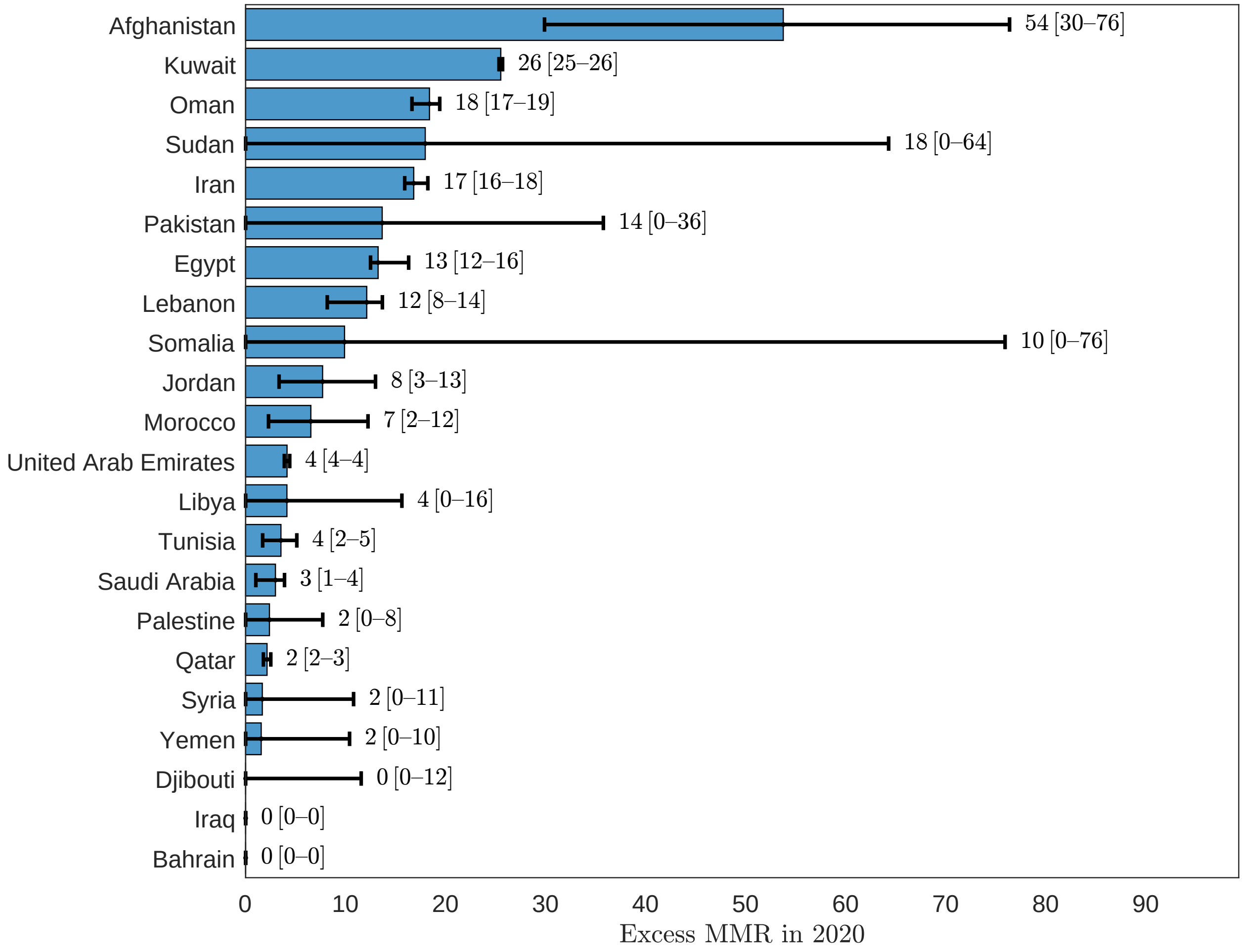


eFigure 35: Excess maternal mortality ratio, Eastern Mediterranean, 2020.

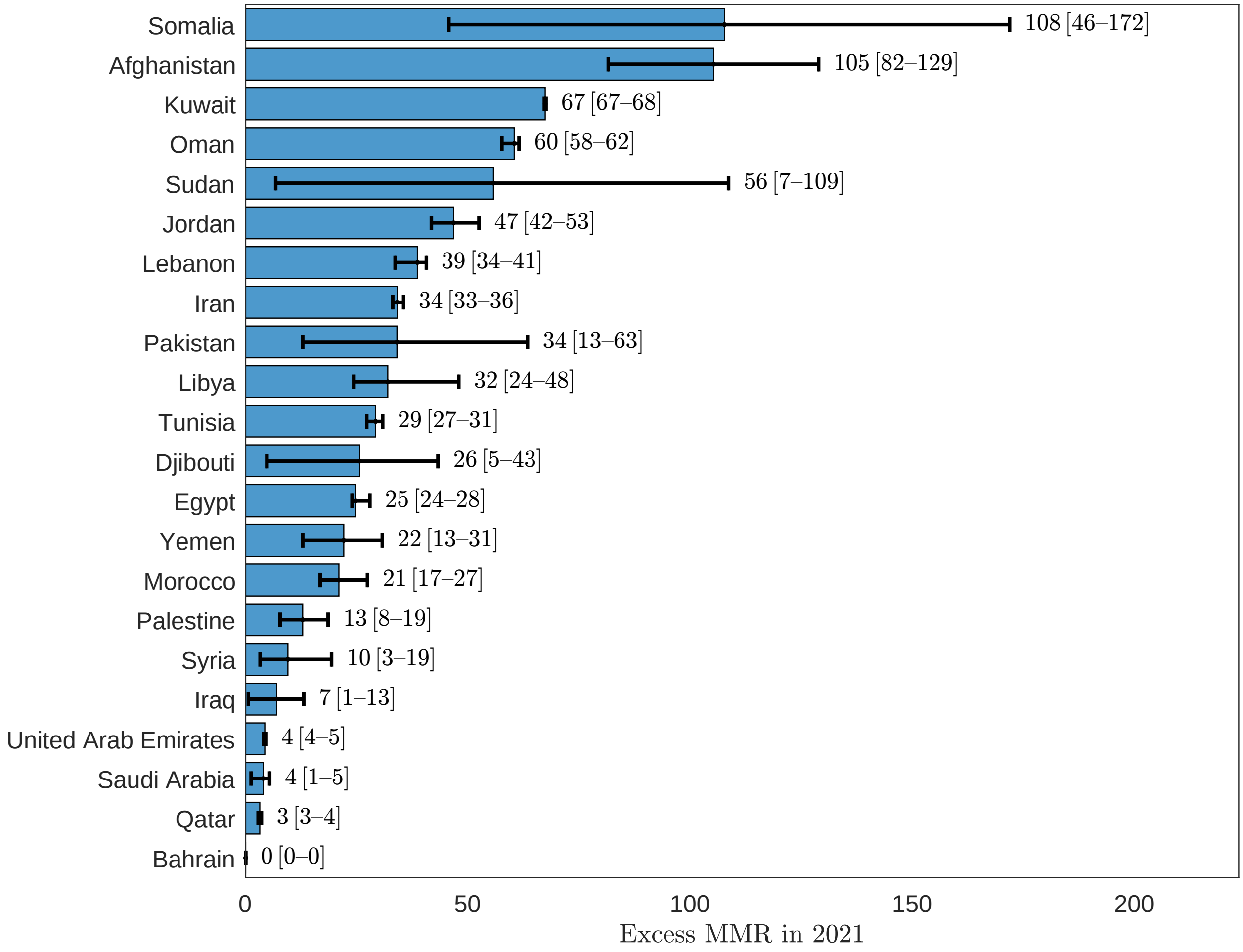


eFigure 36: Excess maternal mortality ratio, Eastern Mediterranean, 2021.

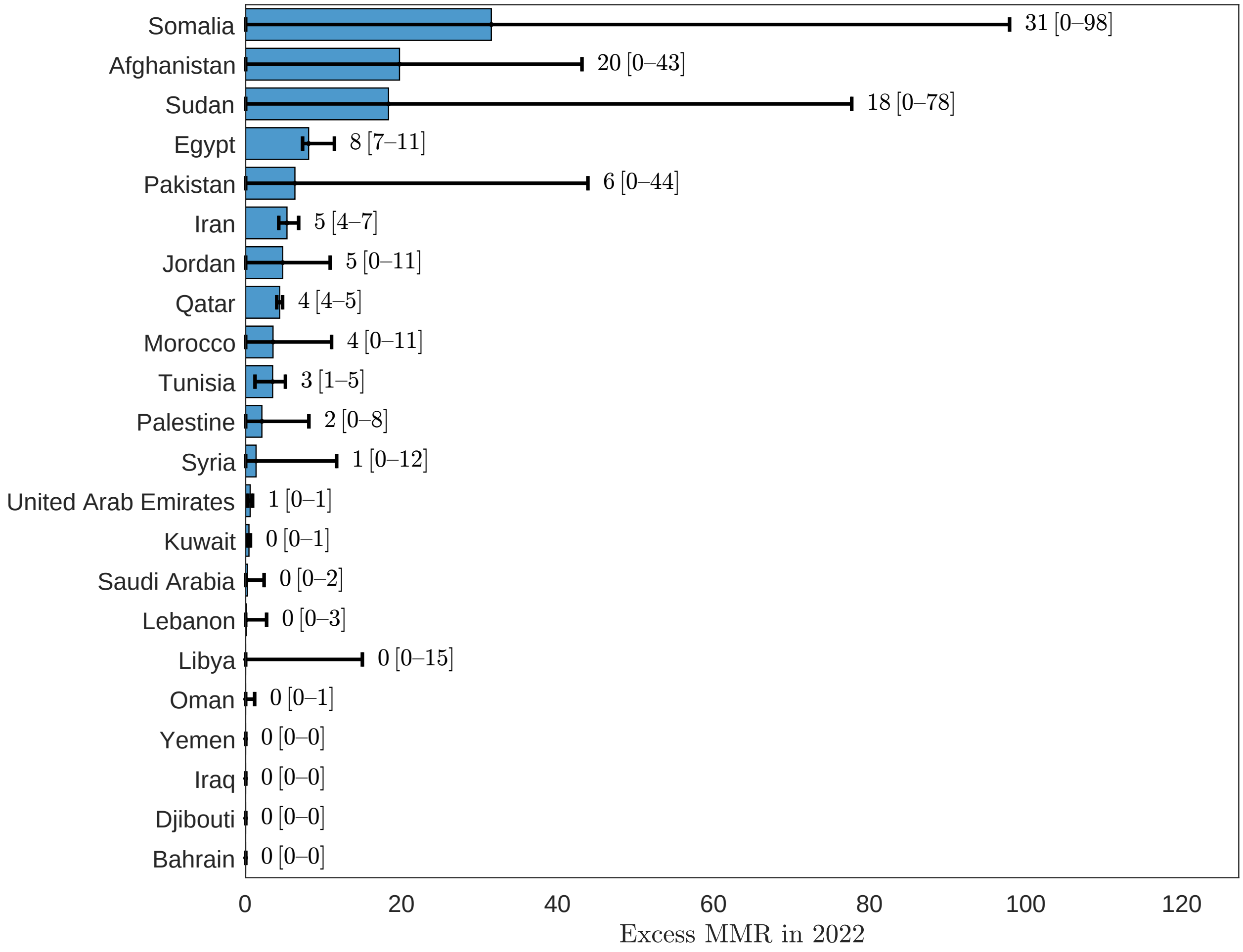


eFigure 37: Excess maternal mortality ratio, Eastern Mediterranean, 2022.

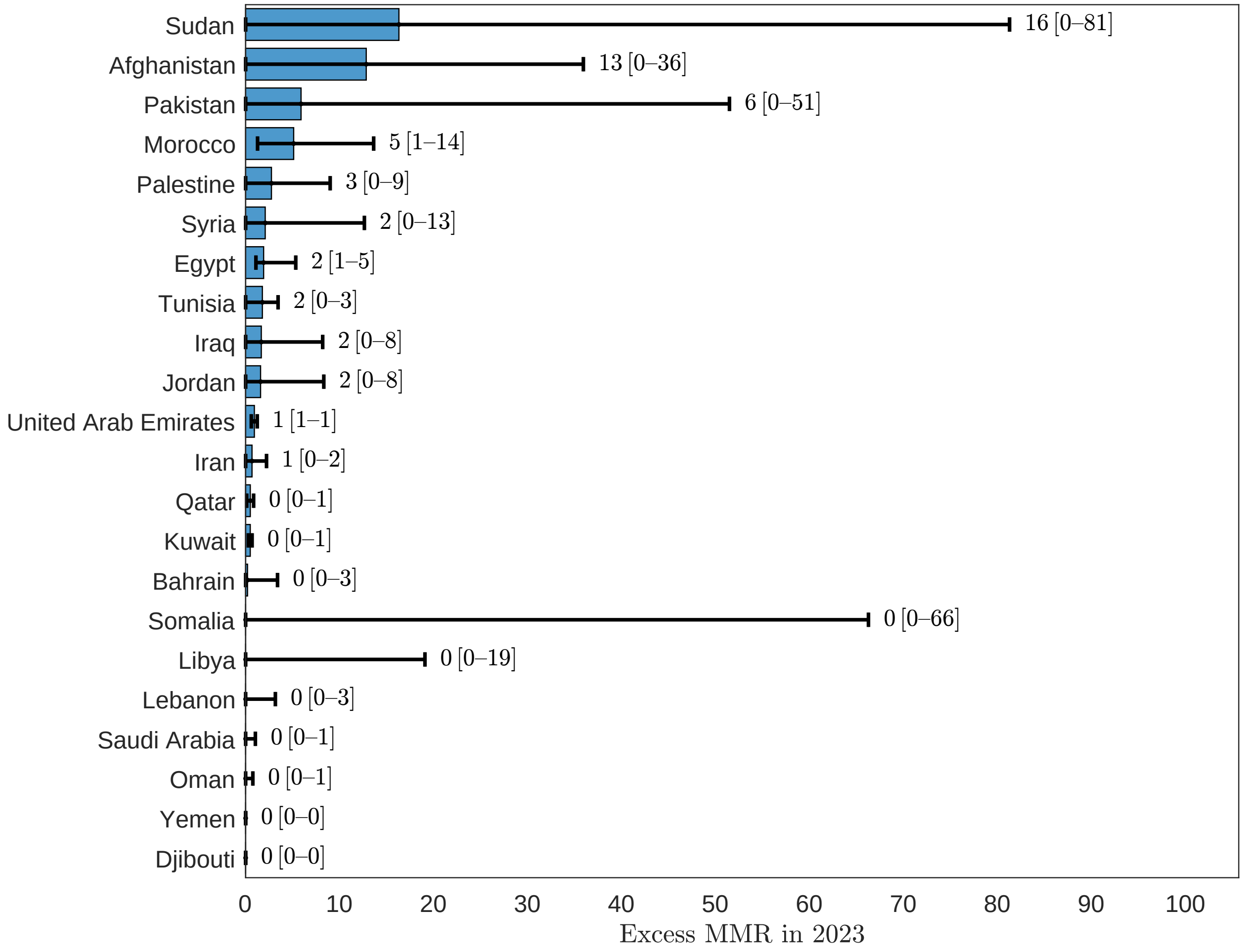


eFigure 38: Excess maternal mortality ratio, Eastern Mediterranean, 2023.

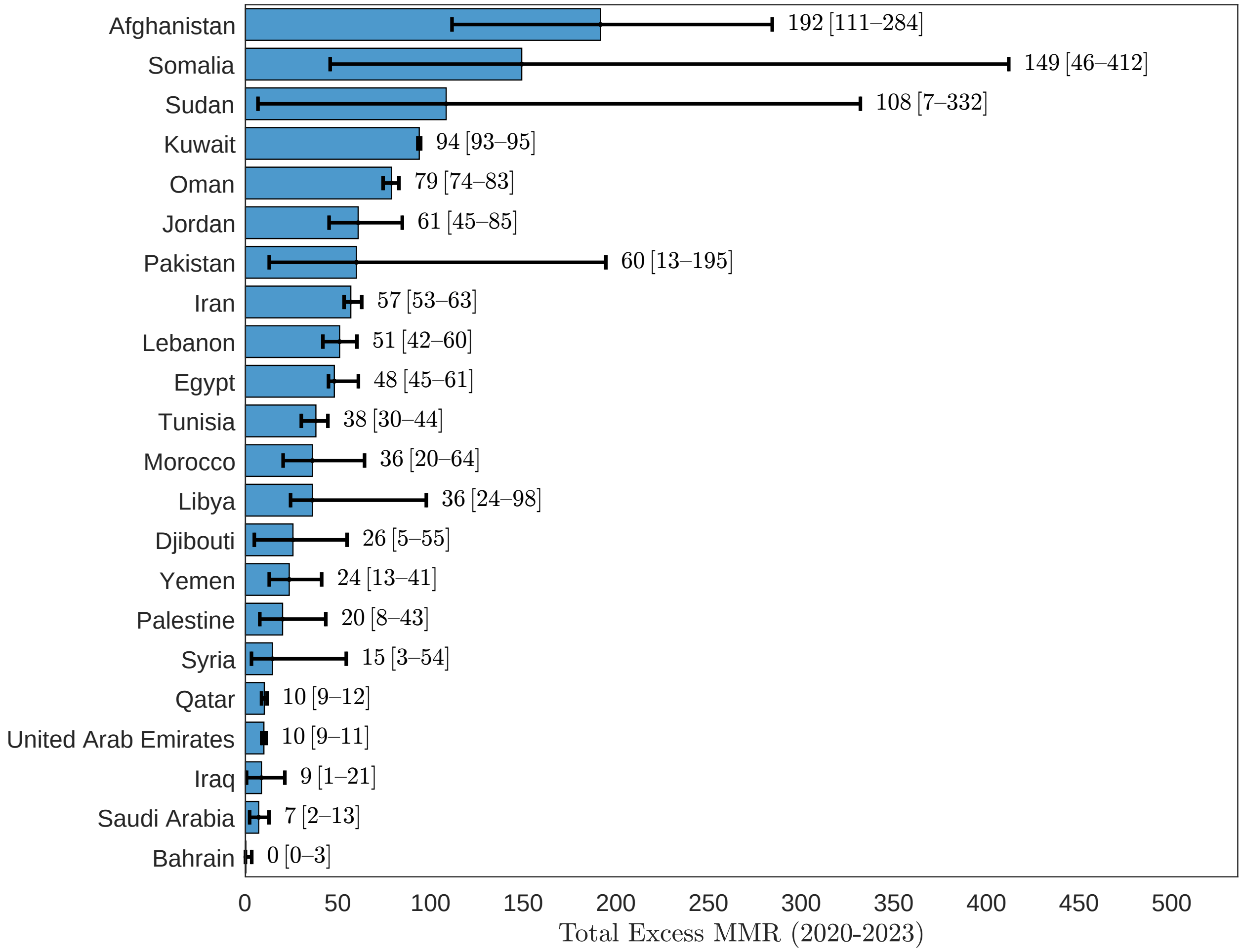


eFigure 39: Excess maternal mortality ratio, Eastern Mediterranean, 2020-2023.

#### 3.4 African Region

##### 3.4.1 Maternal deaths

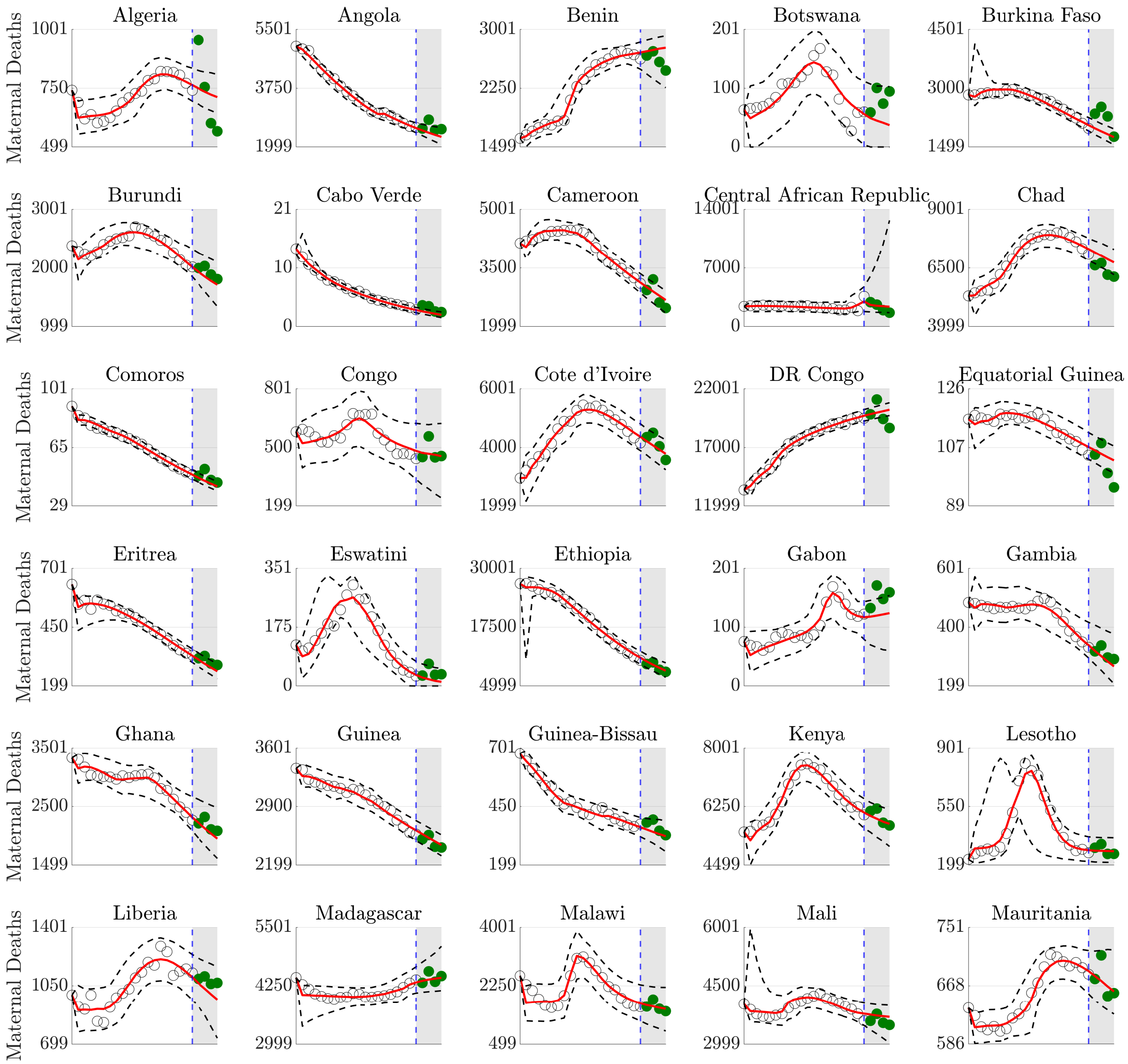


eFigure 40: Counterfactual maternal death forecasts, African Region.

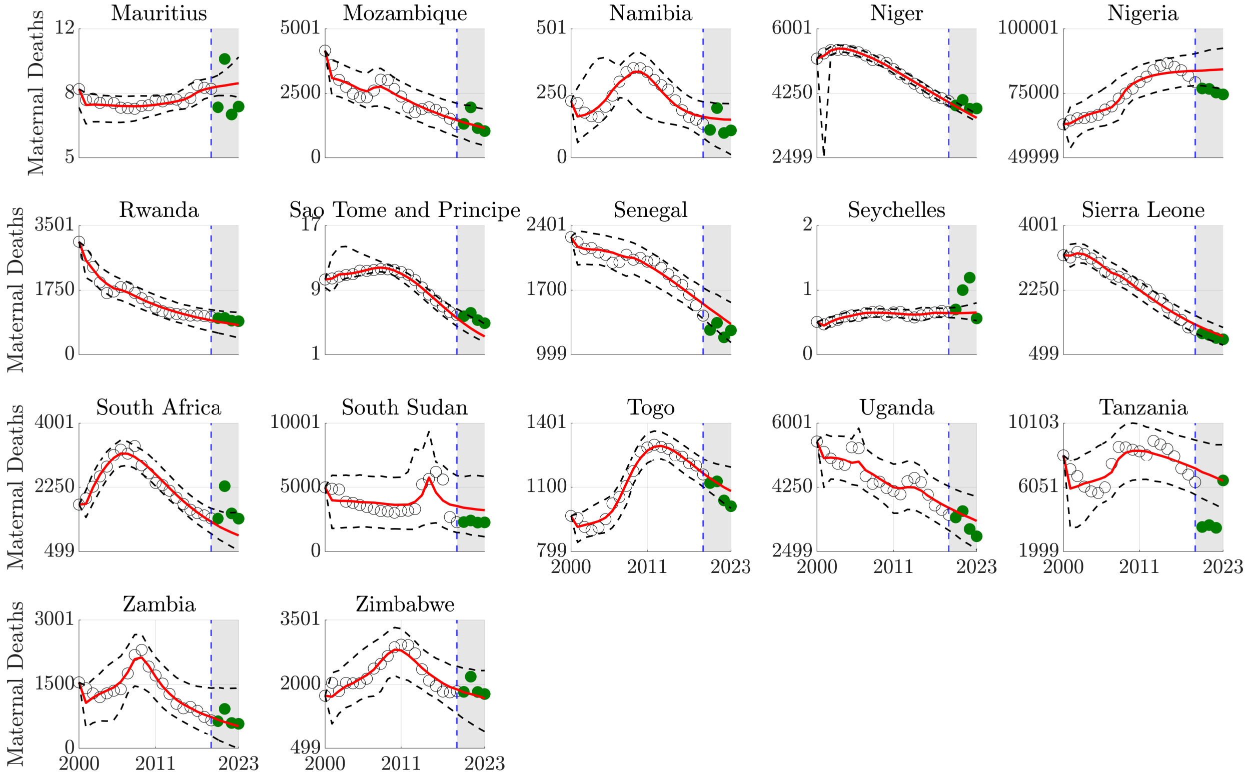


Counterfactual maternal death forecasts, African Region (continued).

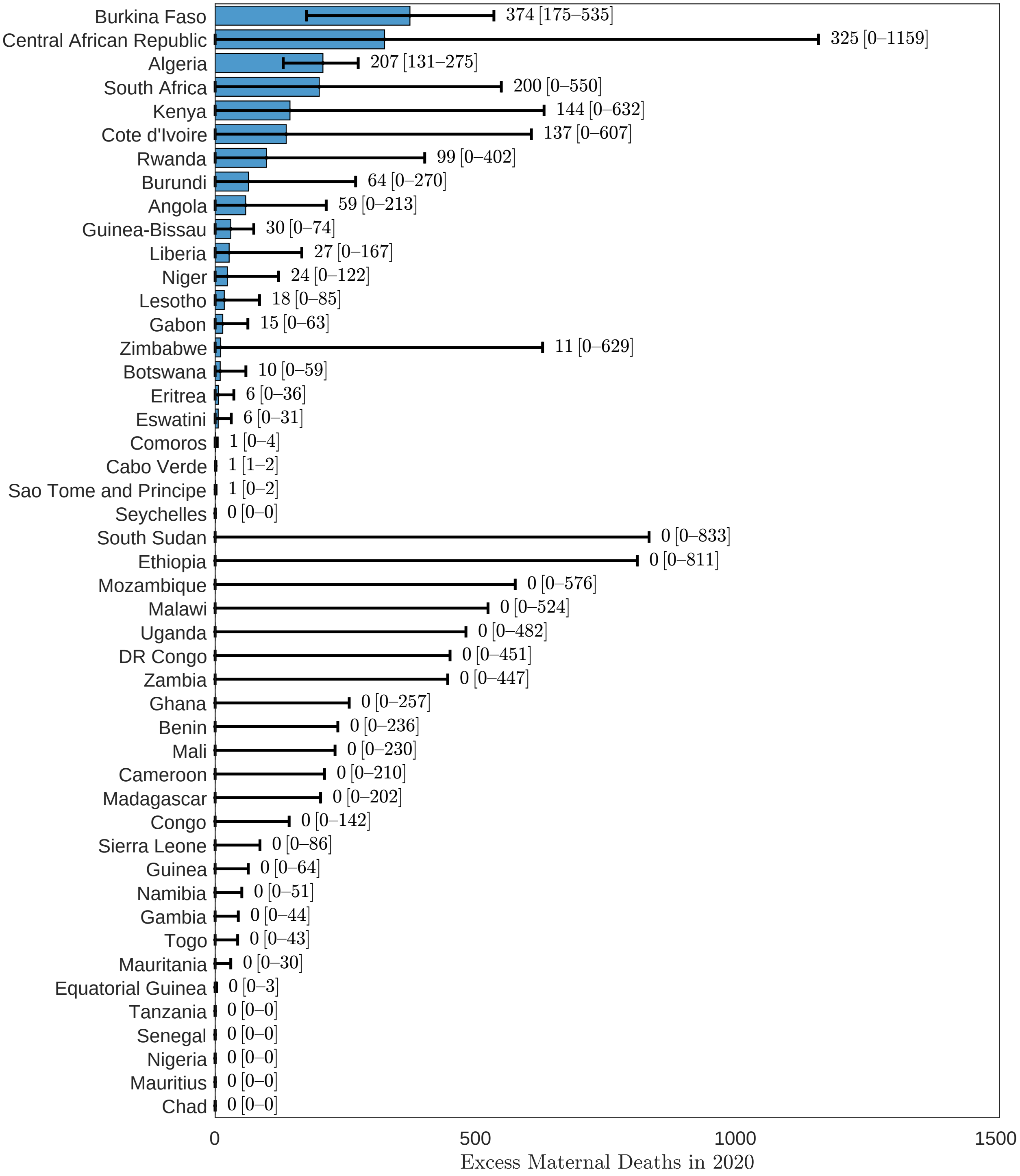


eFigure 41: Excess maternal deaths, African Region, 2020.

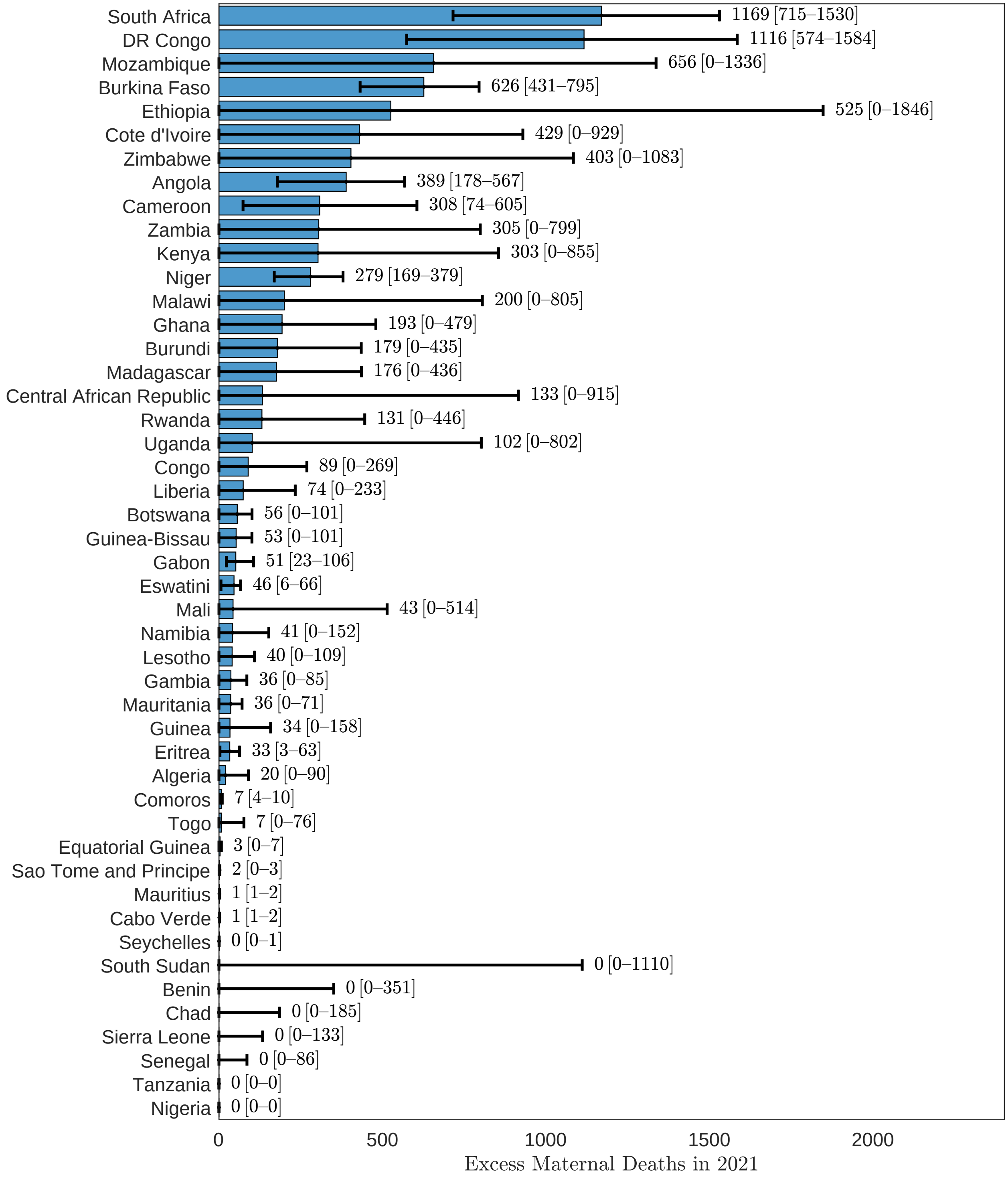


eFigure 42: Excess maternal deaths, African Region, 2021.

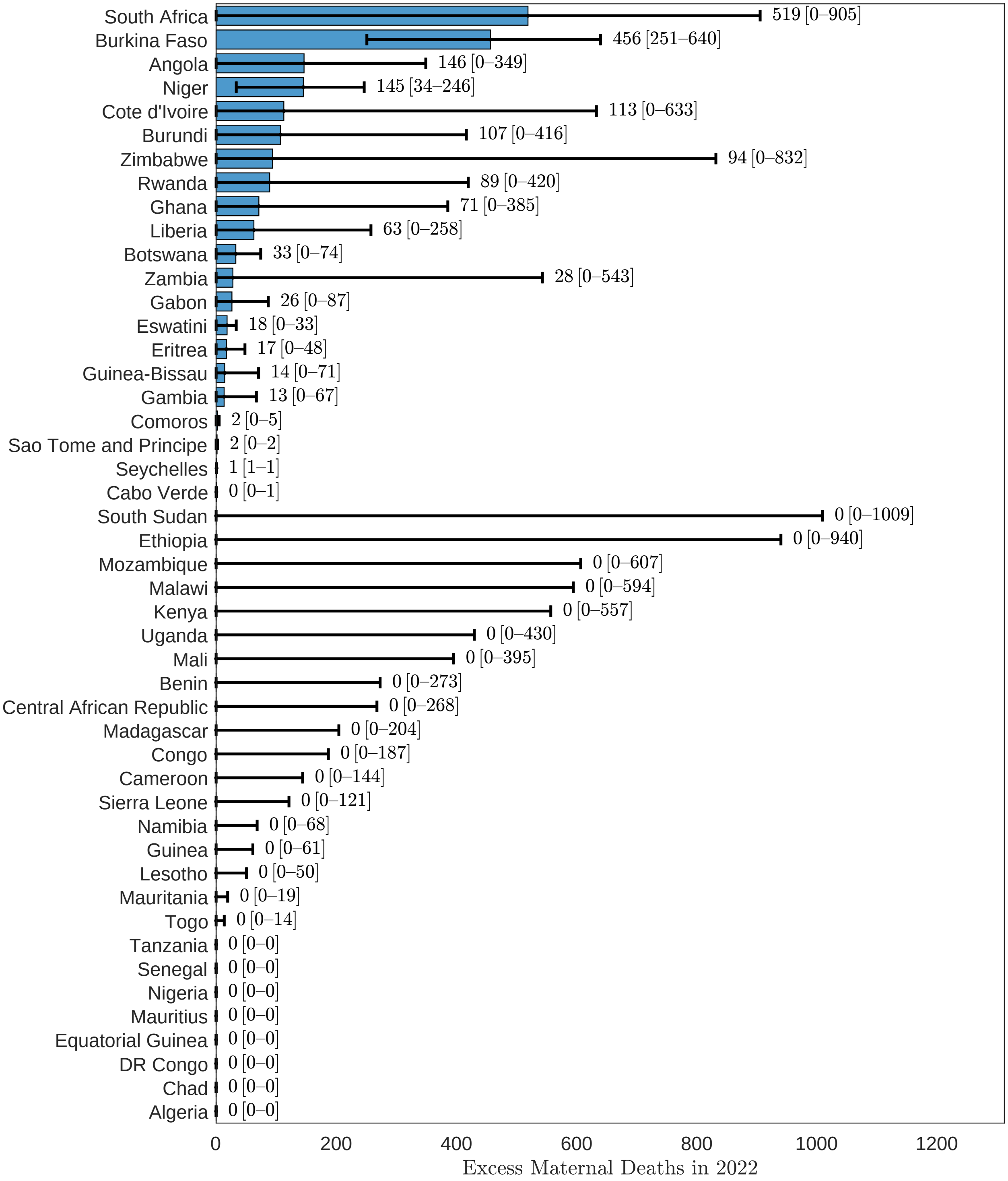


eFigure 43: Excess maternal deaths, African Region, 2022.

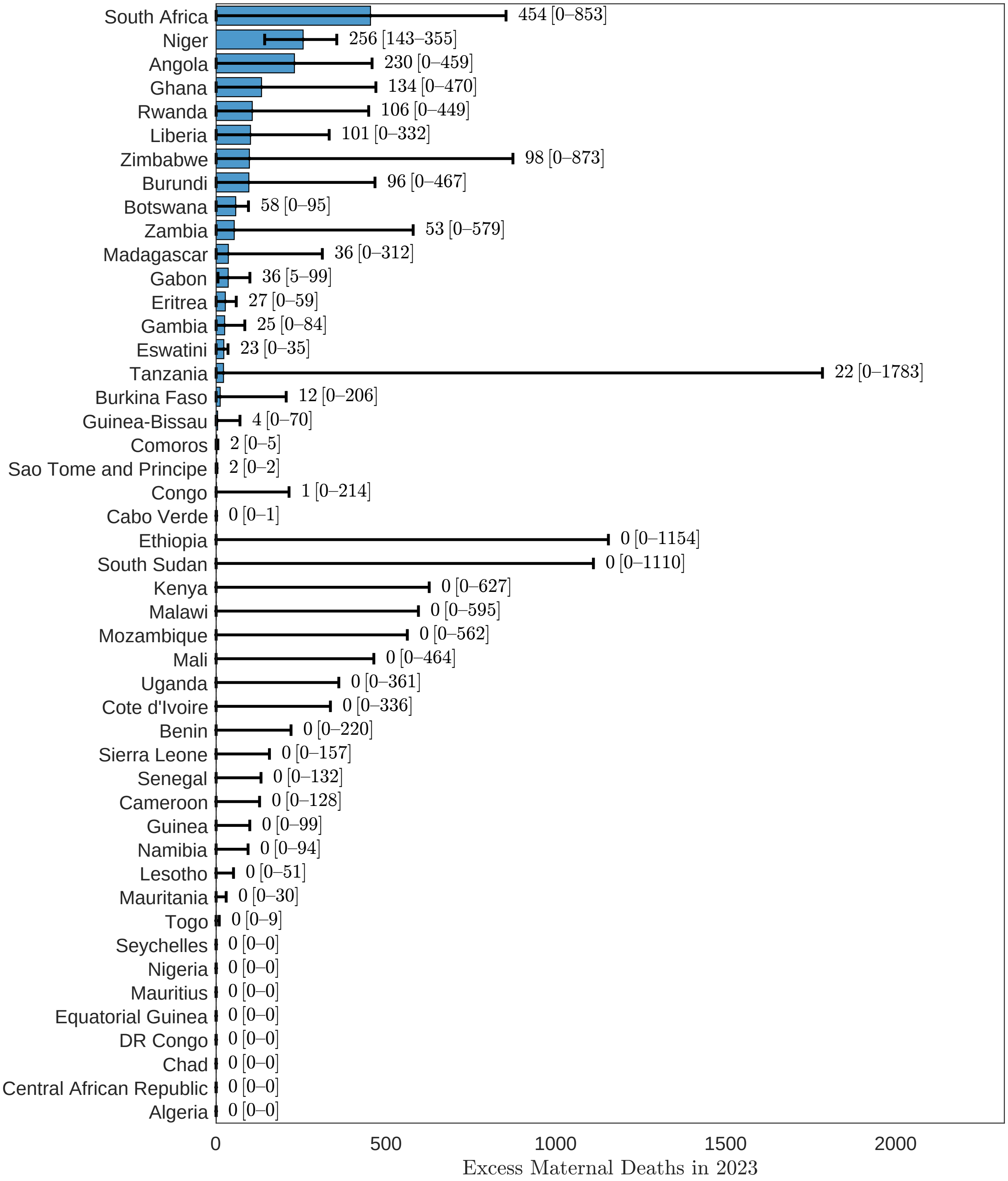


eFigure 44: Excess maternal deaths, African Region, 2023.

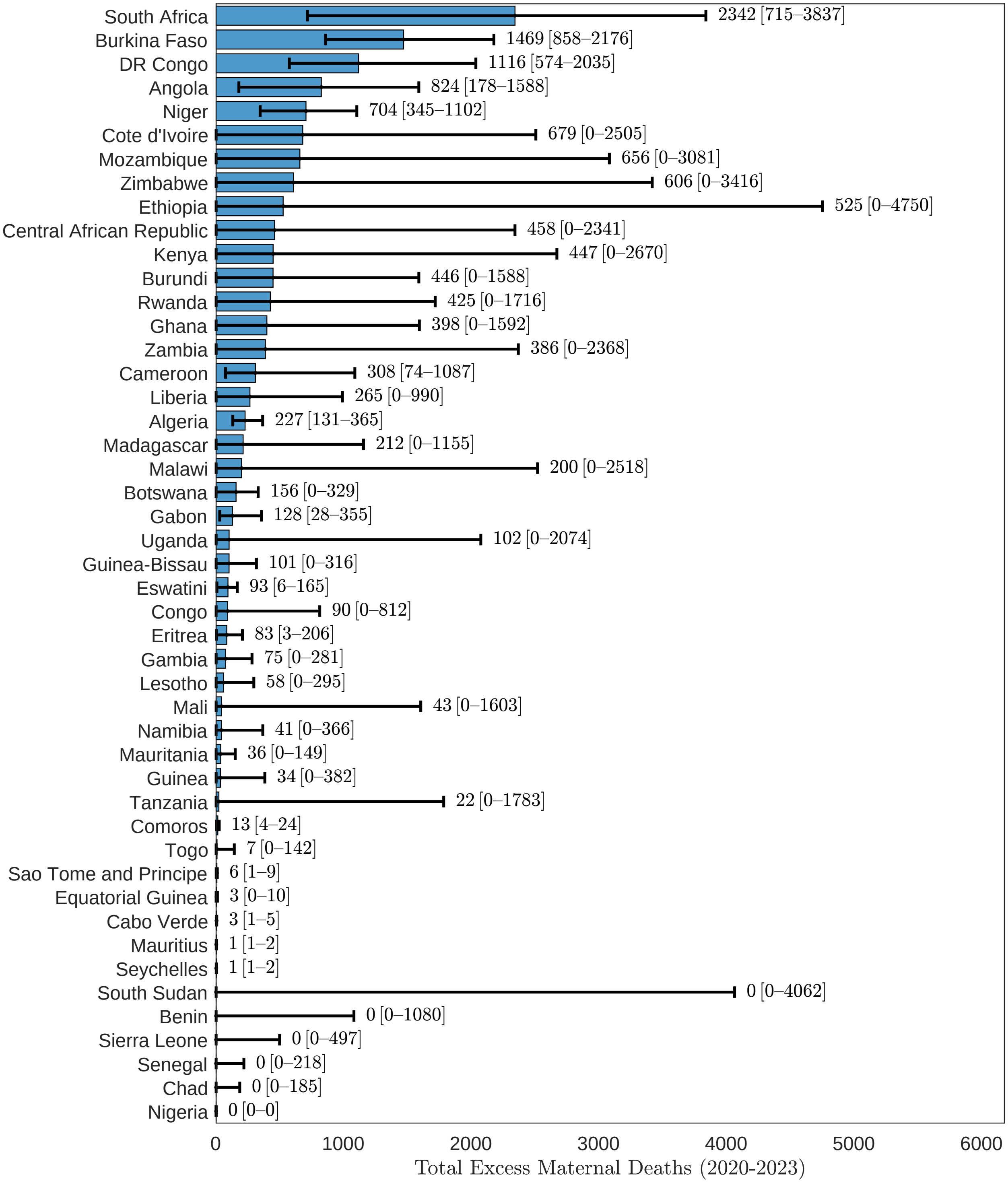


eFigure 45: Excess maternal deaths, African Region, 2020-2023.

##### 3.4.2 MMR

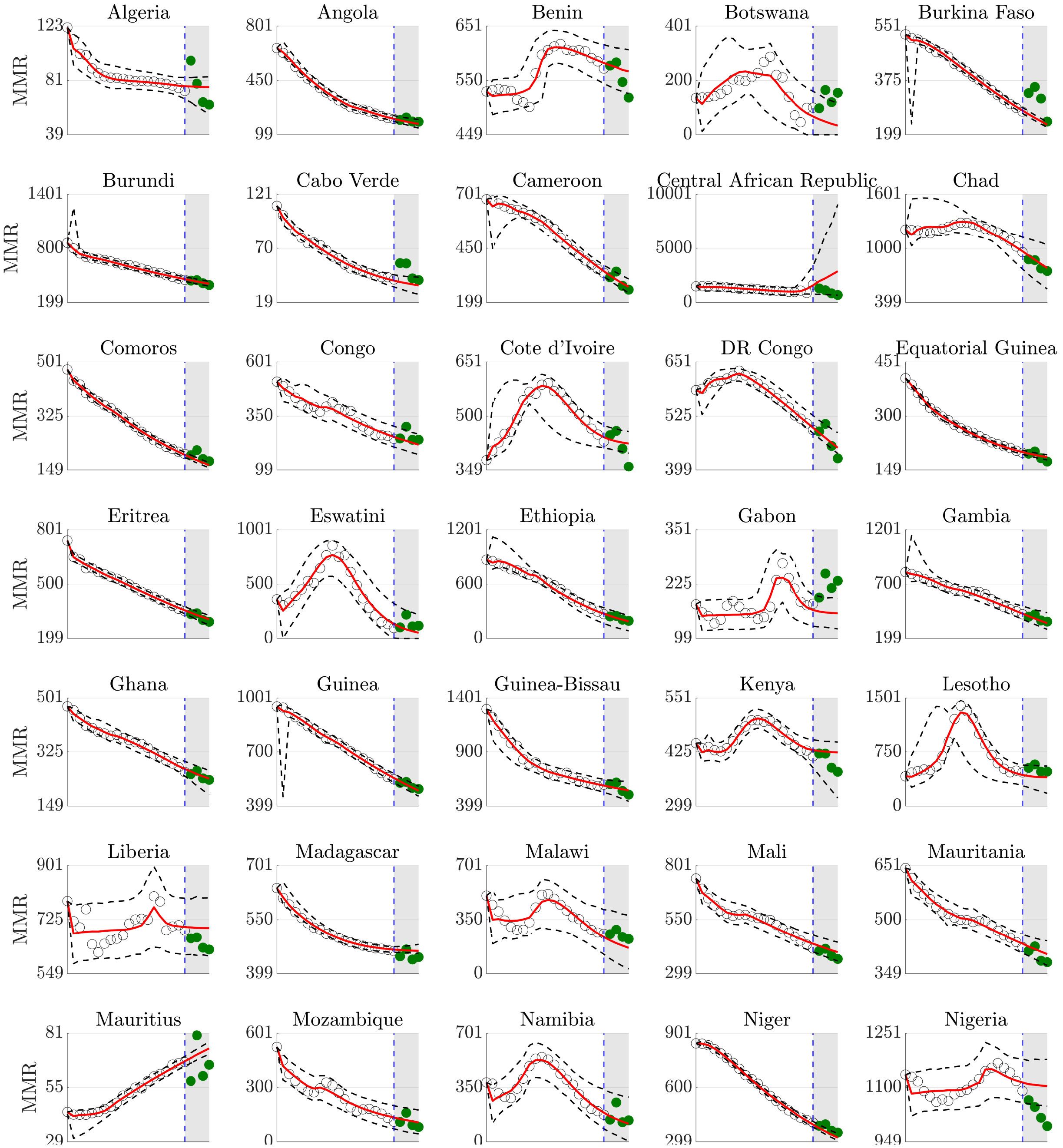


eFigure 46: Counterfactual maternal mortality ratio forecasts, African Region.

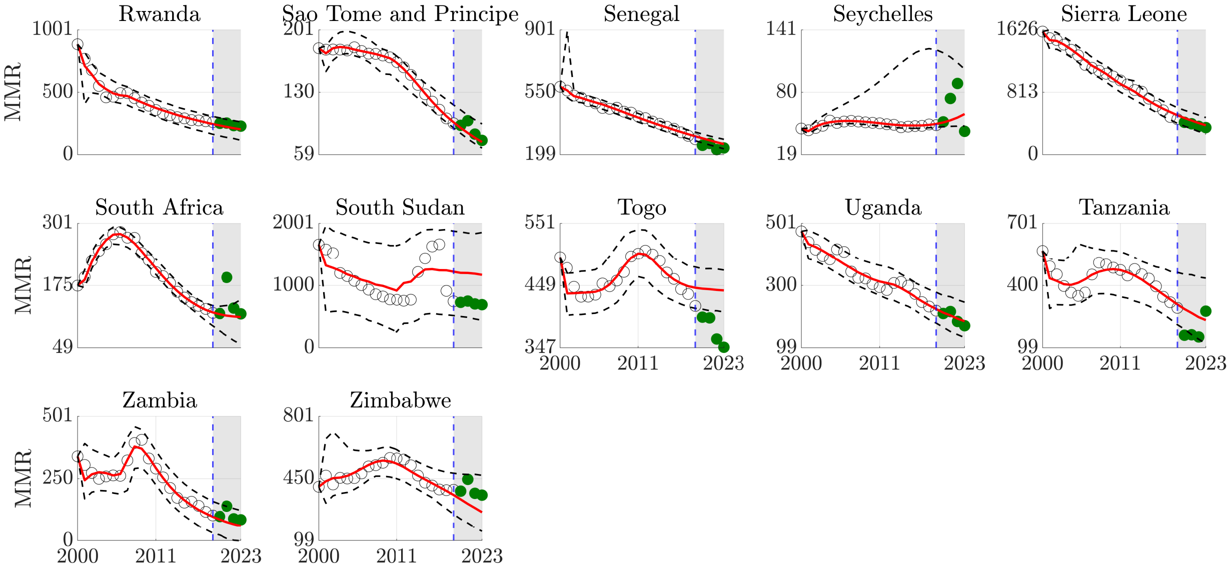


Counterfactual maternal mortality ratio forecasts, African Region (continued).

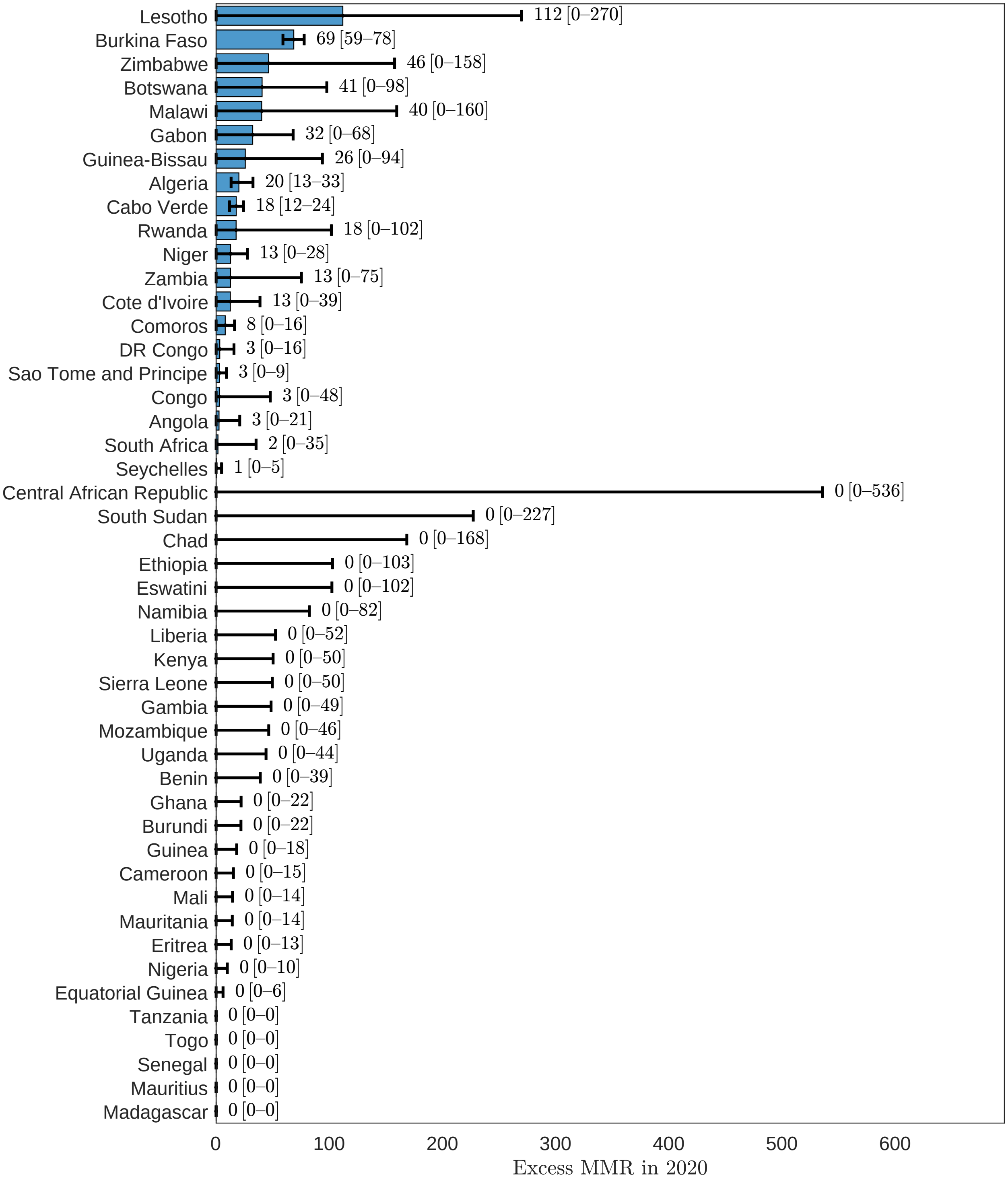


eFigure 47: Excess maternal mortality ratio, African Region, 2020.

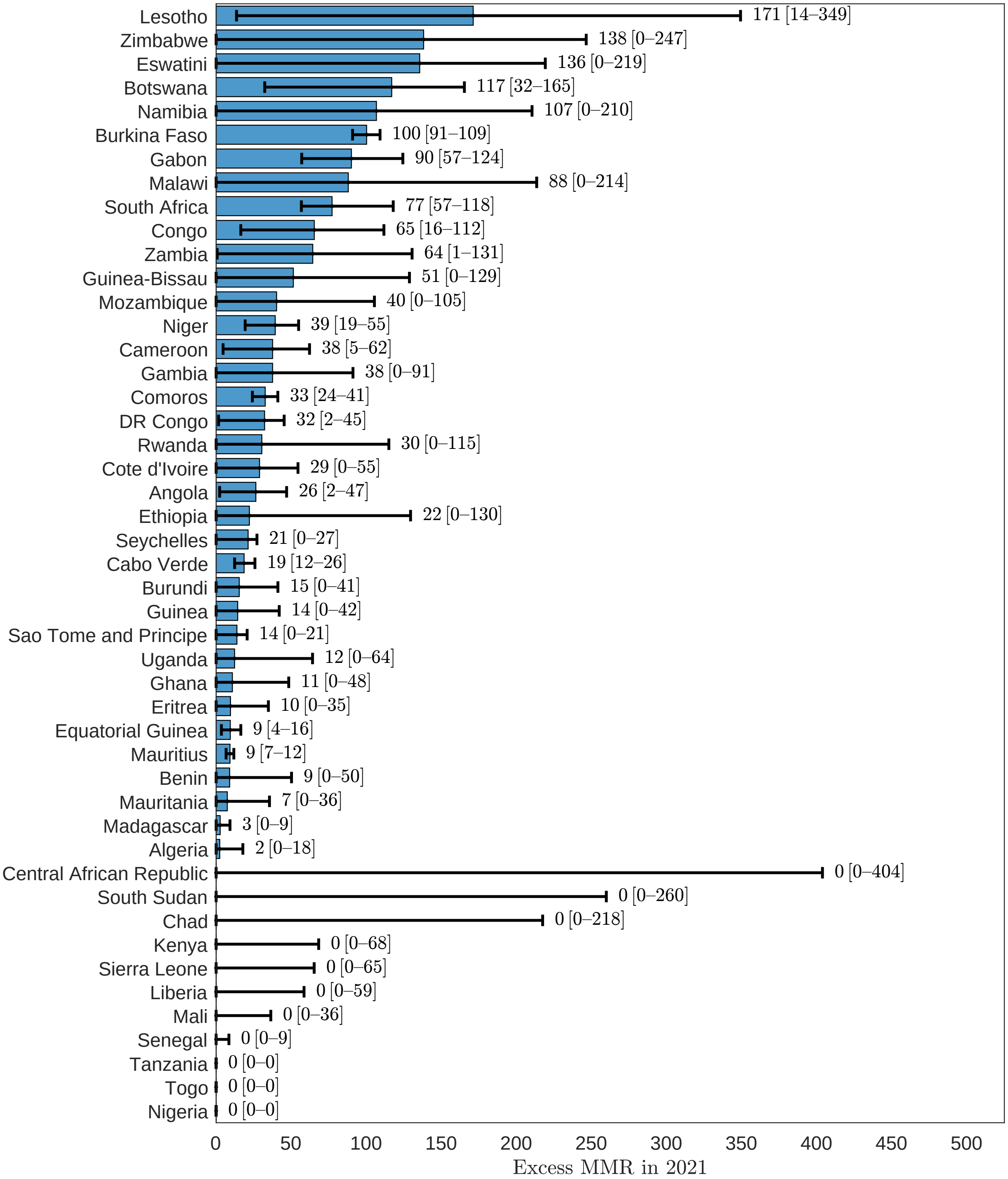


eFigure 48: Excess maternal mortality ratio, African Region, 2021.

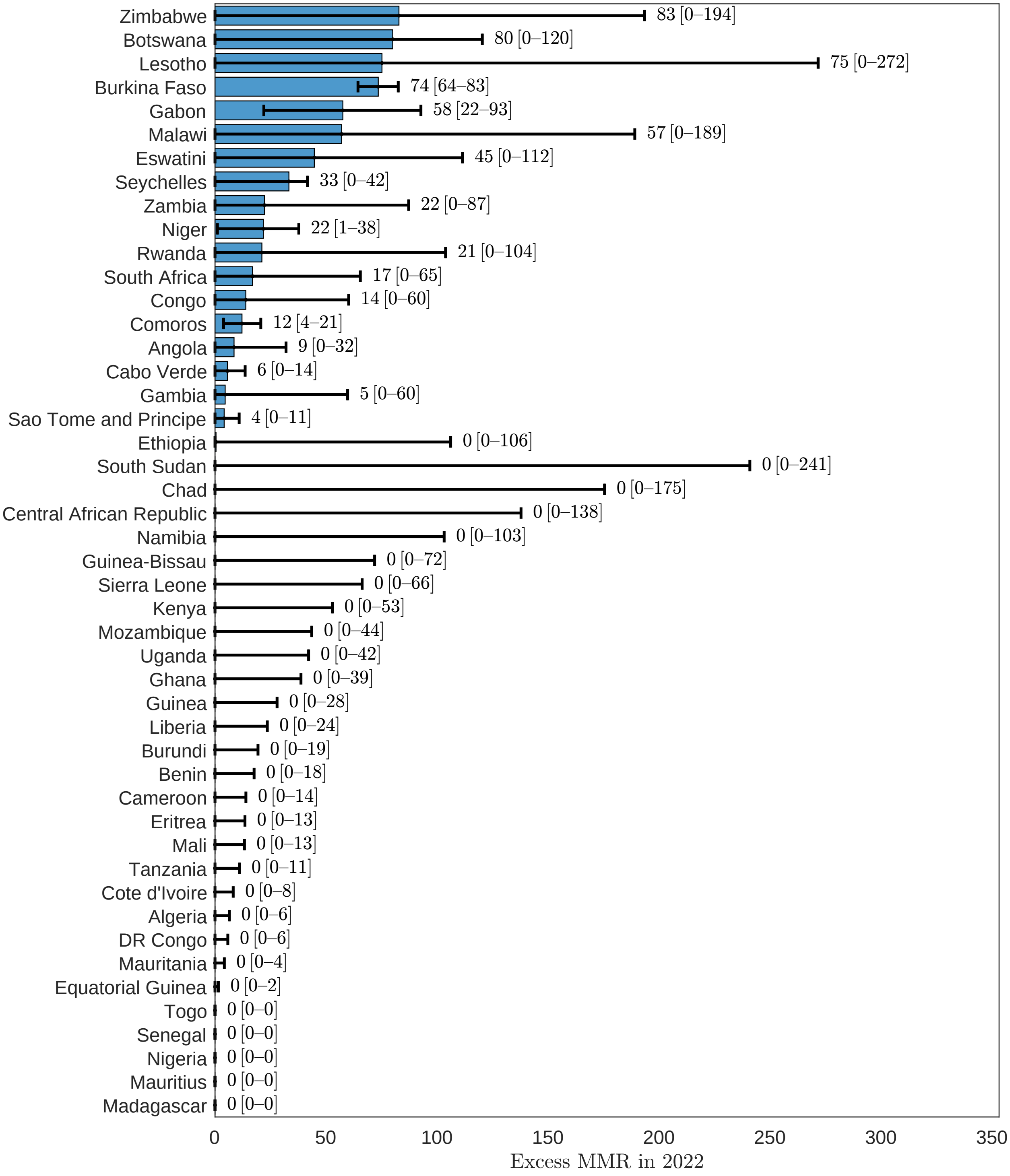


eFigure 49: Excess maternal mortality ratio, African Region, 2022.

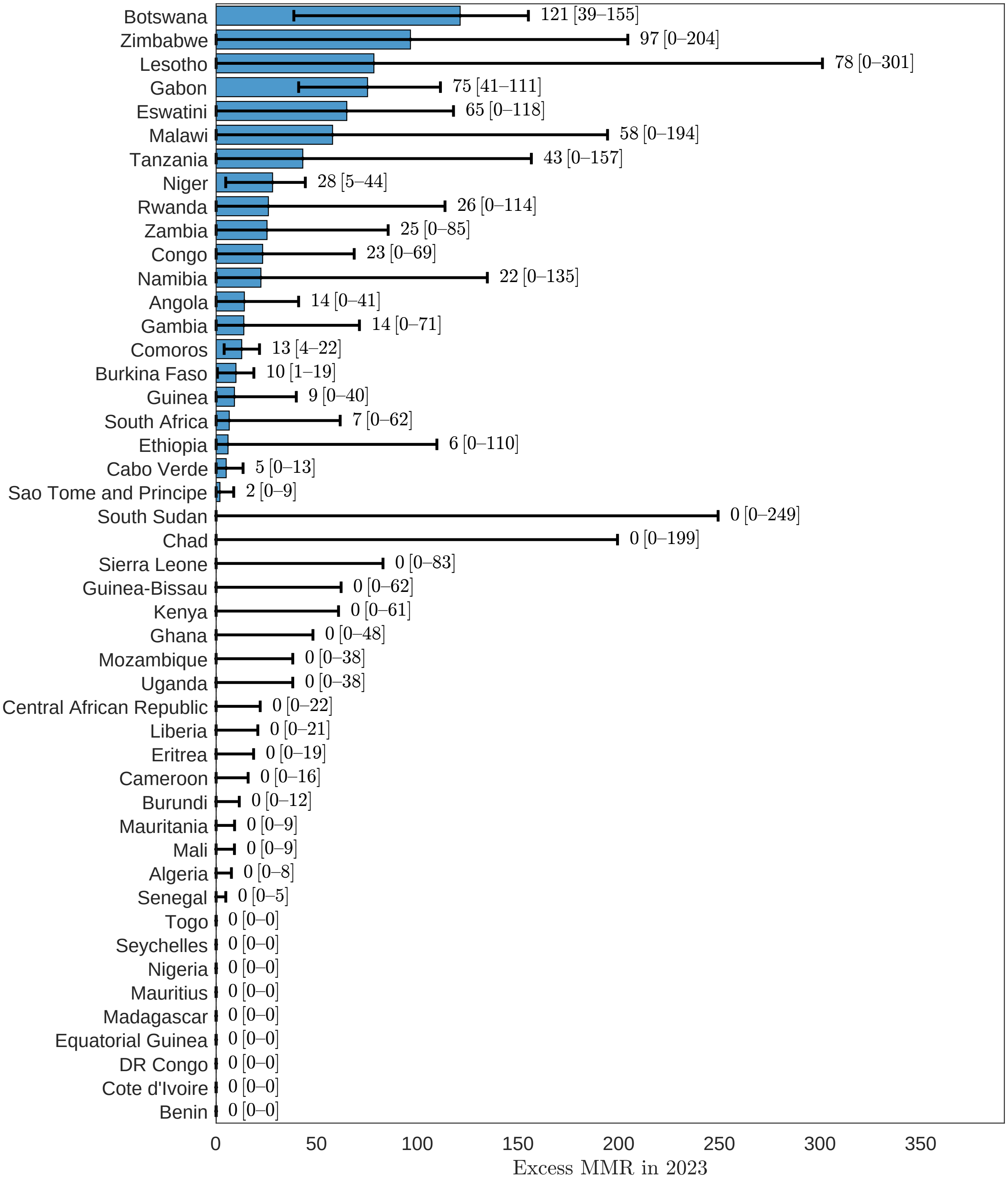


eFigure 50: Excess maternal mortality ratio, African Region, 2023.

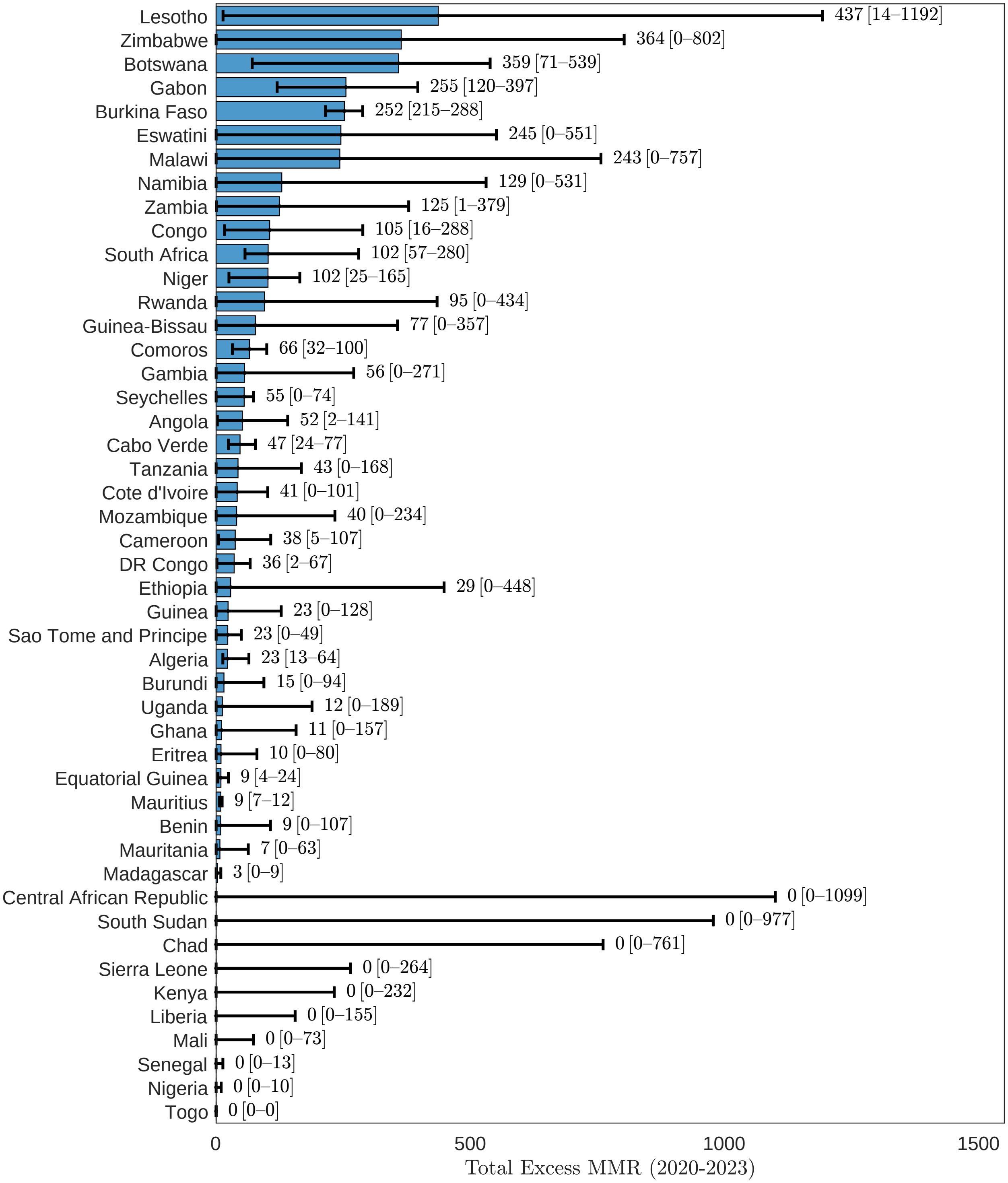


eFigure 51: Excess maternal mortality ratio, African Region, 2020-2023.

#### 3.5 Western Pacific Region

##### 3.5.1 Maternal deaths

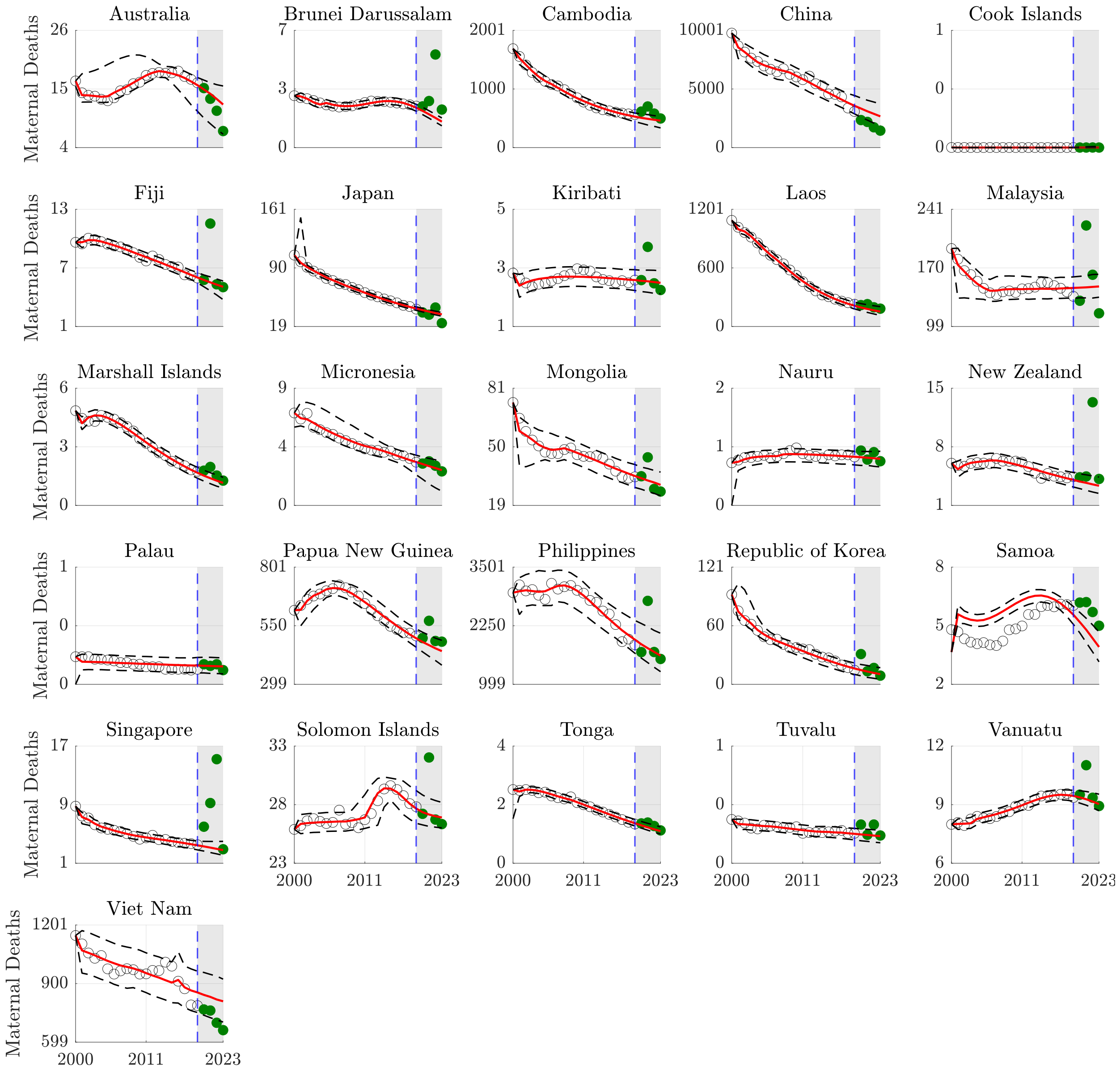


eFigure 52: Counterfactual maternal death forecasts, Western Pacific Region.

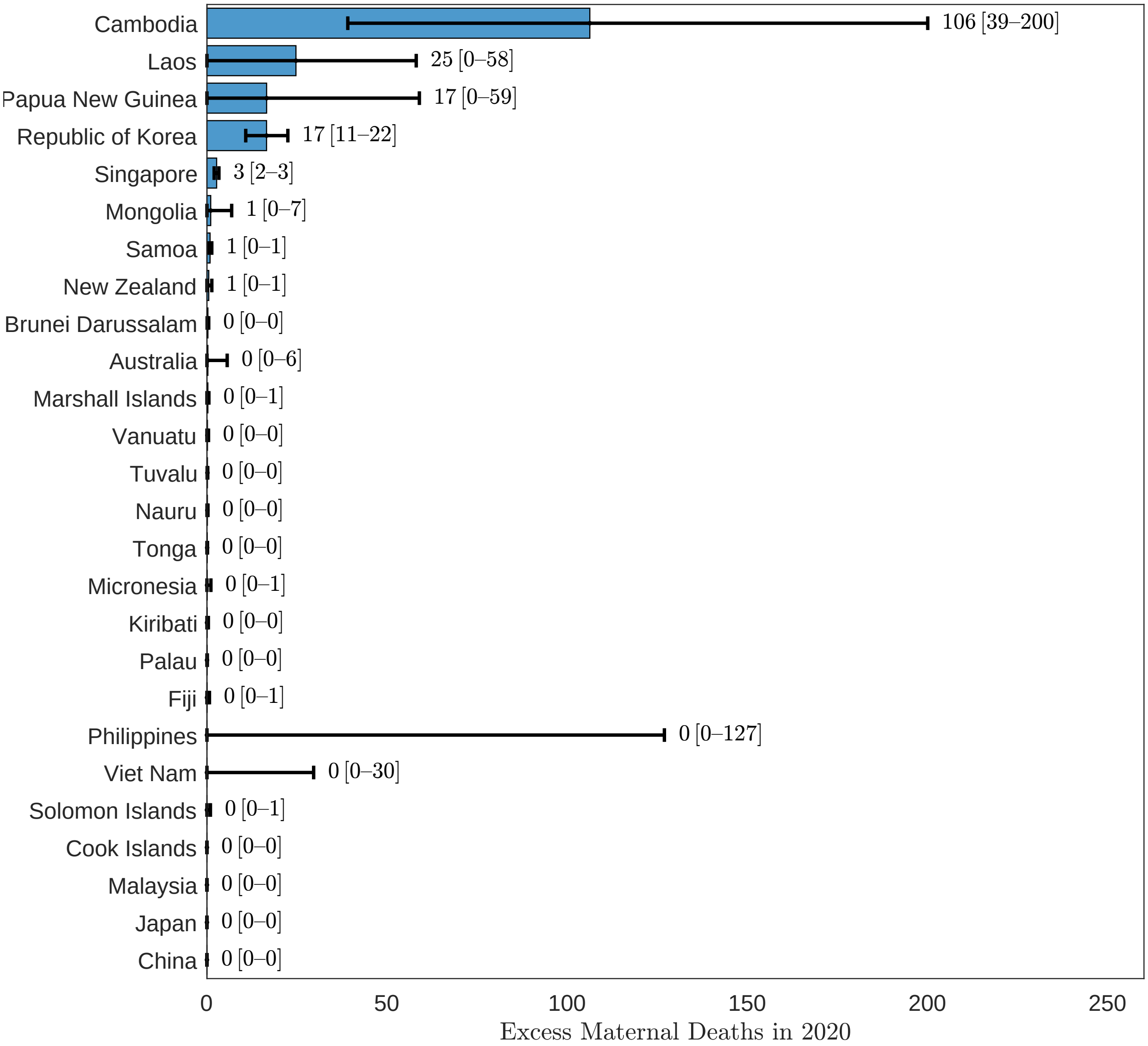


eFigure 53: Excess maternal deaths, Western Pacific Region, 2020.

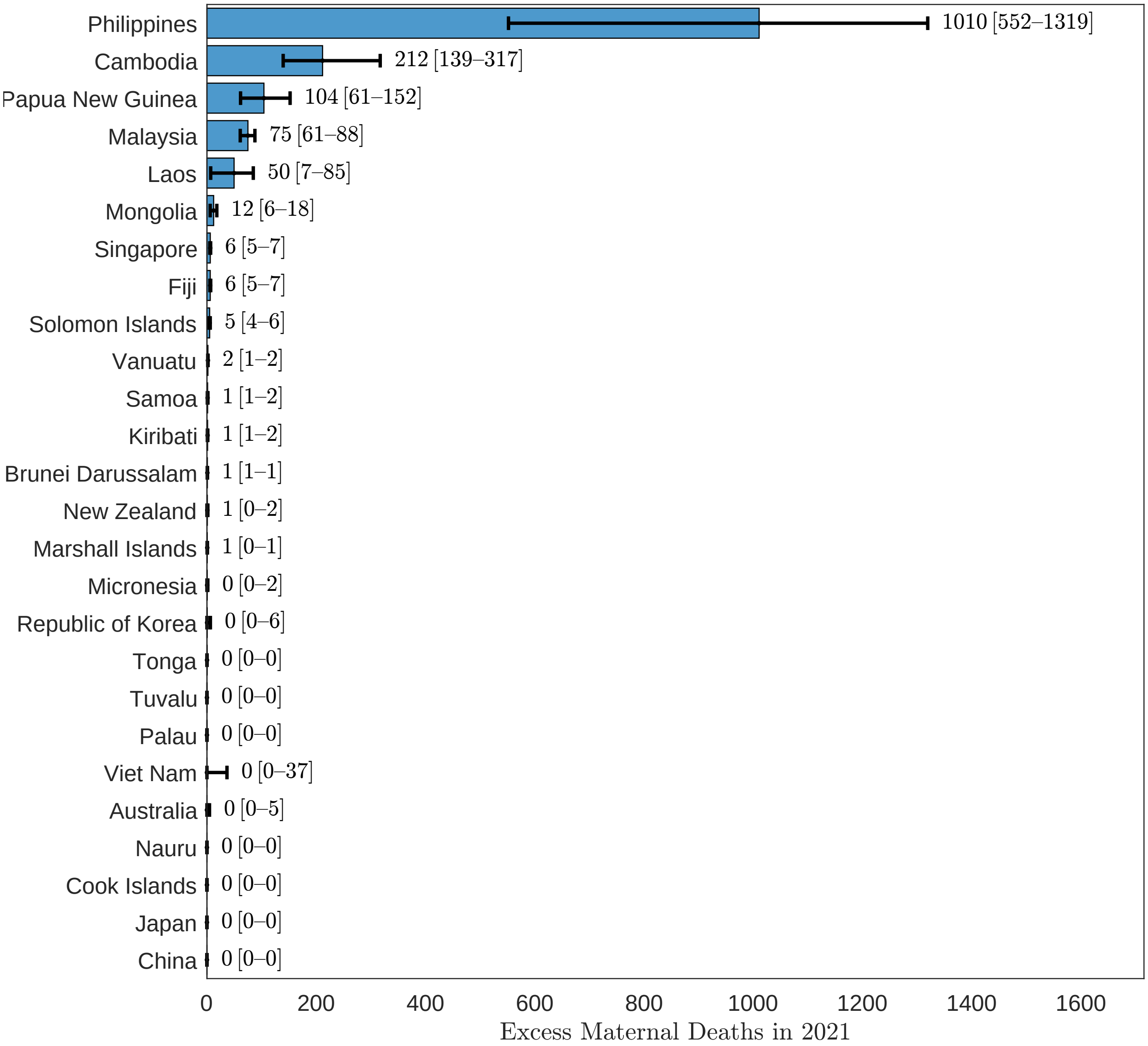


eFigure 54: Excess maternal deaths, Western Pacific Region, 2021.

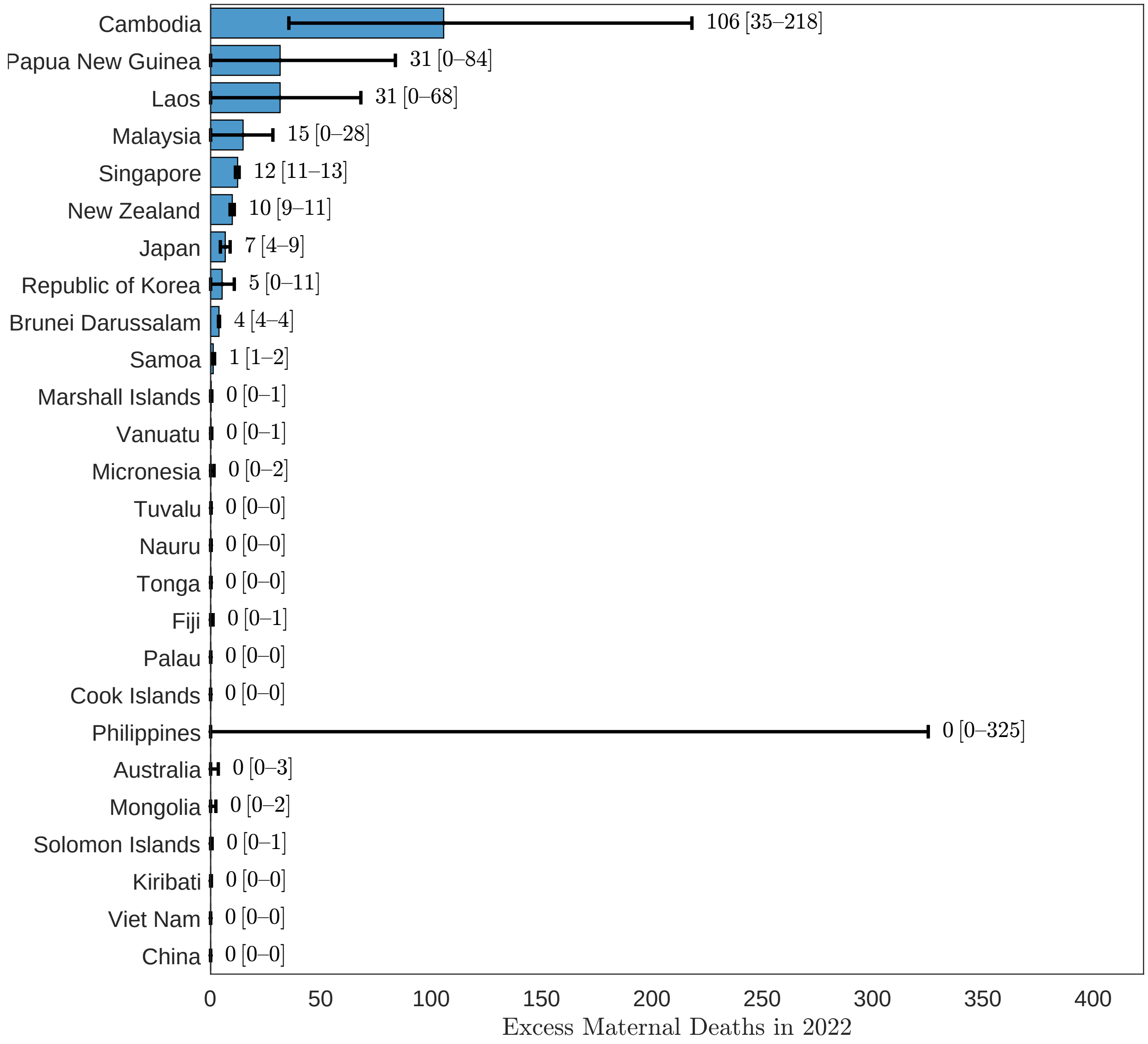


eFigure 55: Excess maternal deaths, Western Pacific Region, 2022.

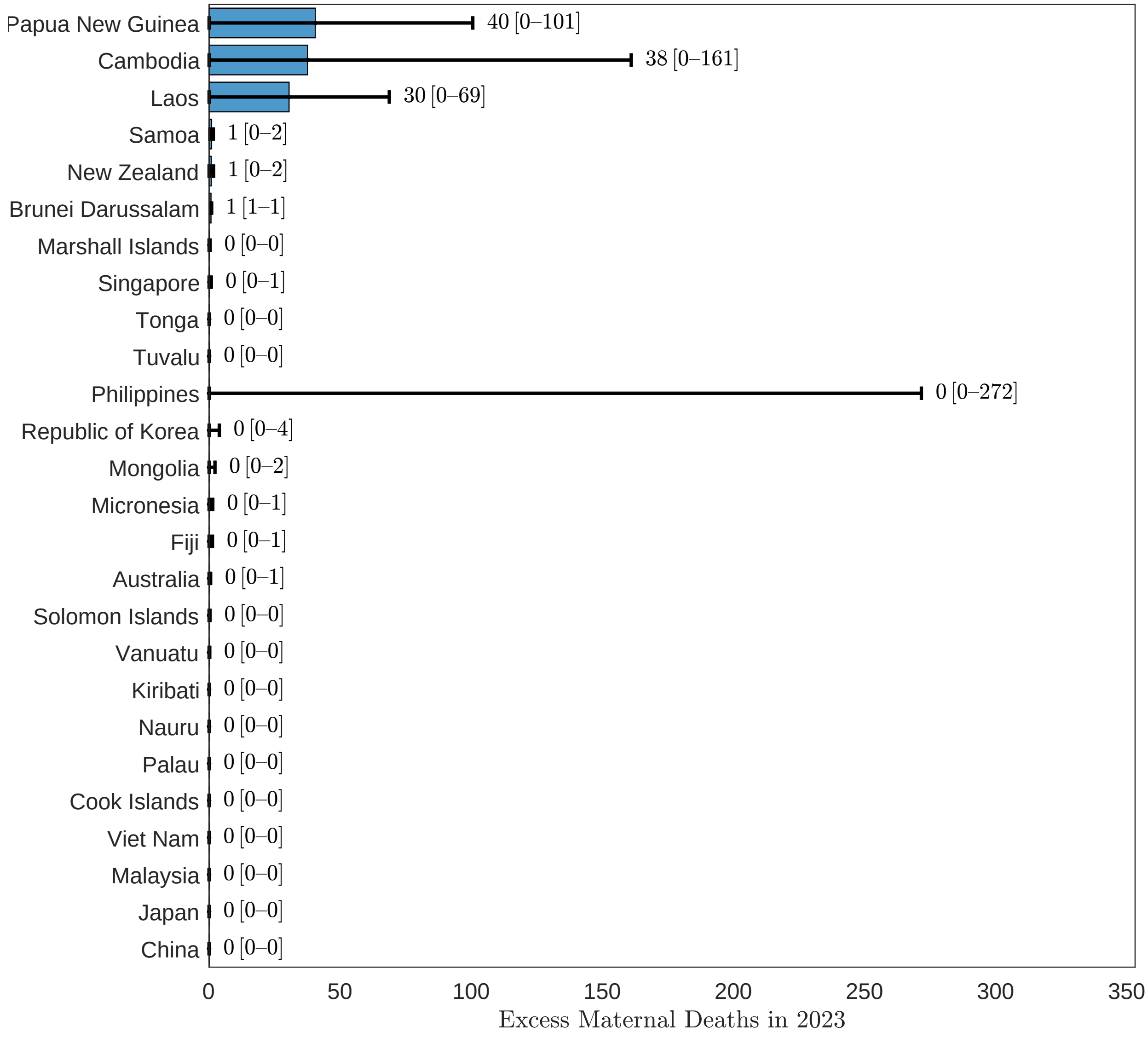


eFigure 56: Excess maternal deaths, Western Pacific Region, 2023.

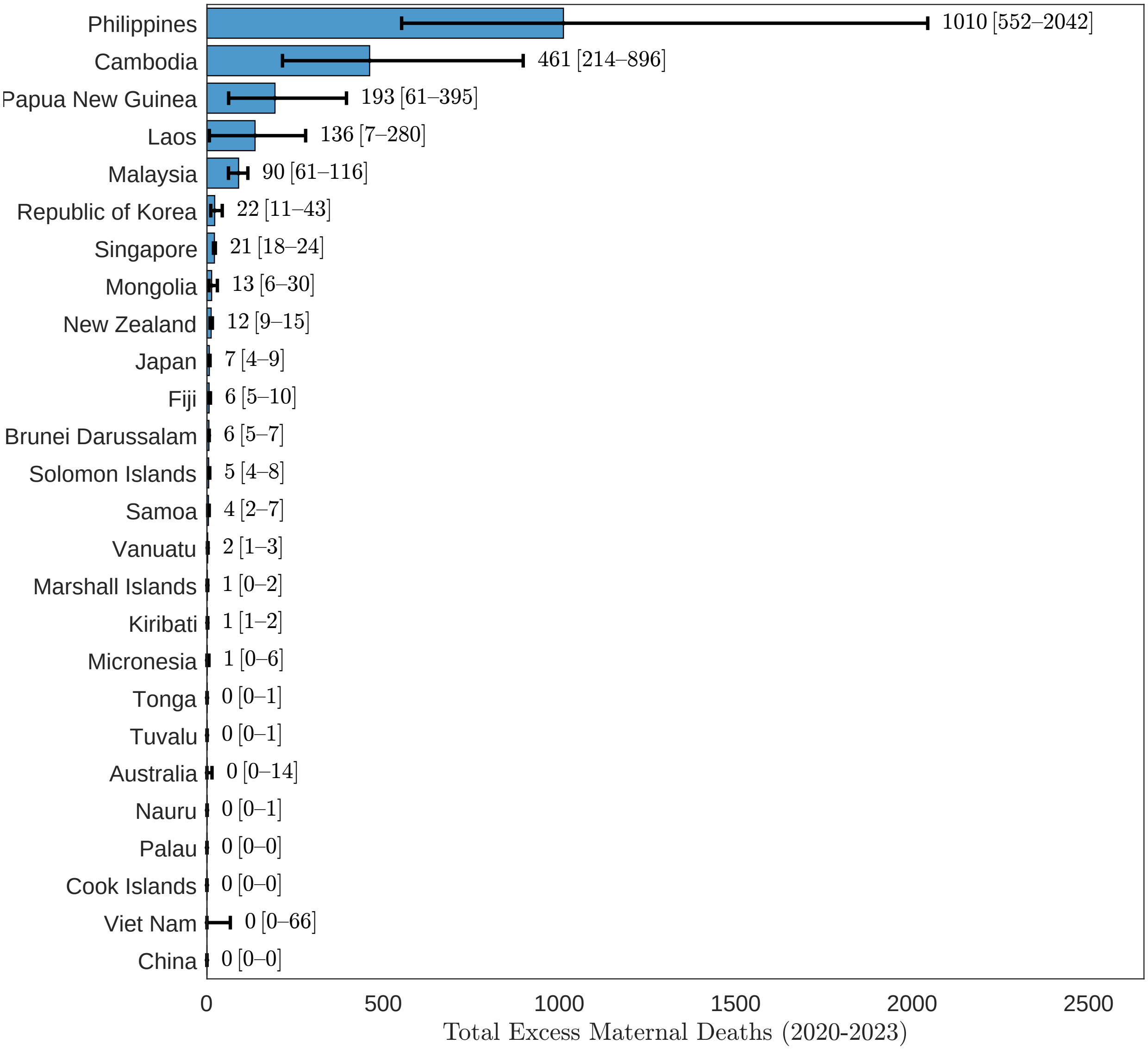


eFigure 57: Excess maternal deaths, Western Pacific Region, 2020-2023.

##### 3.5.2 MMR

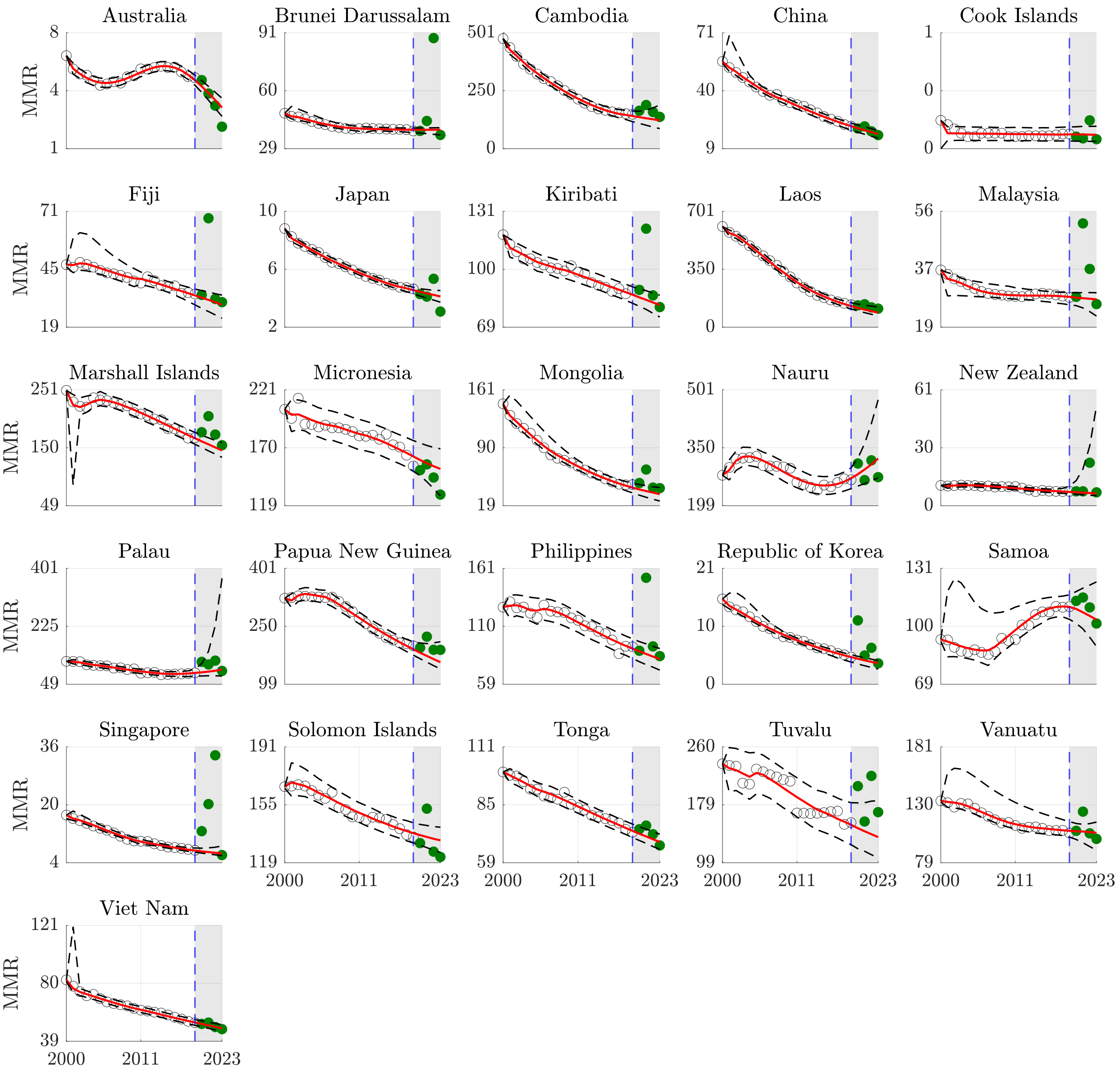


eFigure 58: Counterfactual maternal mortality ratio forecasts, Western Pacific Region.

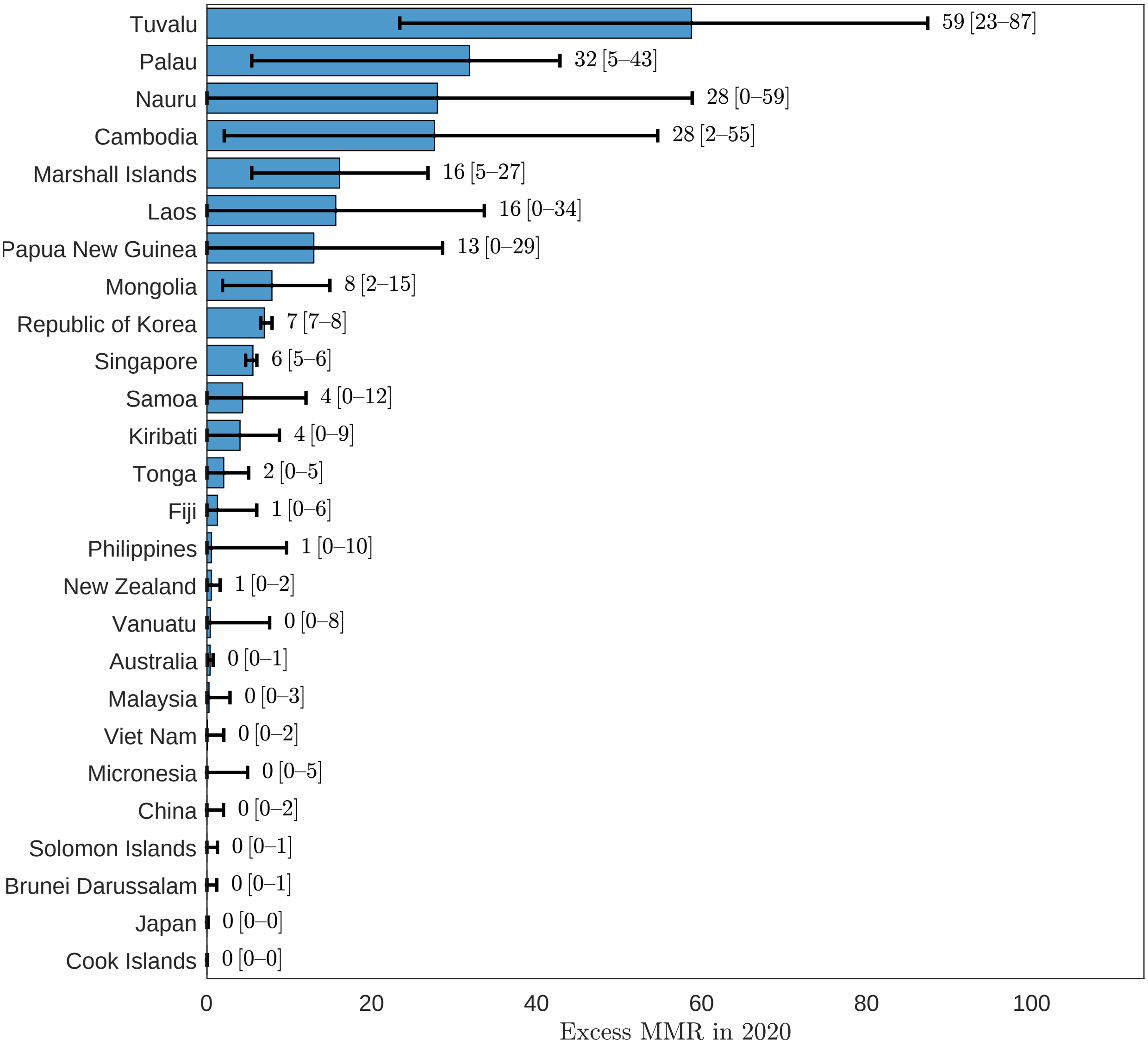


eFigure 59: Excess maternal mortality ratio, Western Pacific Region, 2020.

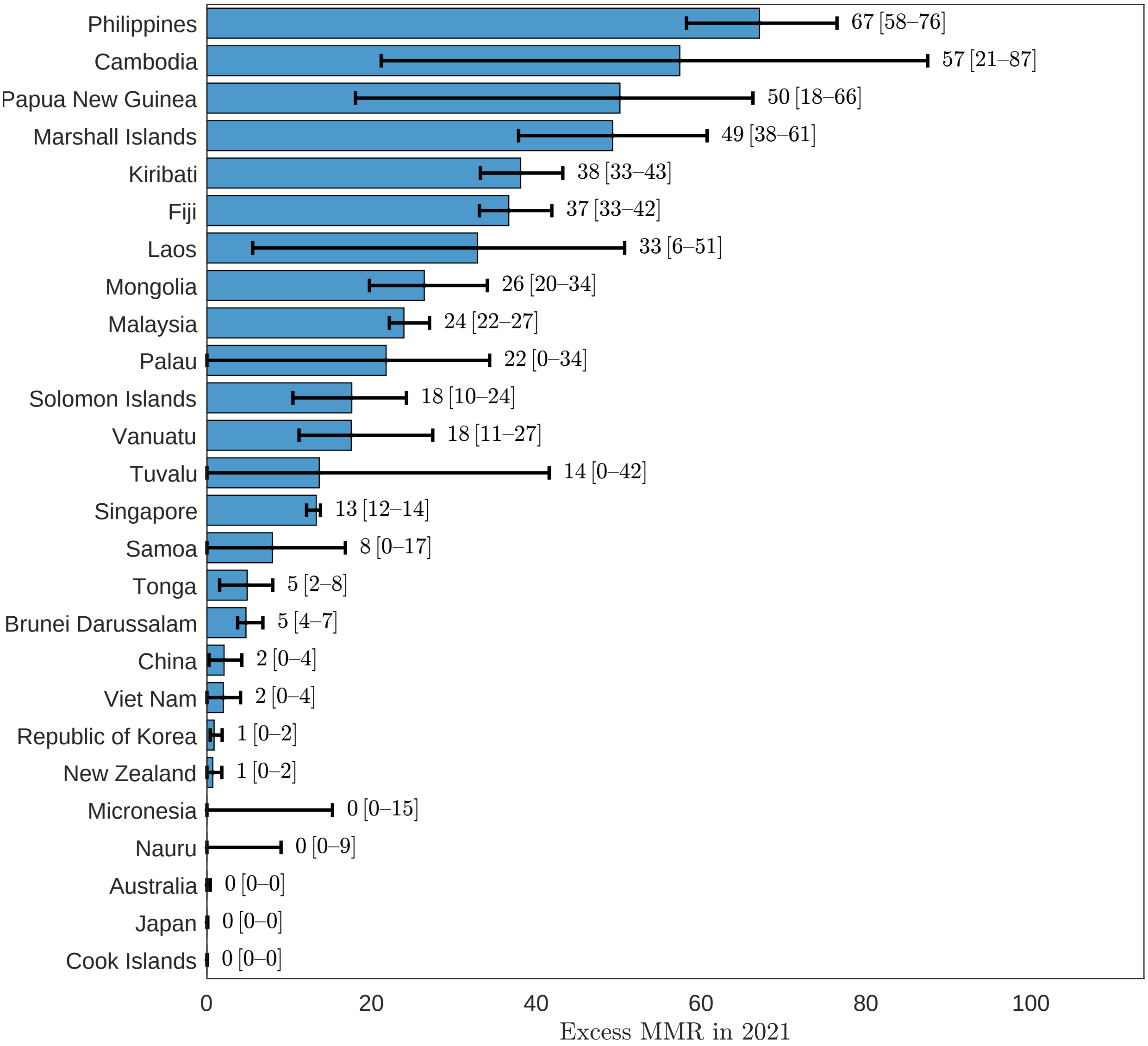


eFigure 60: Excess maternal mortality ratio, Western Pacific Region, 2021.

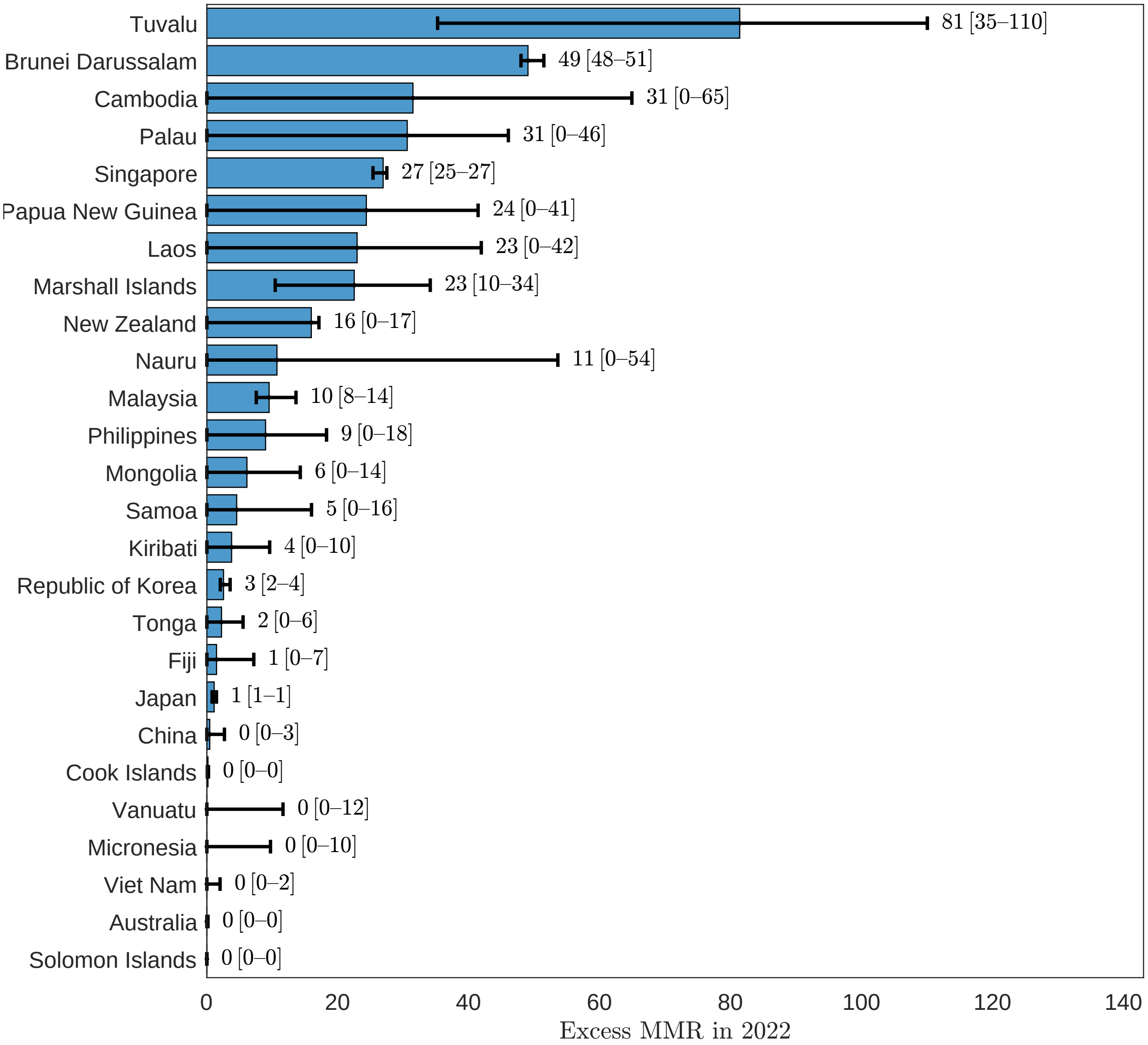


eFigure 61: Excess maternal mortality ratio, Western Pacific Region, 2022.

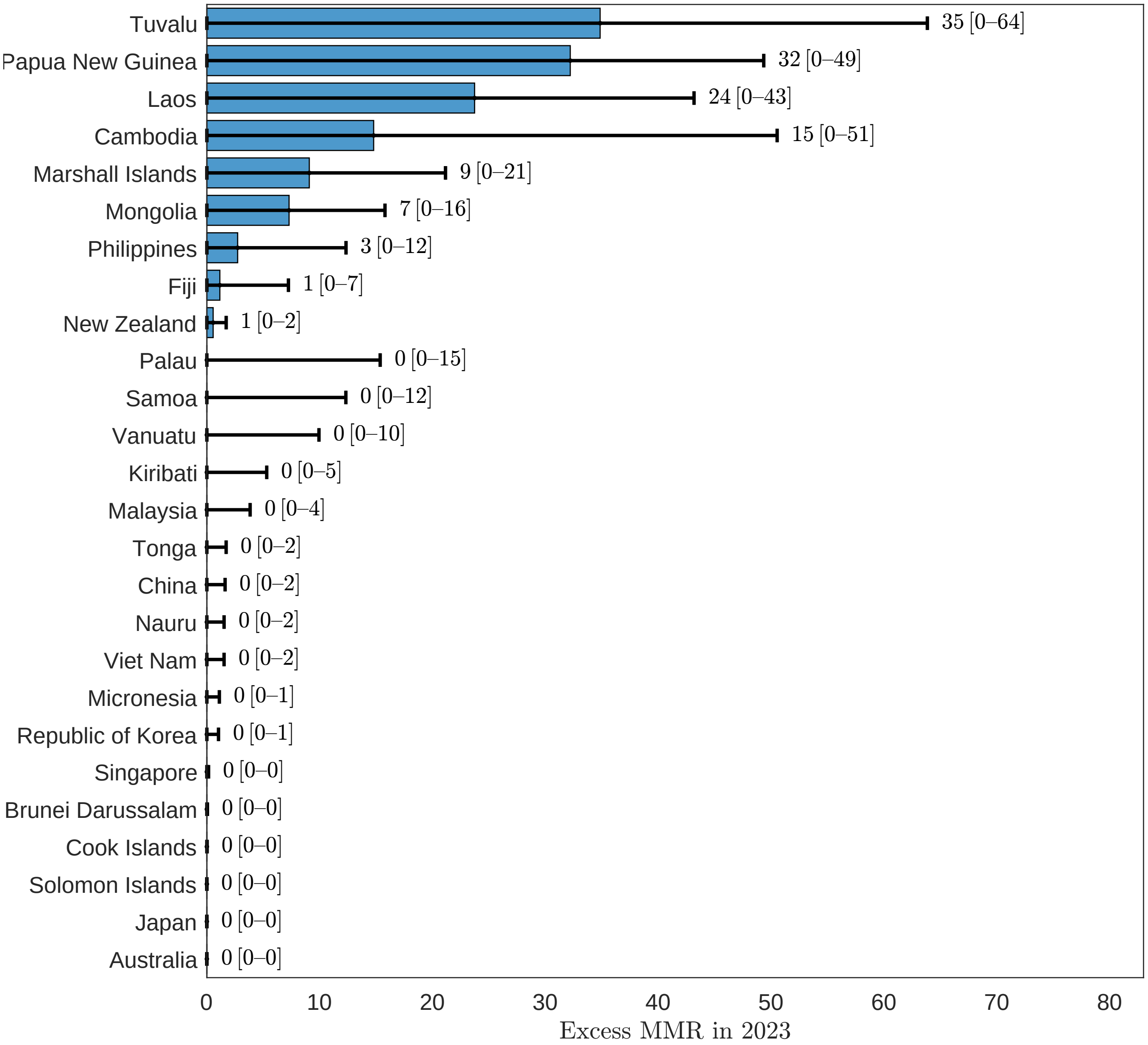


eFigure 62: Excess maternal mortality ratio, Western Pacific Region, 2023.

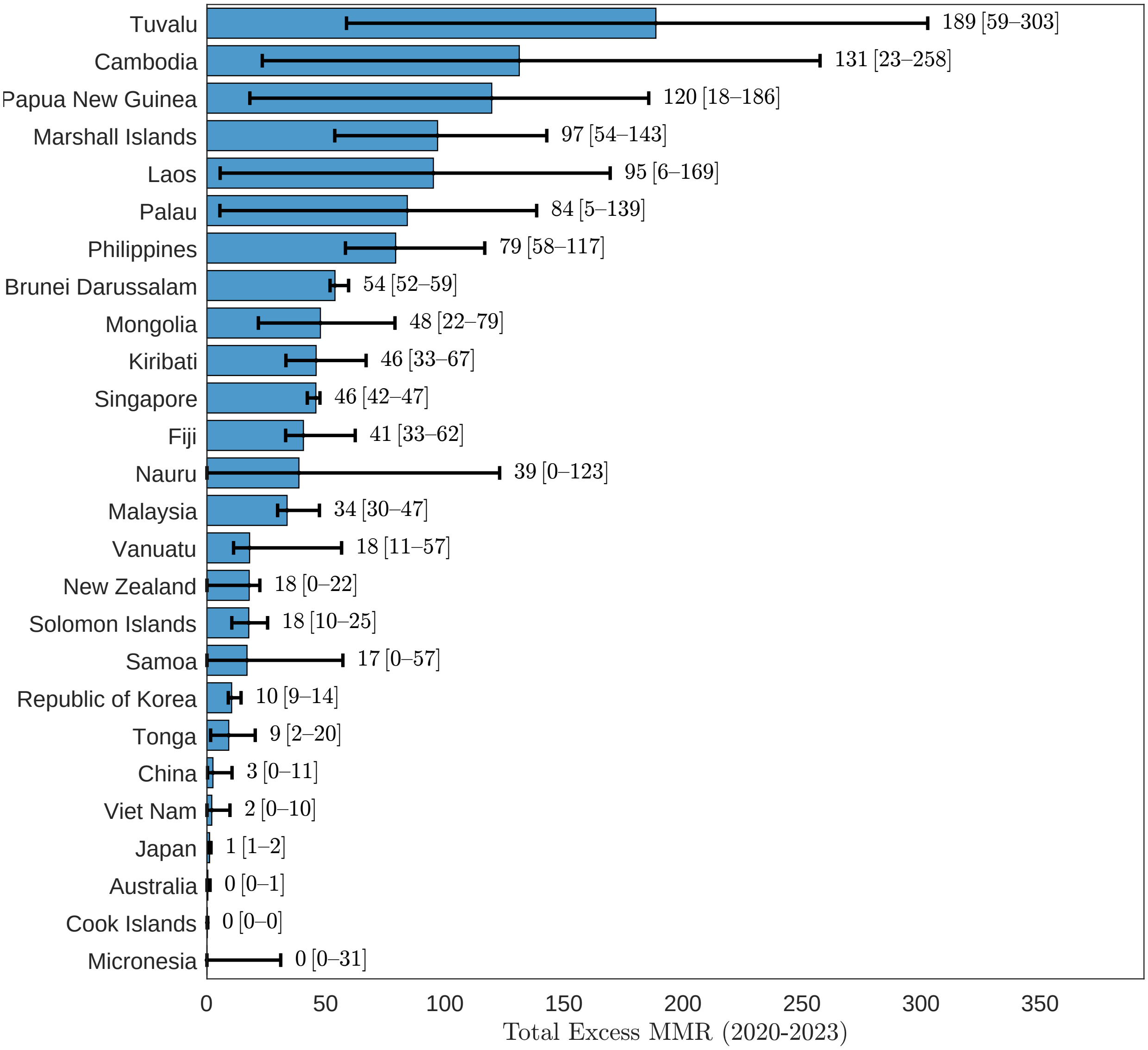


eFigure 63: Excess maternal mortality ratio, Western Pacific Region, 2020-2023.

#### 3.6 Region of the Americas

##### 3.6.1 Maternal deaths

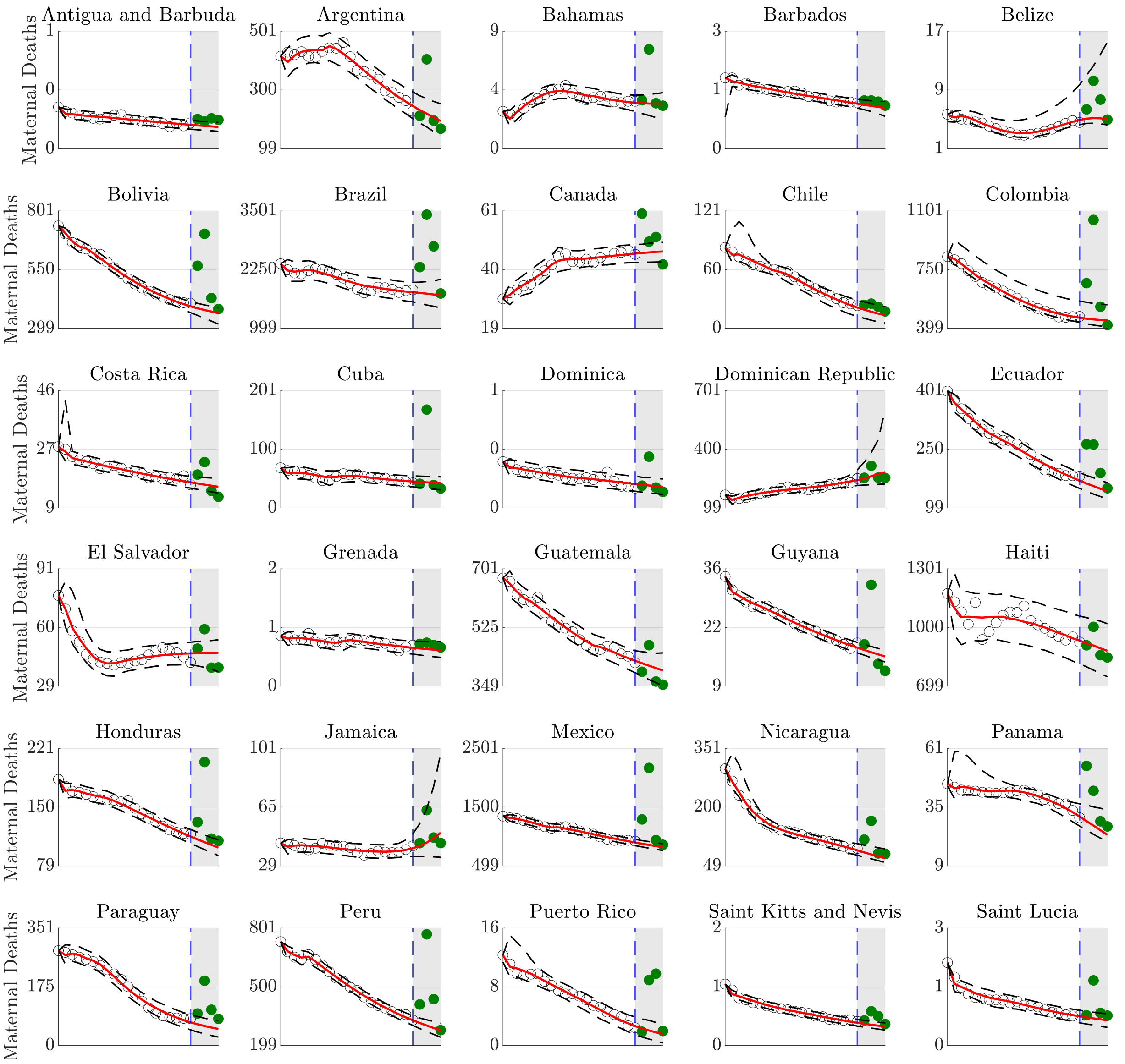


eFigure 64: Counterfactual maternal death forecasts, Region of the Americas.

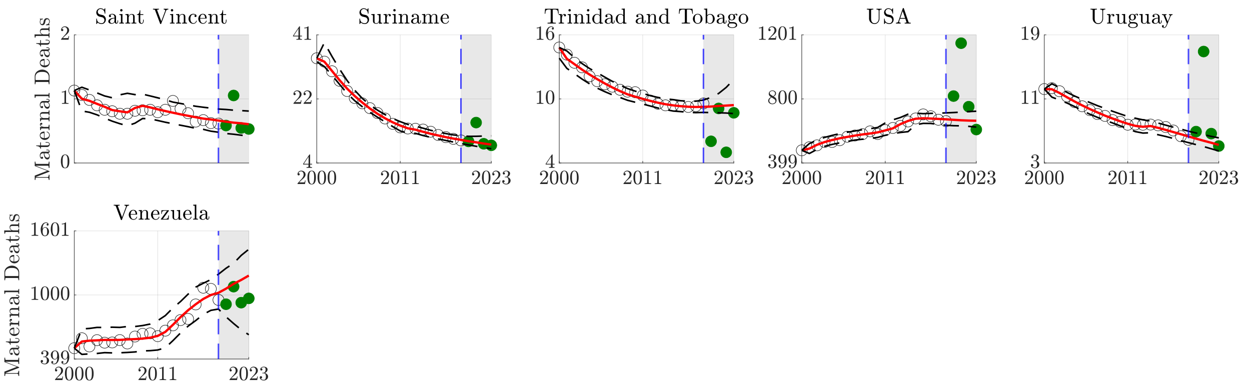


Counterfactual maternal death forecasts, Region of the Americas (continued).

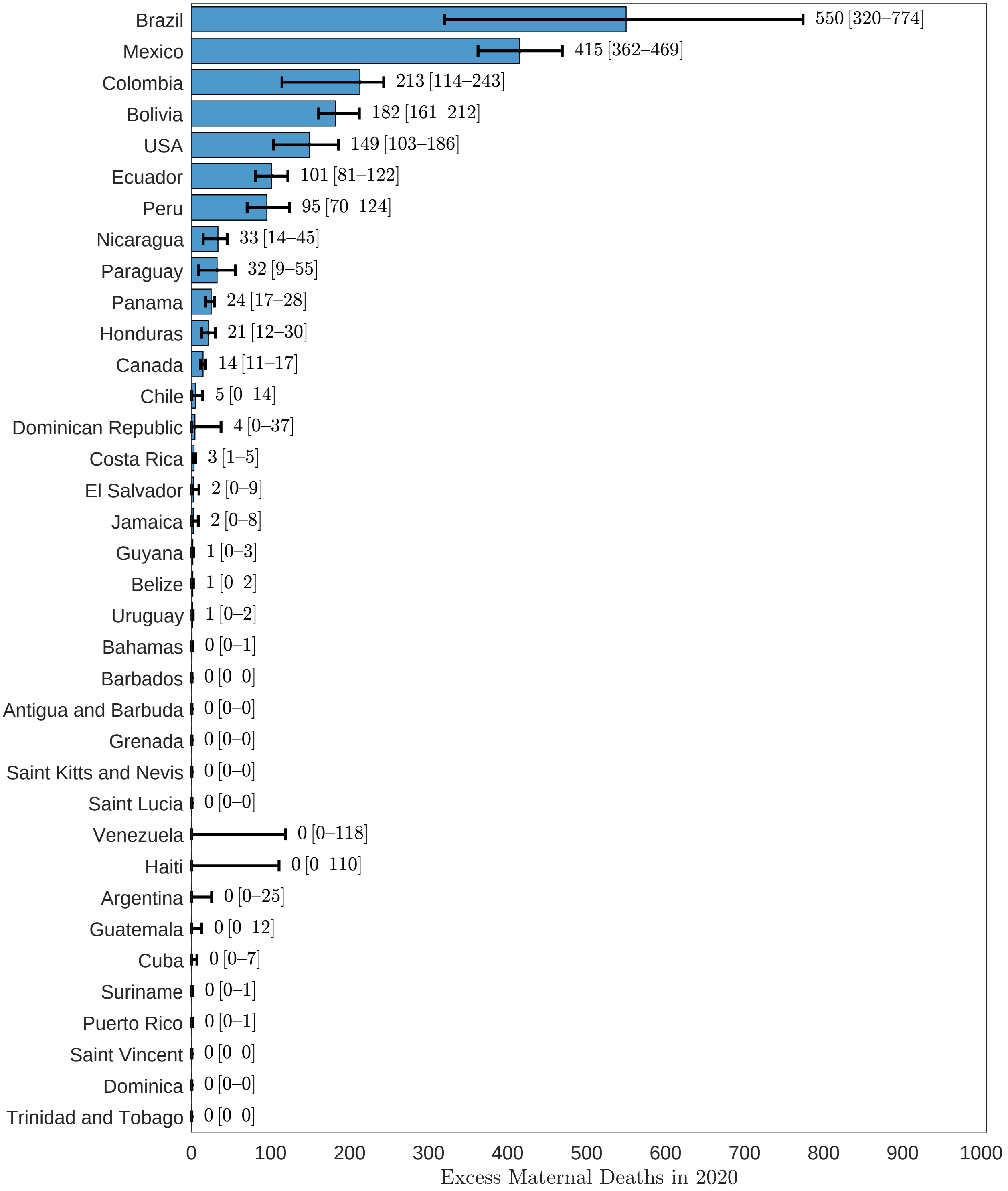


eFigure 65: Excess maternal deaths, Region of the Americas, 2020.

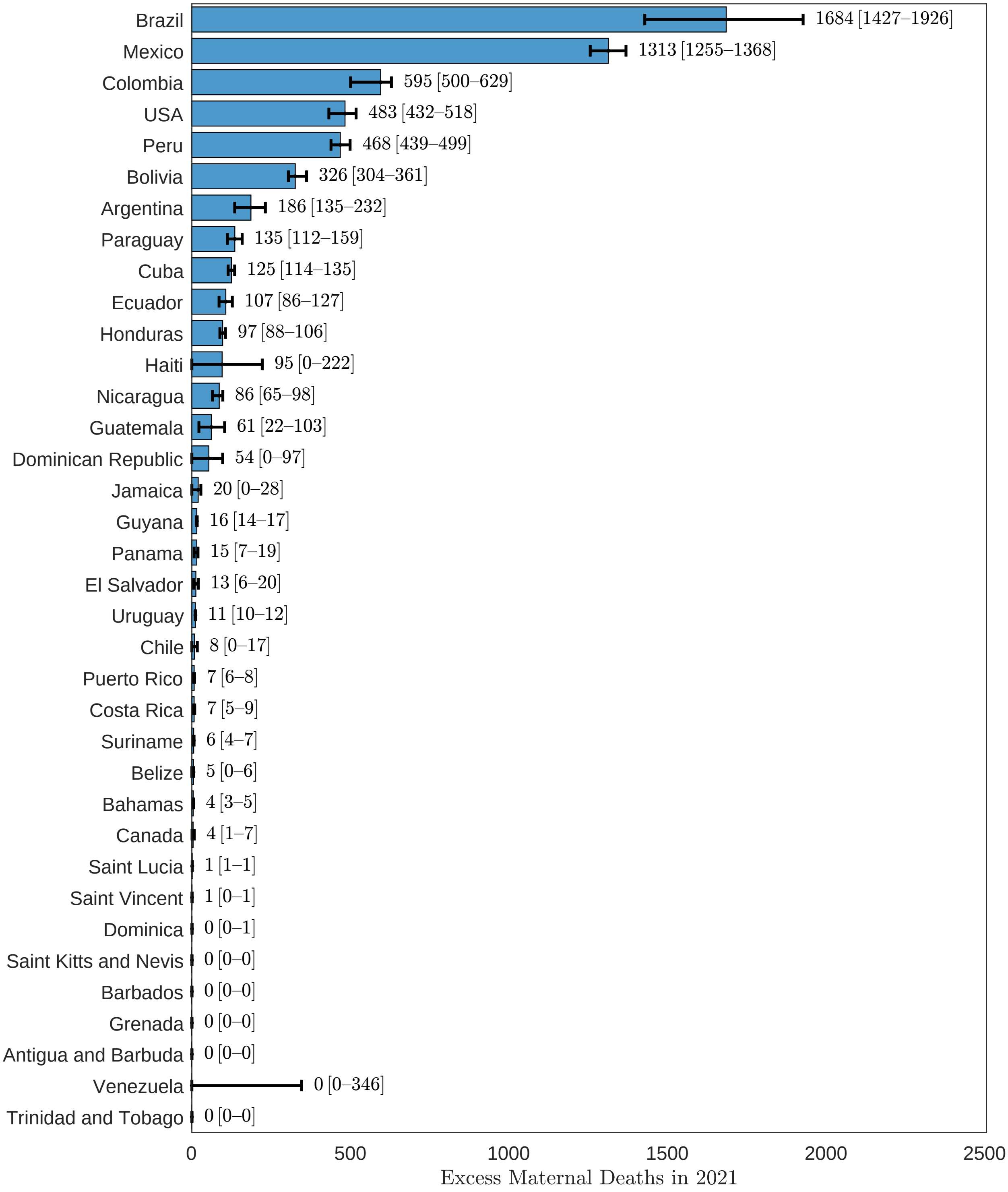


eFigure 66: Excess maternal deaths, Region of the Americas, 2021.

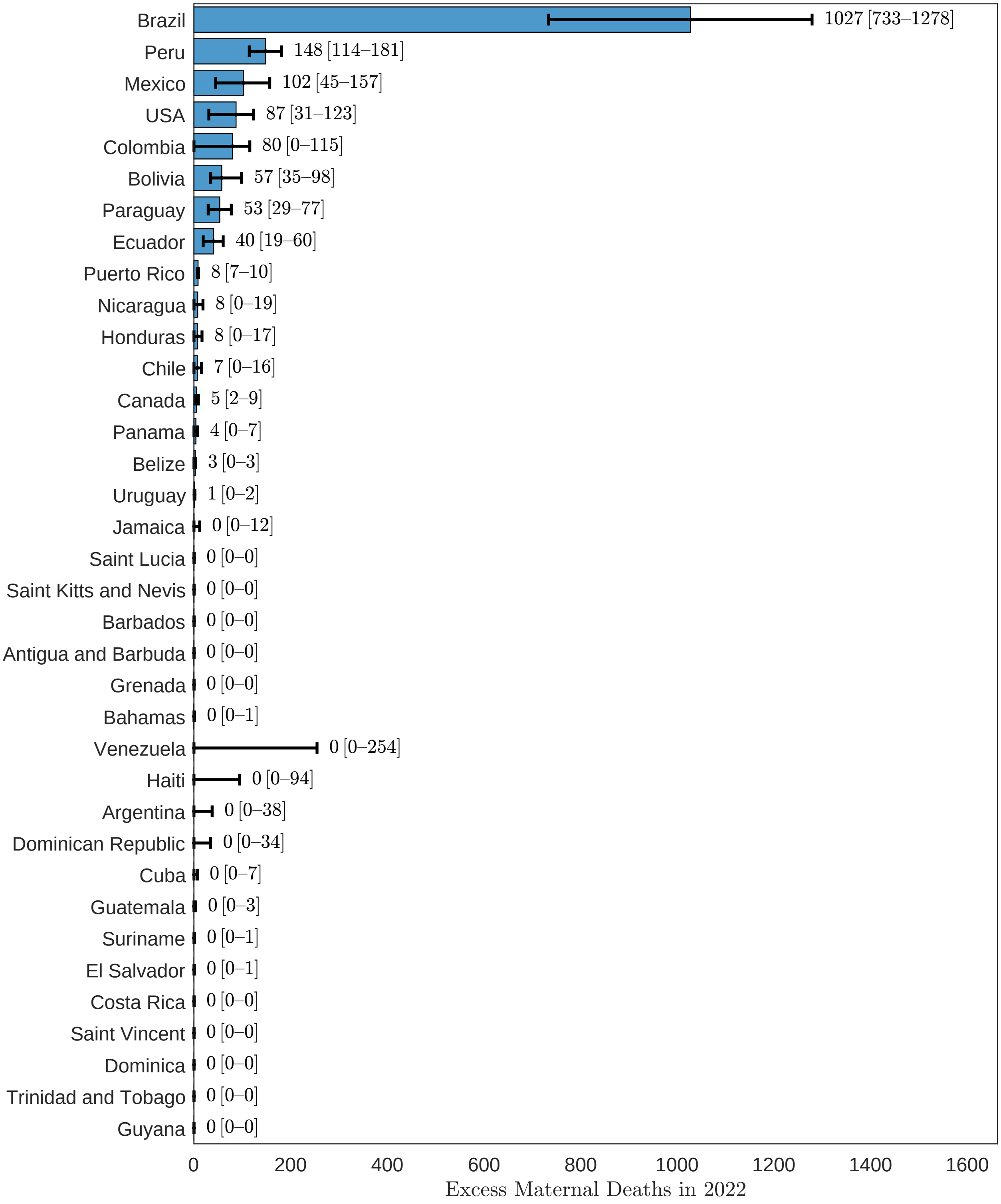


eFigure 67: Excess maternal deaths, Region of the Americas, 2022.

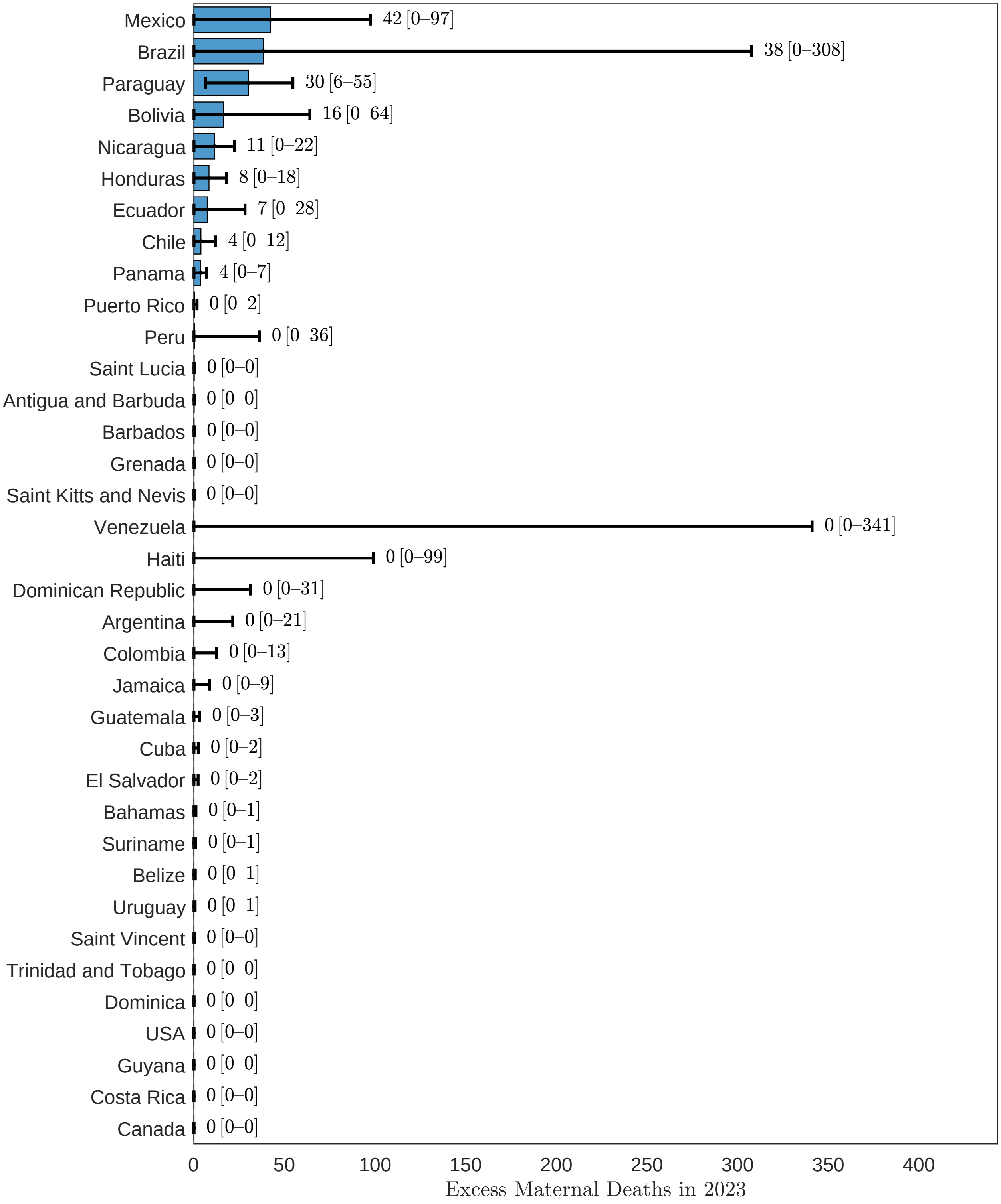


eFigure 68: Excess maternal deaths, Region of the Americas, 2023.

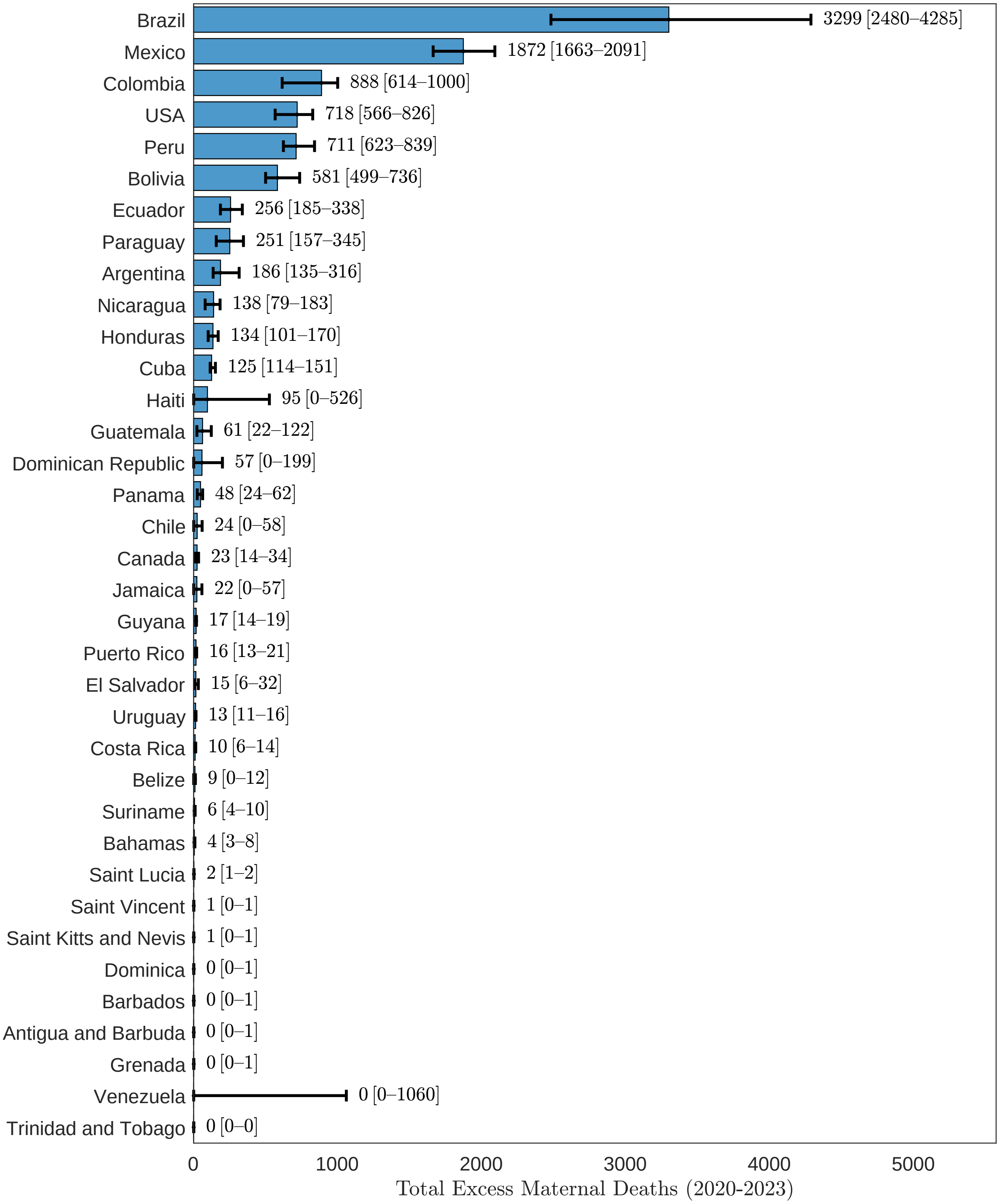


eFigure 69: Excess maternal deaths, Region of the Americas, 2020-2023.

##### 3.6.2 MMR

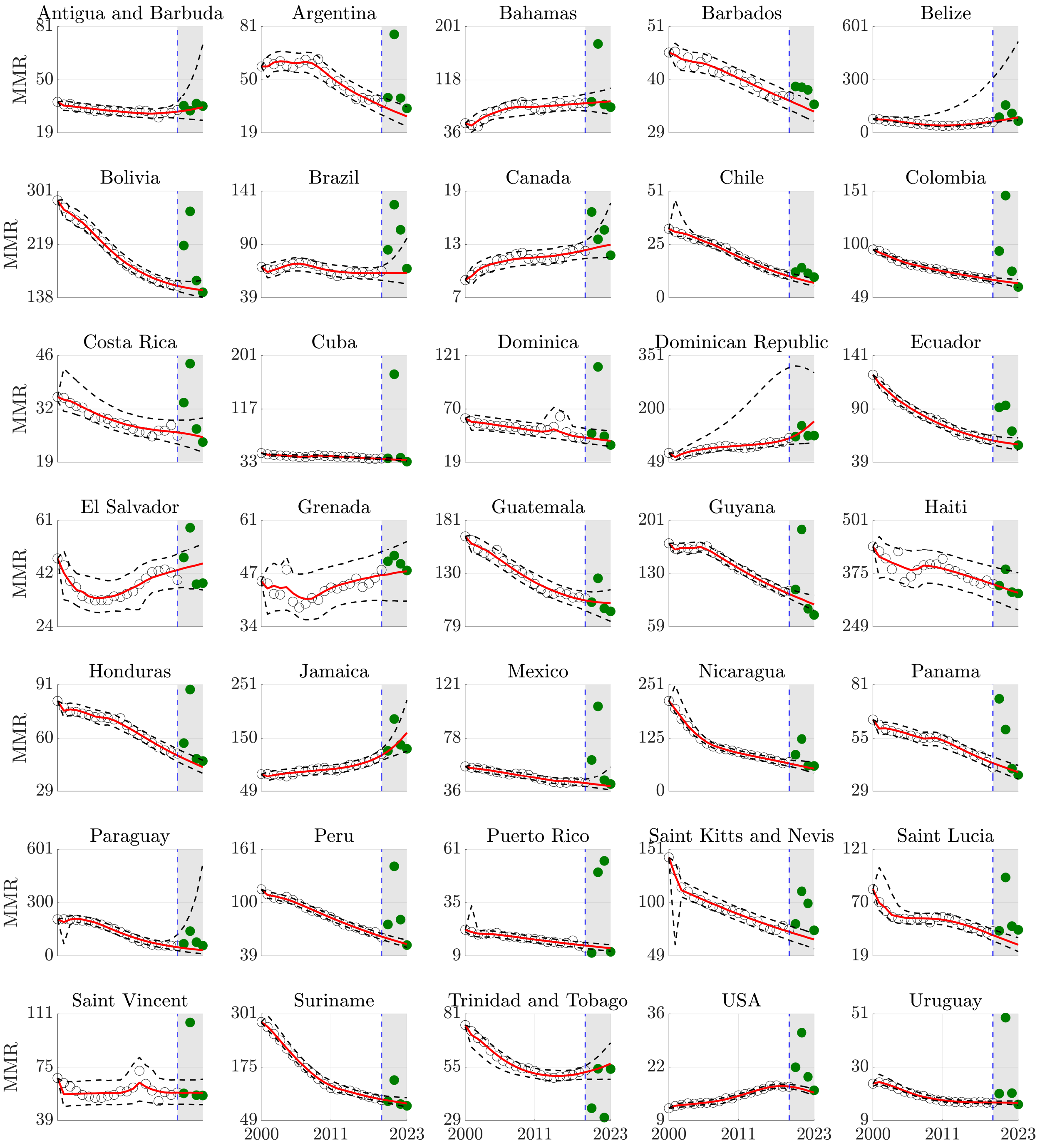


eFigure 70: Counterfactual maternal mortality ratio forecasts, Region of the Americas.

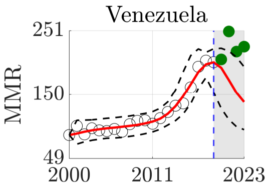


Counterfactual maternal mortality ratio forecasts, Region of the Americas (continued).

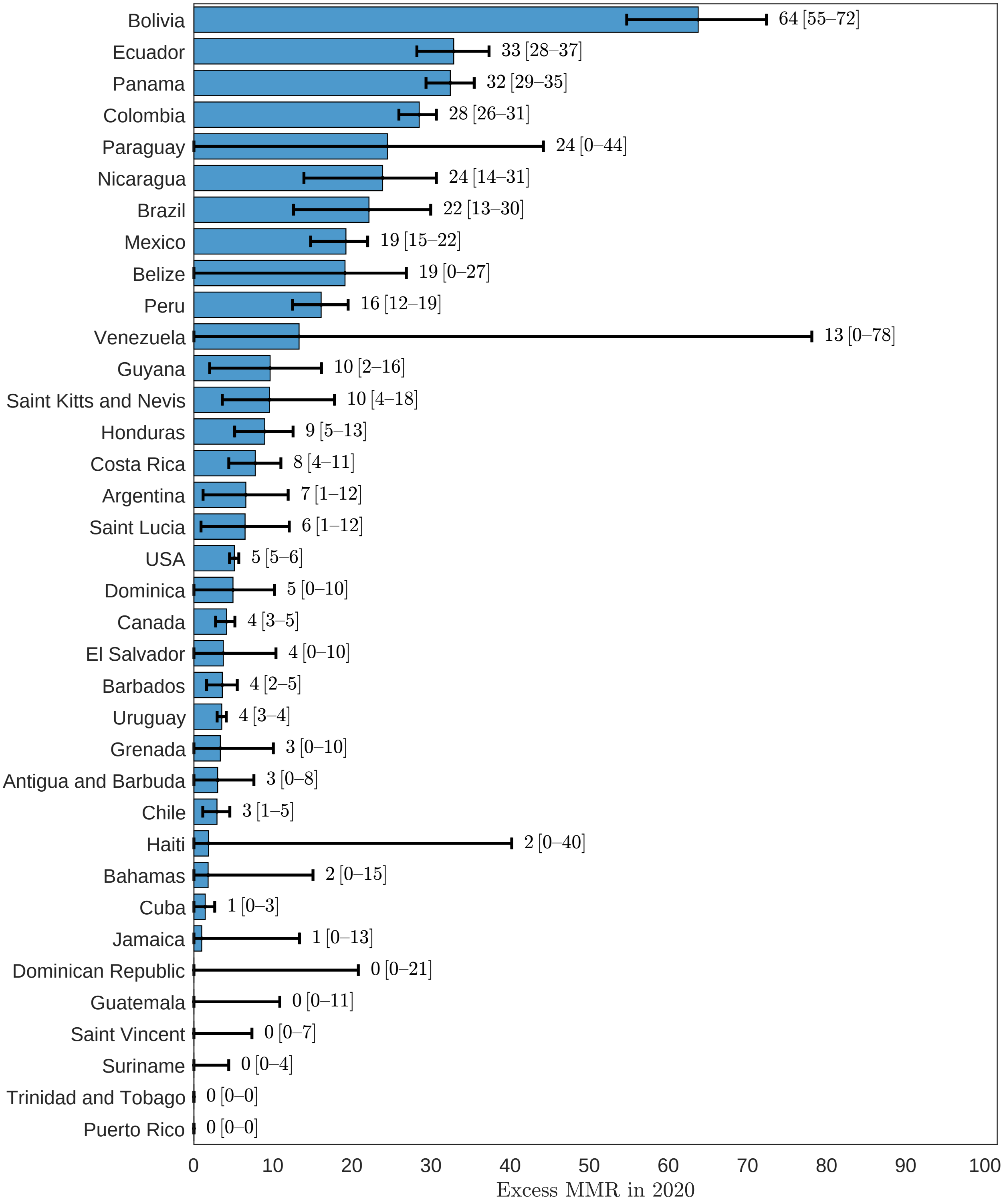


eFigure 71: Excess maternal mortality ratio, Region of the Americas, 2020.

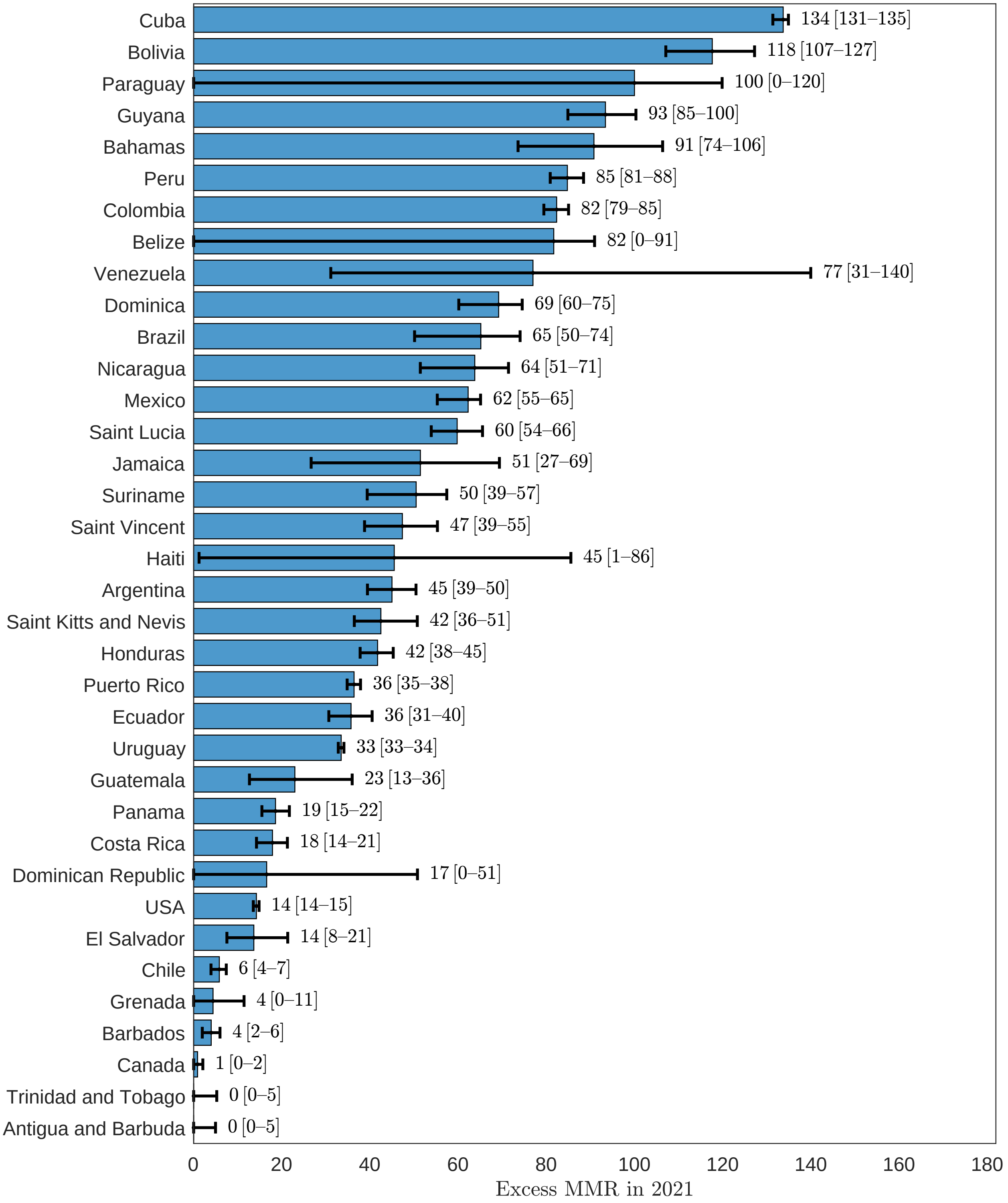


eFigure 72: Excess maternal mortality ratio, Region of the Americas, 2021.

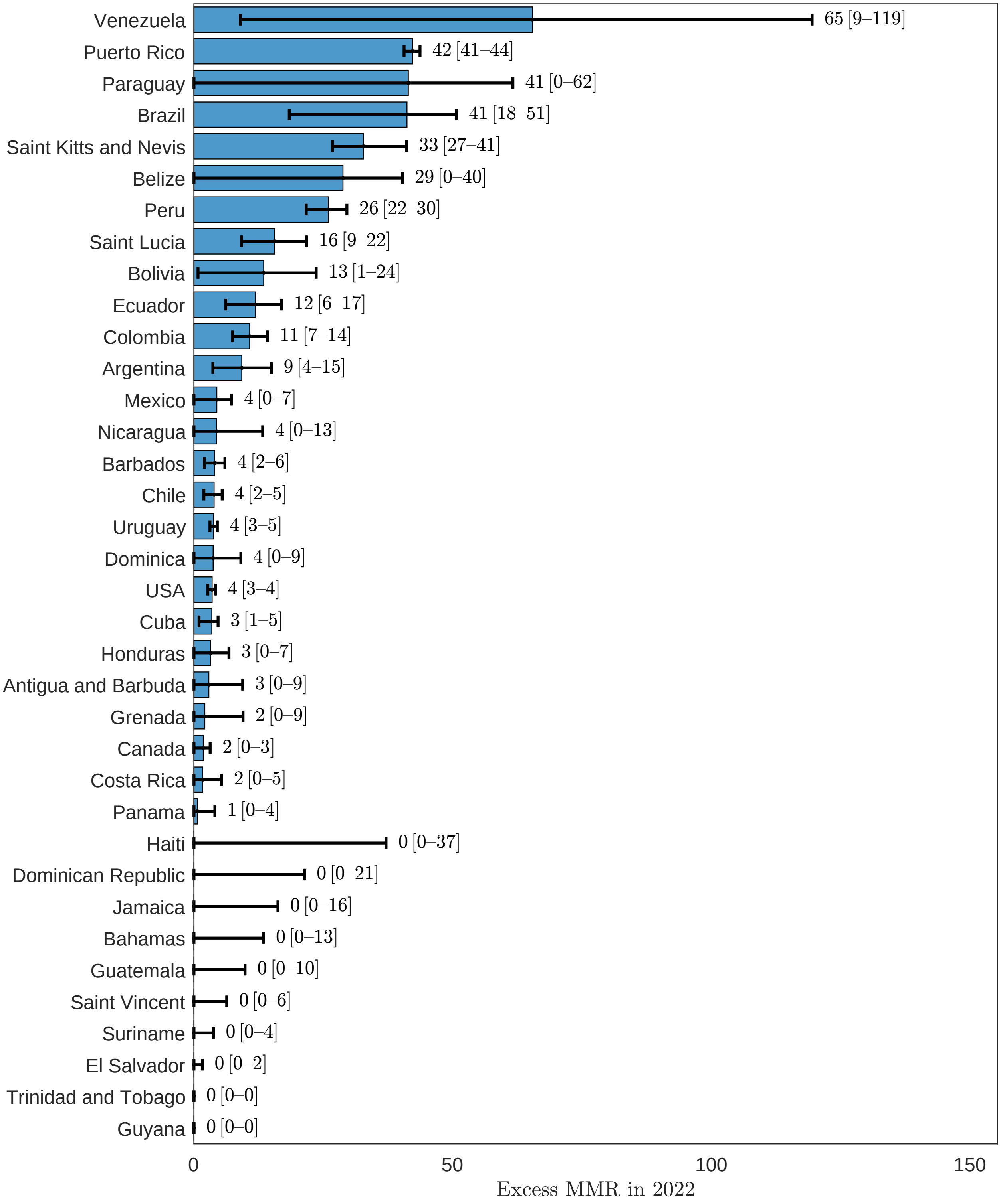


eFigure 73: Excess maternal mortality ratio, Region of the Americas, 2022.

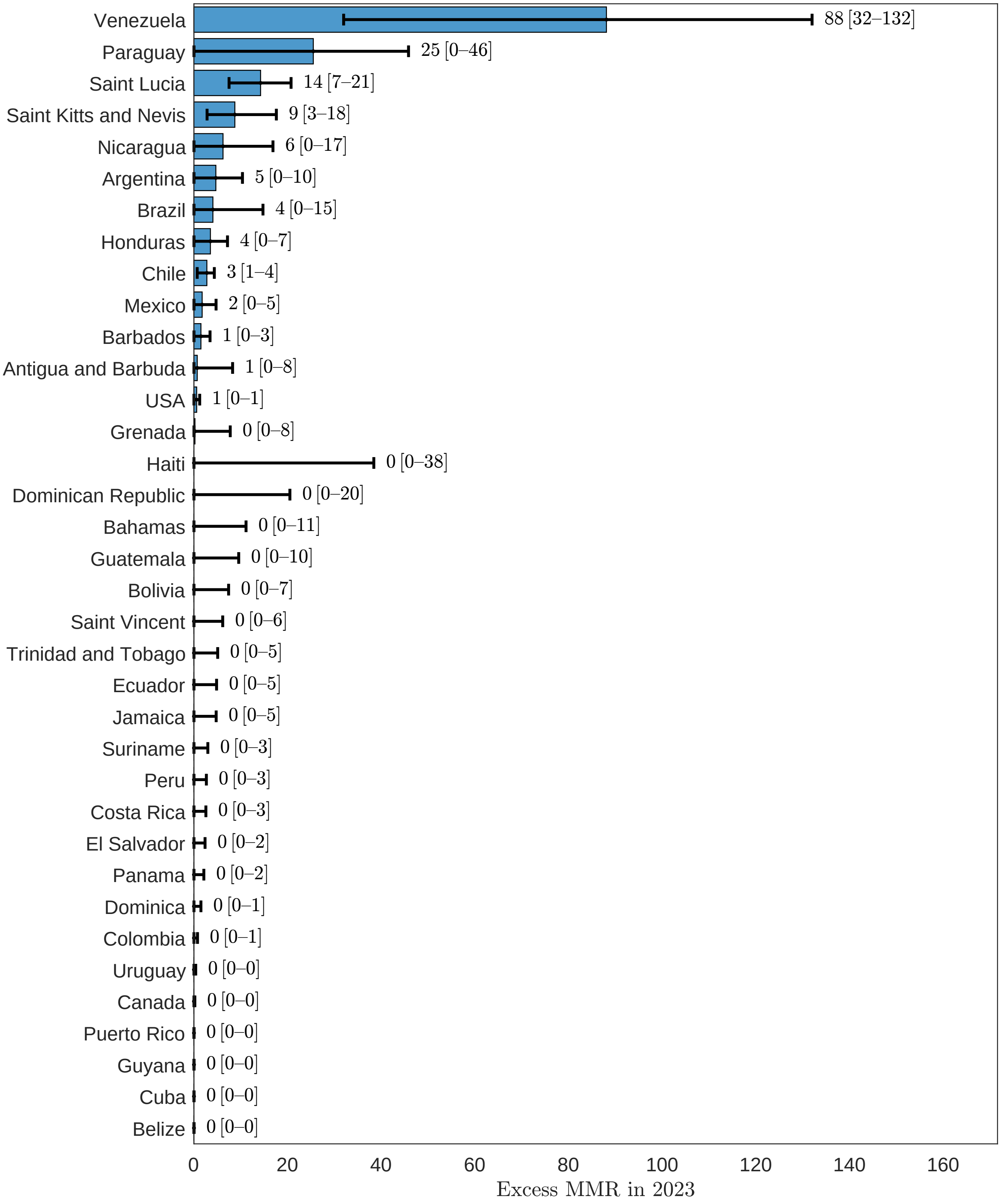


eFigure 74: Excess maternal mortality ratio, Region of the Americas, 2023.

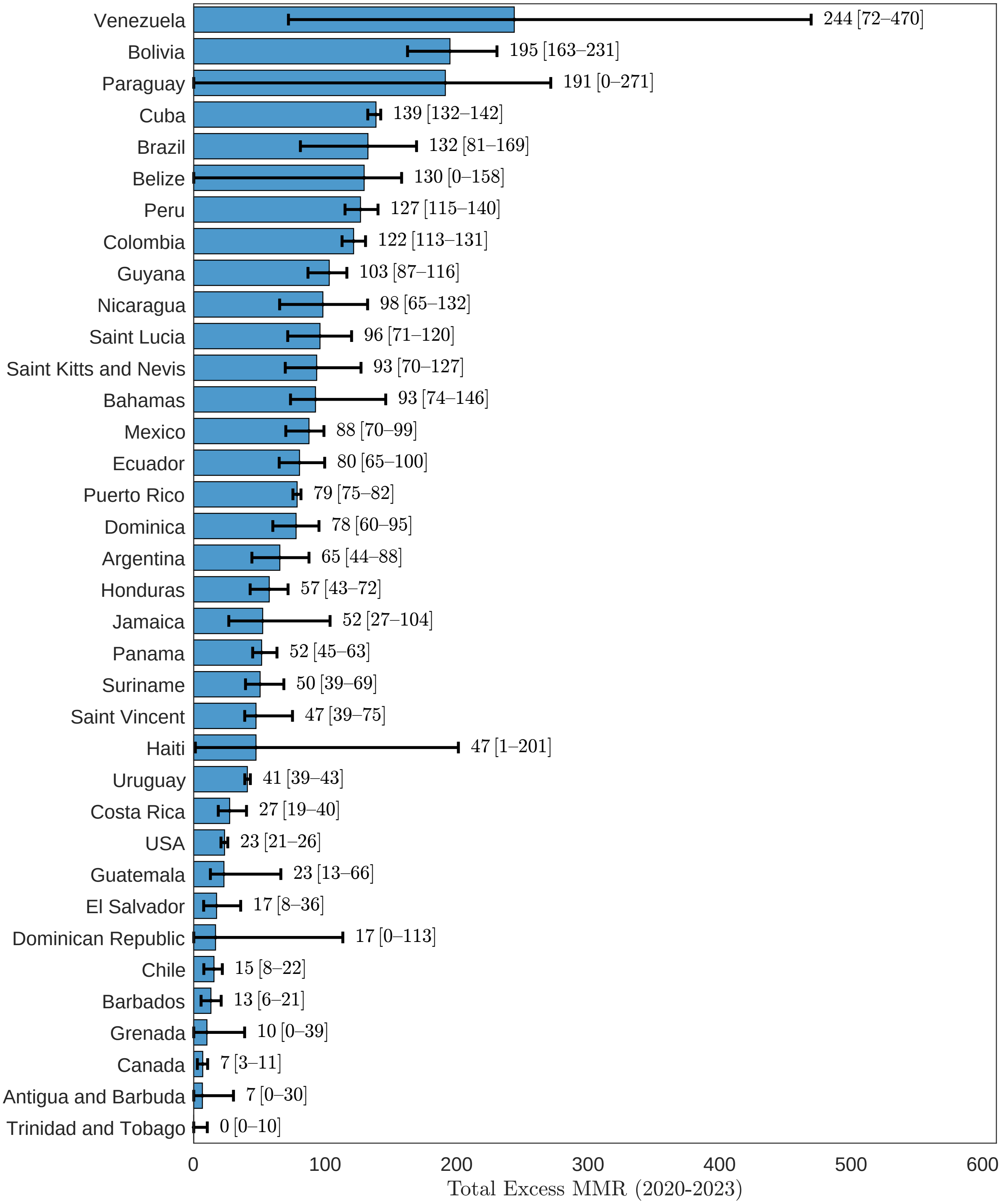


eFigure 75: Excess maternal mortality ratio, Region of the Americas, 2020-2023.

#### 3.7 Illustrative high-burden settings

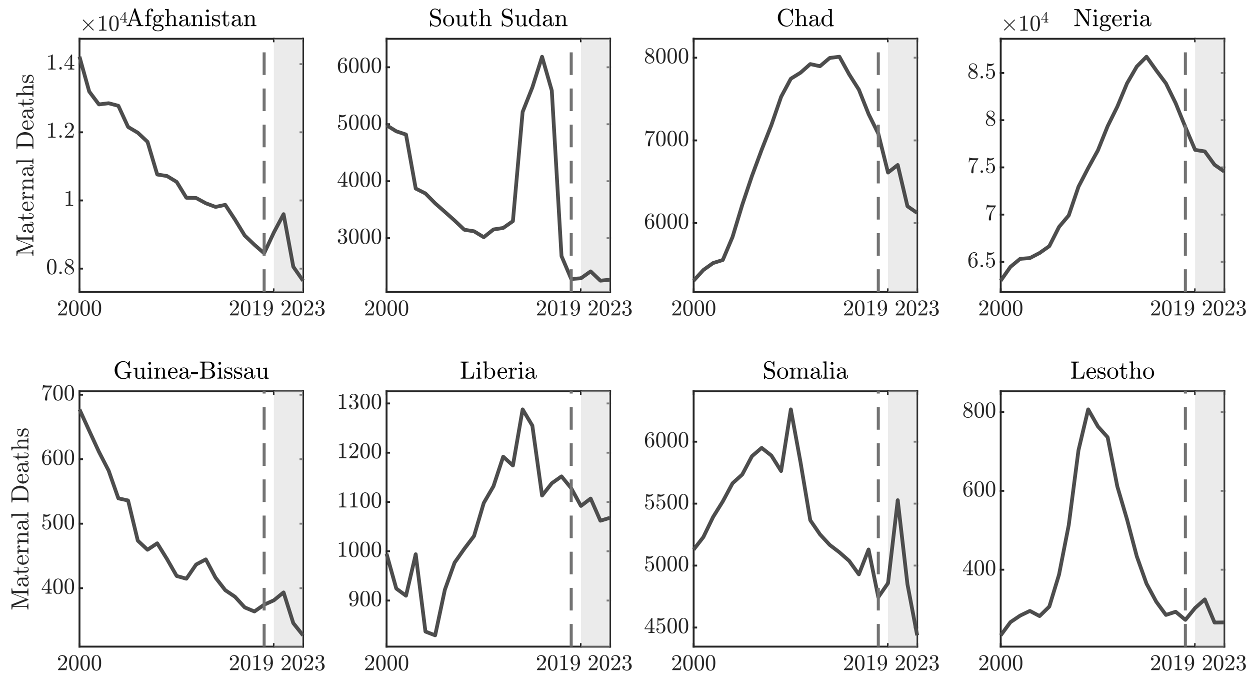


eFigure 76: Annual maternal death estimates in the eight illustrative high-burden settings.

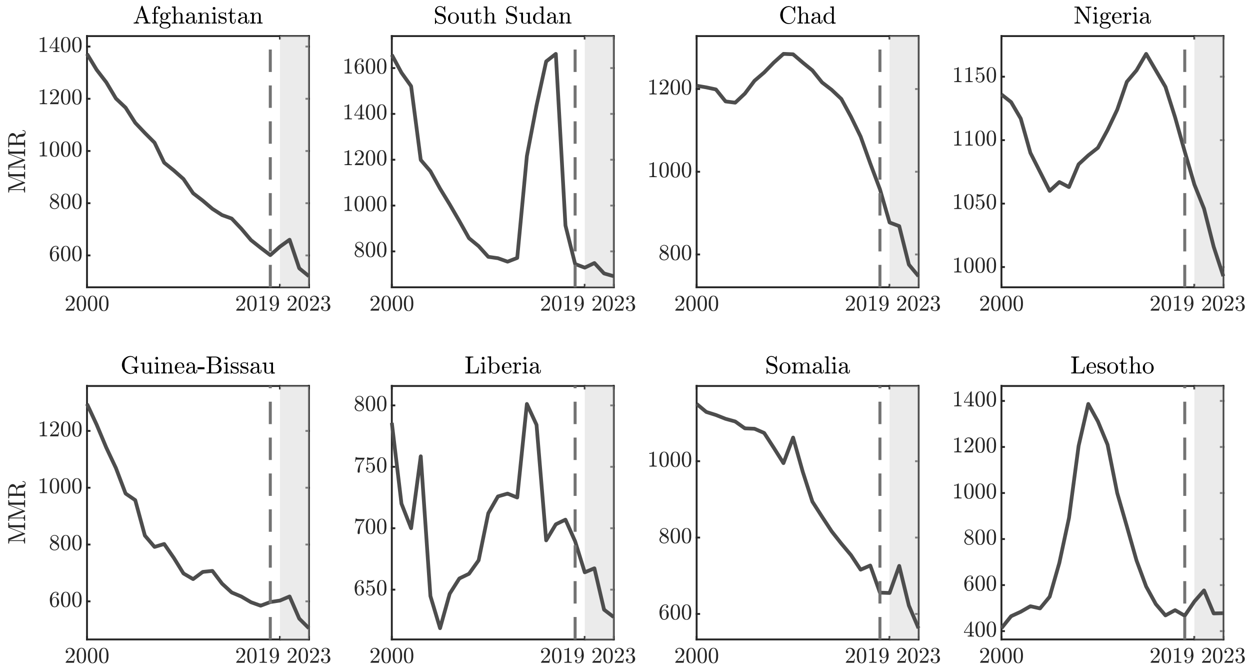


eFigure 77: Annual maternal mortality ratio estimates in the eight illustrative high-burden settings.

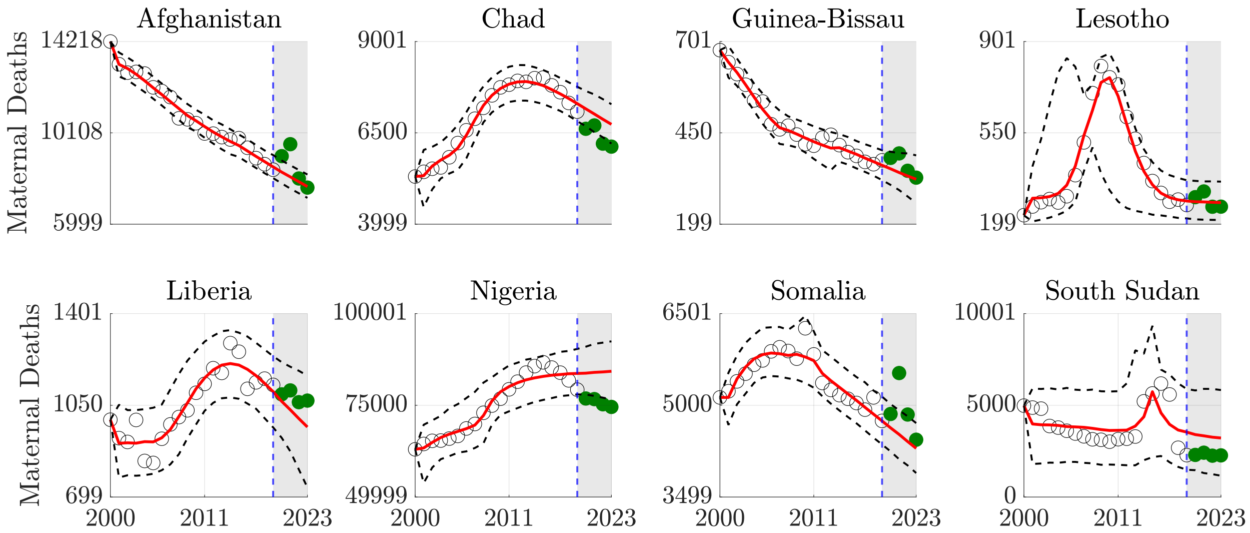


eFigure 78: Counterfactual maternal death forecasts in the eight illustrative high-burden settings.

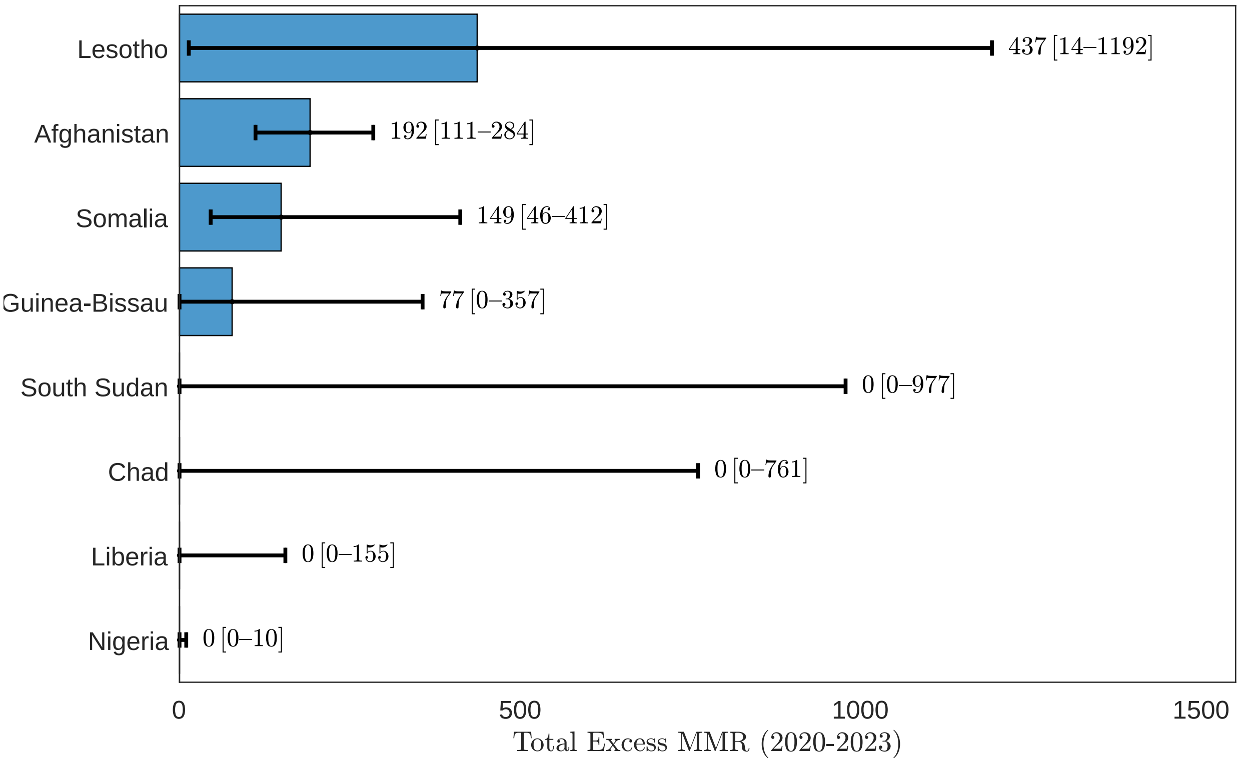


eFigure 79: Cumulative excess maternal mortality ratio in the eight illustrative high-burden settings, 2020-2023.

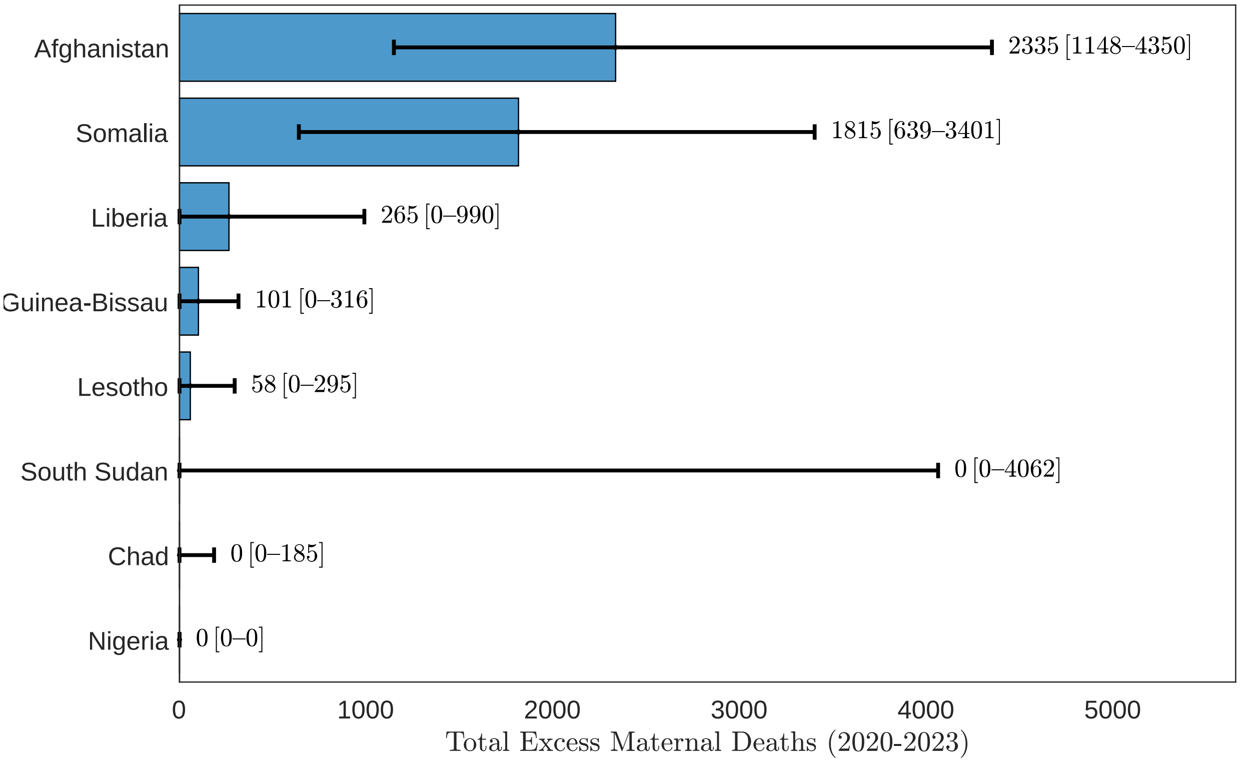


eFigure 80: Cumulative excess maternal deaths in the eight illustrative high-burden settings, 2020-2023.
